# Cross-omic dissection reveals locus-specific heterogeneity and antagonistic pleiotropy between Alzheimer’s disease and type 2 diabetes

**DOI:** 10.64898/2026.03.23.26349030

**Authors:** Emmanuel O Adewuyi, Asa Auta, Olayinka S Okoh, Kaja Selmer, Kristina Gervin, Dale R Nyholt, Gavin Pereira

## Abstract

Observational studies associate type 2 diabetes (T2D) with increased dementia risk; however, the specificity of this relationship to Alzheimer’s disease (AD) and its biological basis remains unresolved. We apply an integrative cross-omic framework to dissect genetic links between AD and T2D. Genome-wide analyses reveal a modest positive genetic correlation and robust polygenic sign concordance of AD with T2D. High-resolution analyses demonstrate locus-specific heterogeneity, with coexisting positive and predominantly negative correlations, and strong inverse associations at *APOE* and *HLA*. Cross-trait GWAS meta-analyses indicate that most genome-wide significant signals reflect trait-specific effects, with only a limited set of variants supported in both AD and T2D. Colocalisation reveals distinct causal variants at most shared loci. Gene-based analyses highlight convergence at functional genes, including *PLEKHA1, VKORC1, ACE, and APOE*, without implying concordant variant-level effects. Bidirectional Mendelian randomisation (MR) shows no evidence of a causal relationship between AD and T2D in either direction. Summary-data MR prioritises genes whose expression or methylation affects both AD and T2D, mostly with opposing effects. Only *PLEKHA1* (eQTL) and *CAMTA2* (mQTL) show concordant positive associations. Five genes, *GALNT10, HSD3B7, BCKDK, KAT8*, and *ACE*, are supported across both regulatory layers, while numerous signals cluster within a regulatory hotspot at 16p11.2, supporting convergent transcriptional and epigenetic involvement, despite directional divergence. These results refine the AD–T2D relationship; rather than a simple shared-risk model, overlap reflects locus-specific heterogeneity and cross-omic convergence often showing opposing effects on AD versus T2D risk, consistent with antagonistic pleiotropy.

## Introduction

Alzheimer’s disease (AD) is the most common cause of dementia, accounting for approximately 60–80% of cases worldwide^1^. With population ageing, its prevalence continues to rise, and it now constitutes a major and growing global public health challenge^1-3^. Type 2 diabetes mellitus (T2D), a chronic metabolic disorder with a similarly increasing global burden^4-6^, has been described as ‘one of the fastest growing health emergencies of the 21st century’^6^. In 2024, an estimated 589 million adults lived with diabetes worldwide, a number projected to reach 853 million by 2050, with T2D comprising over 90% of all cases^7,8^. A substantial epidemiological literature links T2D to an increased risk of late-life cognitive impairment, with observational studies consistently reporting elevated dementia risk among individuals with diabetes^9-13^. However, contrasting evidence also exists^14,15^, and findings are not fully uniform; reported effect sizes vary considerably, particularly when distinguishing AD from vascular or mixed dementias^9-13,16,17^. Meta-analyses consistently support an association between diabetes and dementia overall^9-12^, although estimates specific to AD are generally more modest and heterogeneous. These findings reinforce the epidemiological association while leaving unresolved the extent to which T2D specifically, rather than diabetes more broadly, increases risk for AD as distinct from other dementia subtypes.

Biological evidence provides plausibility for a relationship between T2D and AD. For instance, Insulin and insulin-like growth factor-1 signalling in the brain play central roles in synaptic plasticity, neuronal survival, and aspects of amyloid-β metabolism^18-20^. Similarly, post-mortem studies show impaired brain insulin signalling in AD^21^, contributing to the concept of brain insulin resistance^17,21^. Metabolic disturbances characteristic of T2D, including hyperglycaemia, oxidative stress, mitochondrial dysfunction, dyslipidaemia, and systemic inflammation, can exacerbate cerebrovascular injury and interact with neurodegenerative processes^16,20,22^. However, clinicopathological studies indicate that diabetes is more strongly associated with cerebrovascular lesions than with classical AD hallmarks^23-25^. This position is echoed in other studies, supporting the view that much of the diabetes–dementia association reflects vascular or mixed lesions rather than typical AD features ^24,25^. These findings argue against a single, uniform mechanism linking T2D to AD and underscore the need to disentangle disease-specific from vascular pathways.

Human genetic studies add further resolution to this picture. Large-scale genome-wide association studies (GWAS) have identified numerous susceptibility loci for AD and hundreds for T2D, reflecting the heterogeneous biology of each disorder^26-30^. However, overlap among genome-wide significant loci is limited, and most genome-wide analyses report weak or non-significant global genetic correlation^31,32^. Consistent with this position, Hardy et⍰al. reported no global genetic correlation, no cross-trait replication of genome-wide significant loci, and no association between T2D polygenic risk scores and AD^33^. Conversely, several studies have reported shared genetic overlap or biological pathways between AD and T2D^17,32,34-37^, whereas others argued that they represent largely distinct disorders^14,15,38^, highlighting substantial and sometimes conflicting evidence. Mendelian randomisation (MR) mirrors this complexity, with consistent evidence indicating that genetic liability to T2D shows no strong causal effect on AD^39^. Together, these findings further highlight unresolved aspects of the AD–T2D relationship and suggest that global metrics can obscure locus-specific or trait-aligned mechanisms, including effects that may operate in opposite directions across the genome.

Although pleiotropy is pervasive in cross-trait genetic architecture^40-44^, its directionality and regulatory basis across AD and T2D remain poorly characterised. For instance, it is largely unclear whether shared loci exert concordant (same-direction) or antagonistic (opposite-direction) effects, and which regulatory mechanisms drive these relationships. Antagonistic pleiotropy is plausible at the interface of metabolism, immunity, and neurodegeneration, including canonical loci such as *APOE* and *MHC* immune regulatory regions^16,45,46^. We perform an integrative, multi-method investigation of AD–T2D genetics, combining genome-wide and locus-specific analyses with transcriptomic and epigenomic regulatory evidence. Using several well-powered datasets, we quantify global and local genetic correlation, perform heterogeneity-aware cross-trait meta-analysis (RE2)^47,48^, and conduct bidirectional MR. RE2, incorporating m-values and binary effect support, accommodates cross-trait heterogeneity, separating trait-specific signals from single-variant pleiotropy. Expression- and methylation-based summary-data MR (SMR)^49,50^ prioritise putative regulatory drivers and systematically assess concordant and discordant effects.

Our multi-layered, locus-resolved, cross-omic framework moves beyond global summary metrics to identify where concordant and putative antagonistic pleiotropy potentially operate and to prioritise candidate genes for functional follow-up. We emphasise that genome-wide metrics such as global correlation^31^ or effect concordance^51^ capture diffuse polygenic covariance, whereas locus-resolved approaches^52^ reveal regional heterogeneity and locally concordant or discordant effects, and colocalisation distinguishes shared from distinct causal variants^53^. These pleiotropic patterns do not imply causality: MR tests genetically mediated causality and may yield null results even when substantial biological overlap exists. Accordingly, genome-wide overlap, locus-specific directionality, and causal inference should be viewed as complementary rather than contradictory dimensions of cross-trait architecture. Our study demonstrates that AD and T2D share a complex, multilayered genetic architecture in which modest genome-wide correlation coexists with pronounced locus-specific heterogeneity. We found no evidence of a causal association; most genome-wide significant signals were trait-specific, and Bayesian colocalisation showed that overlapping loci were largely driven by distinct causal variants. SMR analyses further indicated that most shared regulatory signals act in opposite directions, with only a small set of concordant cross-omic candidates identified for further investigation.

## Results

### Study design overview

Figure 1 summarises our integrative, multi-layer cross-omics analytical framework for dissecting shared genetic and regulatory architecture between AD and T2D. We first quantified genome-wide polygenic overlap using linkage disequilibrium score regression (LDSC)^31^ and assessed SNP effect concordance as well as SNP-level genetic overlap across p-value thresholds using SNP effect concordance analysis (SECA)^51^.

**Figure 1:**
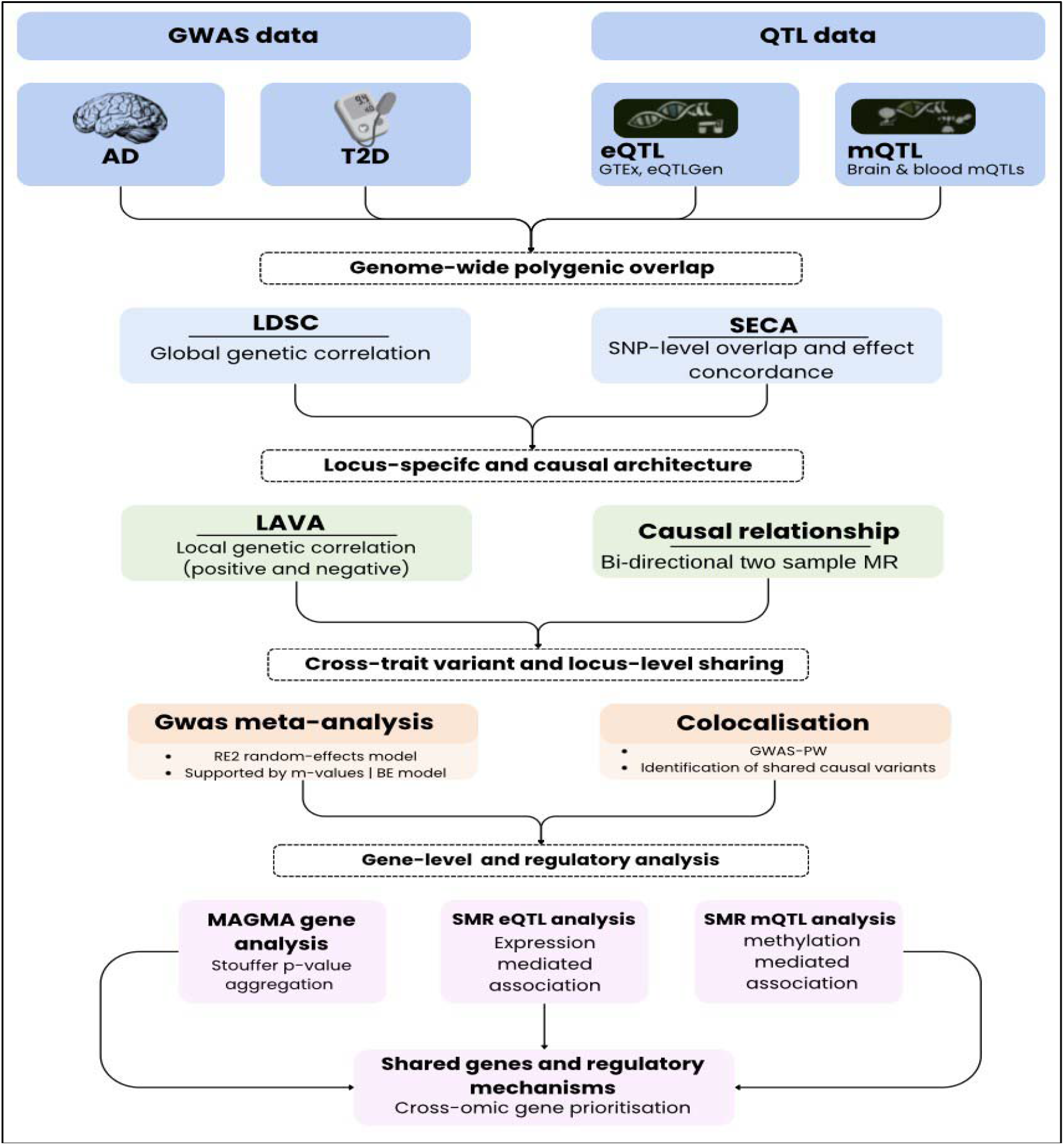
Study overview and workflow for dissecting the relationship between AD and T2D. Workflow integrating Alzheimer’s disease (AD) and type 2 diabetes (T2D) GWAS with expression QTL (eQTL) and methylation QTL (mQTL) datasets. Genome-wide overlap was assessed using linkage disequilibrium score regression (LDSC) and SNP effect concordance analysis (SECA). Local genetic architecture was evaluated using local analysis of [co]variant association (LAVA) and bidirectional two-sample Mendelian randomisation (MR). Cross-trait variant and locus-level sharing was examined through RE2 random-effects meta-analysis and GWAS-pairwise (GWAS-PW) colocalisation. Gene-level and regulatory analysis was assessed using MAGMA gene-based tests and summary-data–based MR (SMR) integrating eQTL and mQTL data.

We then characterised locus-specific genetic architecture using LAVA (Local Analysis of [co]Variant Association) to estimate regional correlation and directionality^52^. We investigated cross-trait variant- and locus-level sharing using the modified random-effects GWAS meta-analysis (RE2), supported by diagnostic m-values and the binary-effects models^47,48^. Bayesian colocalisation was used to distinguish shared from trait-specific causal signals^53^. Causal relationships were assessed using bidirectional two-sample MR with multiple estimators and sensitivity analyses^54,55^. At the gene and regulatory levels, we integrated gene-based association testing (in MAGMA)^56^, p-value aggregation (Stouffer)^57^, and SMR^49,50^ across expression and methylation quantitative trait loci (QTL) datasets to prioritise candidate genes^29,30^.

### Summary of data sources

We used AD GWAS summary data (71,880 cases and 383,378 controls), including both clinically diagnosed and proxy-defined AD cases^26^. We conducted a potential (partial) replication using a dataset restricted to clinically diagnosed AD cases (17,008 cases and 37,154 controls^29^). T2D summary statistics were from the DIAGRAM consortium (74,124 cases and 824,006 controls^28^). We obtained additional T2D datasets from Xue *et*⍰*al*.^30^, UK Biobank, and FinnGen (release R10)^58^ for replication testing. All participants were of European ancestry. Lastly, we used cis-eQTL summary statistics from GTEx v8, eQTLGen, and BrainMeta, and cis-mQTL data from BrainMeta and the McRae blood mQTL resource, providing complementary regulatory coverage across brain and blood tissues. The methods and Supplementary Table 1 provide more information on data sources.

### Genome-wide genetic correlation between AD and T2D

We conducted a comprehensive analysis of the global genetic correlations between AD and T2D using data from multiple studies and cohorts. Our findings reveal consistently positive and statistically significant genetic correlations between the two disorders (Figure 2 and Supplementary Table 2). First, utilising GWAS data encompassing clinically diagnosed and proxy cases of AD^26^, we observed a significant global genetic correlation with T2D ^28^ (rG = 0.14, P = 1.43 × 10^-4^), and BMI-adjusted T2D (rG = 0.13, P = 2.44 × 10^-4^). This result was validated using additional T2D GWAS datasets from the UK Biobank (rG = 0.19, P = 7.68 × 10^-5^), Xue et al^30^ (rG = 0.17, P = 3.81 × 10^-8^) and Finngen (rG = 0.10, P = 1.05 × 10^-4^). Focusing solely on the clinically diagnosed cases of AD^29^, we similarly replicated a positive and significant global genetic correlation pattern with T2D (Figure 2 and Supplementary Table 2). Notably, the global genetic correlation between AD and T2D remained positive and more statistically significant even after excluding the *APOE* region (Figure 2 and Supplementary Table 2). Supplementary Table 3 summarises pairwise genetic correlations between different T2D GWAS summary-statistics datasets used in our study. The results show high estimates and highly significant genetic correlations, indicating these T2D GWAS largely capture the same underlying genetic signal.

**Figure 2:**
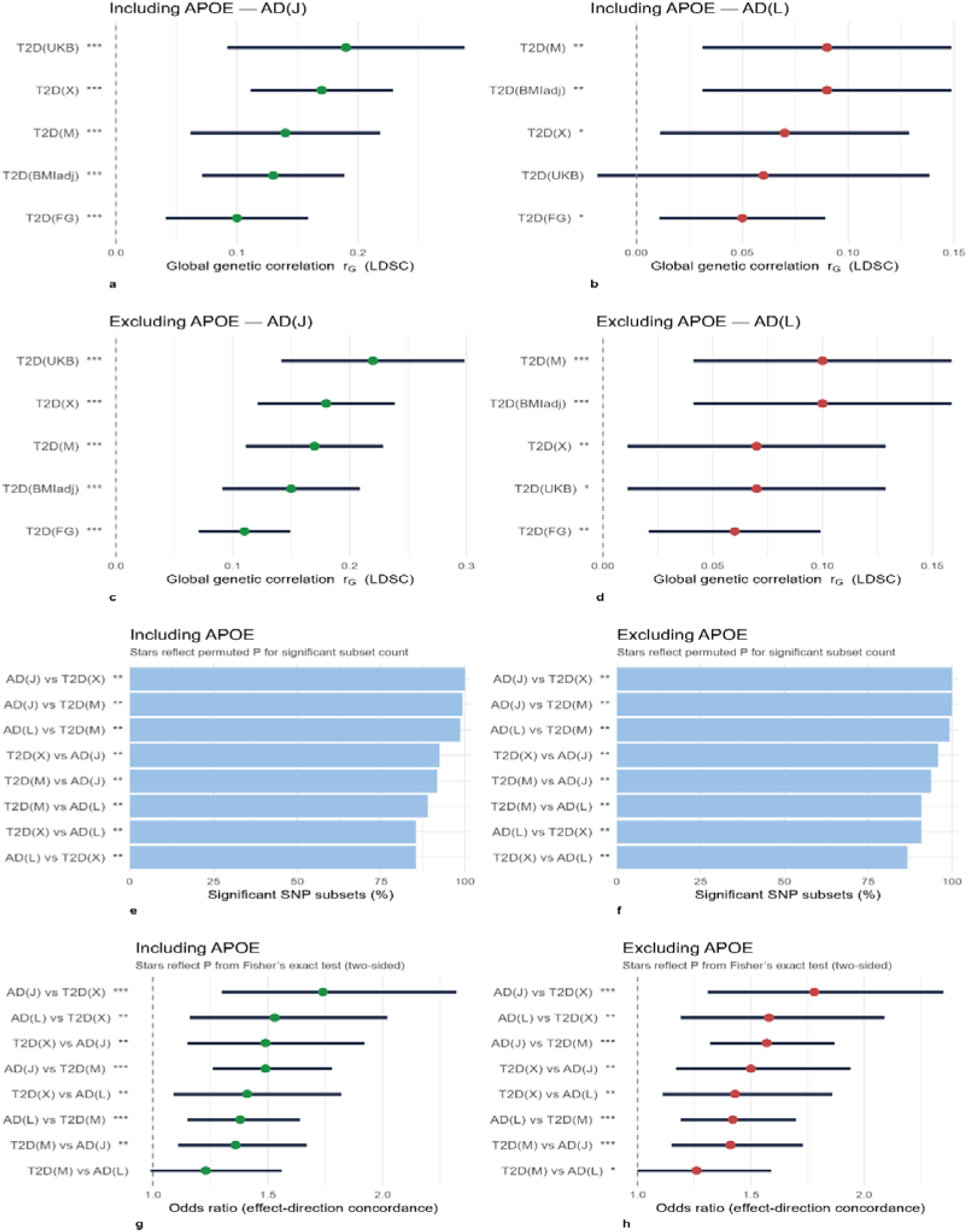
Global genetic correlation and SNP effect concordance between Alzheimer’s disease and type 2 diabetes. Panels a–d show global genetic correlations between AD and T2D estimated using LDSC. Results are shown, including (a–b) and excluding (c–d) the APOE region. Panels are stratified by AD GWAS: AD(J) denotes the GWAS (combining clinically diagnosed and proxy cases), and AD(L) denotes the GWAS (clinically diagnosed cases only). Points represent the genetic correlation estimate (r_G_), horizontal lines indicate 95% confidence intervals, and the vertical dashed line marks r_G_⍰=⍰0. Panels e–h show results from SECA for AD–T2D GWAS pairs, again including (e, g) and excluding (f, h) the APOE region. Panels e–f display the percentage of SNP subsets with significant concordant effects between traits (out of 144 tested subsets). Panels g–h display ORs quantifying the association between SNP effect directions in AD and T2D, with 95% confidence intervals and a dashed vertical line indicating OR⍰=⍰1. In the SECA bar plots, asterisks reflect permuted p-values for observing the number of significant concordant SNP subsets. Permuted P is the p-value for observing significant SNP subsets in the observation. SECA calculates the permuted P value for the number of significant associations with adjustment for testing 144 associations (based on permutations of 1000 replicates). When testing for the association between effect direction in Trait 1 SNPs and Trait 2 SNPs at P_SNP_ < 0.05, we used Fisher’s exact test (two-sided). Note: The SNP effects in trait 1 and trait 2 were positively correlated; given that it is significant, the results indicate the presence of allelic effects that increase the risk for both traits. Asterisks denote statistical significance using conventional thresholds (⍰p⍰<⍰0.05; **⍰p⍰<⍰0.01; ***⍰p⍰<⍰0.001). In LDSC analyses (panels a–d), asterisks reflect the p-value testing whether the global genetic correlation (r_G_) differs from zero. In SECA analyses (panels e–h), asterisks reflect either permuted p-values for the number of significant concordant SNP subsets (bar plots) or two-sided Fisher’s exact test p-values for association between SNP effect directions (forest plots). Abbreviations: AD(J)^26^: Alzheimer’s disease GWAS by Jansen et⍰ al., AD(L)^29^: Alzheimer’s disease GWAS by Lambert et⍰al., LDSC: linkage disequilibrium score regression, SECA: single nucleotide polymorphism effect concordance analysis, T2D(M)^28^: T2D GWAS by Mahajan etIZal., T2D(BMIadj)^28^: T2D GWAS adjusted for body-mass index, T2D(UKB): UK Biobank T2D Phecode 250.2, T2D(X)^30^: T2D GWAS by Xue et⍰al., T2D(FG)^58^: FinnGen Release⍰10 T2D.

### SNP effect concordance and genetic overlap assessment results

We further explored the relationship between AD and various cohorts of T2D, utilising SECA^51^. This tool has some notable strengths, including the potential for assessing effect concordance, SNP-level overlap, and bidirectional relationships^51^. First, we assessed concordance in the SNP effect and tested for the association between the direction of effect (Supplementary Table 4). Of the 144 SNP subsets tested using AD (Jansen et al)^26^ as dataset 1 (12 P-value thresholds) and each T2D GWAS (Mahajan et al^28^ and Xue *et al*.^30^) as dataset 2 (12 thresholds), we observed consistent and statistically significant concordance of effect directions across all SNP subsets (Fisher’s exact test OR > 1, P < 0.05; Supplementary Table 4). Across these threshold combinations, the direction of SNP effects was consistently aligned between AD and T2D, indicating polygenic concordance. Empirical P values derived from 1,000 permutations, accounting for testing across the 144 SNP subsets, remained low for the significant results (Figure 2 and Supplementary Table 4), supporting that the observed concordance is unlikely to be due to chance. Odds ratios for the effect-direction test were greater than one across all SNP subsets, indicating an excess of SNPs with concordant effects between AD and T2D relative to expectation under the null hypothesis of no shared genetic architecture.

In a bidirectional assessment, we evaluated T2D GWASs as dataset 1 and the AD GWAS (Jansen et al ^26^) as dataset 2. This analysis similarly demonstrated consistent concordance in SNP effect directions, indicating that many of the genetic variants associated with T2D tend to show aligned effects in AD, and vice versa, irrespective of the direction of analysis. These findings further support diffuse genome-wide polygenic alignment between AD and T2D, rather than concordant locus-level or regulatory mechanisms. Restricting the analysis to clinically diagnosed AD cases (Lambert et al^29^) yielded comparable results, with strong concordance of effect directions observed (Figure 2 and Supplementary Table 4). Across multiple threshold combinations, the observed alignment of SNP effects consistently exceeded expectations under the null hypothesis, as confirmed by permutation testing, providing evidence of positive and significant effect-direction concordance between AD and T2D. Notably, results were materially unchanged when the *APOE* region was excluded, indicating that the observed concordance is not driven solely by the *APOE* locus (Supplementary Table 4).

Second, we examined SNP-level genetic overlap between AD and T2D, with the results summarised in Supplementary Table 5. We applied the binomial test to assess whether the number of SNPs jointly associated with both traits exceeded that expected by chance (P_binomial_ < 0.05). At the nominal threshold, we observed a significant excess of overlapping SNPs between AD (Jansen et al^26^) and T2D (Mahajan et al^28^). This finding was replicated using another T2D GWAS (Xue et al^30^), suggesting shared genetic architecture between AD and T2D. Reversing the direction of analysis, with T2D specified as dataset 1 and AD as dataset 2, yielded a comparable pattern. The observed proportion of overlapping SNPs significantly exceeded the expected value under the null hypothesis. Replication on the AD side using the clinically-diagnosed cases did not reach statistical significance (Supplementary Table 5), which may reflect the comparatively smaller sample size of the dataset.

At a more stringent threshold (P1 < 1 × 10^-5^ and P2 < 0.05), the degree of SNP-level overlap between AD and T2D increased, suggesting enrichment beyond chance expectations. These findings were replicated using T2D (Xue et al^30^) and largely with clinically-diagnosed AD^29^, further strengthening evidence for genetic overlap between the two traits. Importantly, significant overlap was observed regardless of whether AD or T2D served as dataset 1, suggesting that many SNPs showing stronger evidence of association with trait 1 (at P1 < 1 × 10^-5^) also tend to be associated with the corresponding trait 2. At the most stringent threshold (P1 < 5 × 10^-8^ and P2 < 0.05), that is, genome-wide significant variants in dataset 1, significant SNP overlap between AD and T2D persisted and was largely replicated. Exclusion of the *APOE* region resulted in a modest attenuation of statistical significance (likely due to the reduced number of variants for analysis) but did not alter the overall pattern or interpretation of the results (Supplementary Table 5). These findings provide further evidence of SNP-level genetic overlap between AD and T2D. Notably, SECA evaluates sign concordance aggregated across the genome and, therefore, does not necessarily imply uniform directional agreement at individual loci; consistent with this, subsequent local-correlation and colocalisation analyses revealed marked regional heterogeneity.

### Local genetic correlation between AD and T2D

Our locus-level analyses revealed marked heterogeneity in both the direction and magnitude of genetic overlap between AD and T2D, in contrast to global analyses that consistently indicated a positive and significant global genetic correlation. Several loci demonstrated reproducible local genetic correlations across at least two separate AD–T2D analysis pairs (Figure 3 and Supplementary Table 6), providing robust evidence of locus-specific genetic sharing that extends beyond chance overlap. In total, we conducted 40 local genetic correlation tests for AD (Jansen et al.)-T2D (Mahajan et al.), 17 for AD (Jansen et al.)-T2D (Xue et al.), 18 for AD (Lambert et al.)-T2D (Mahajan et al.), and five for AD (Lambert et al.)-T2D (Xue et al.), corresponding to Bonferroni-corrected significance thresholds of P = 1.25 × 10^-3^, 2.94 × 10^-3^, 2.78 × 10^-3^, and 1.00 × 10^-2^, respectively. In addition to Bonferroni-significant loci, we also considered nominally significant associations (P < 0.05) as supportive evidence when the same locus reached Bonferroni significance in a separate AD-T2D analysis pair, or when the estimated confidence interval for the local genetic correlation encompassed ±1, indicating near-complete local genetic sharing. This framework allowed prioritising biologically meaningful loci while accounting for both statistical stringency and cross-dataset validation (Supplementary Table 6).

**Figure 3:**
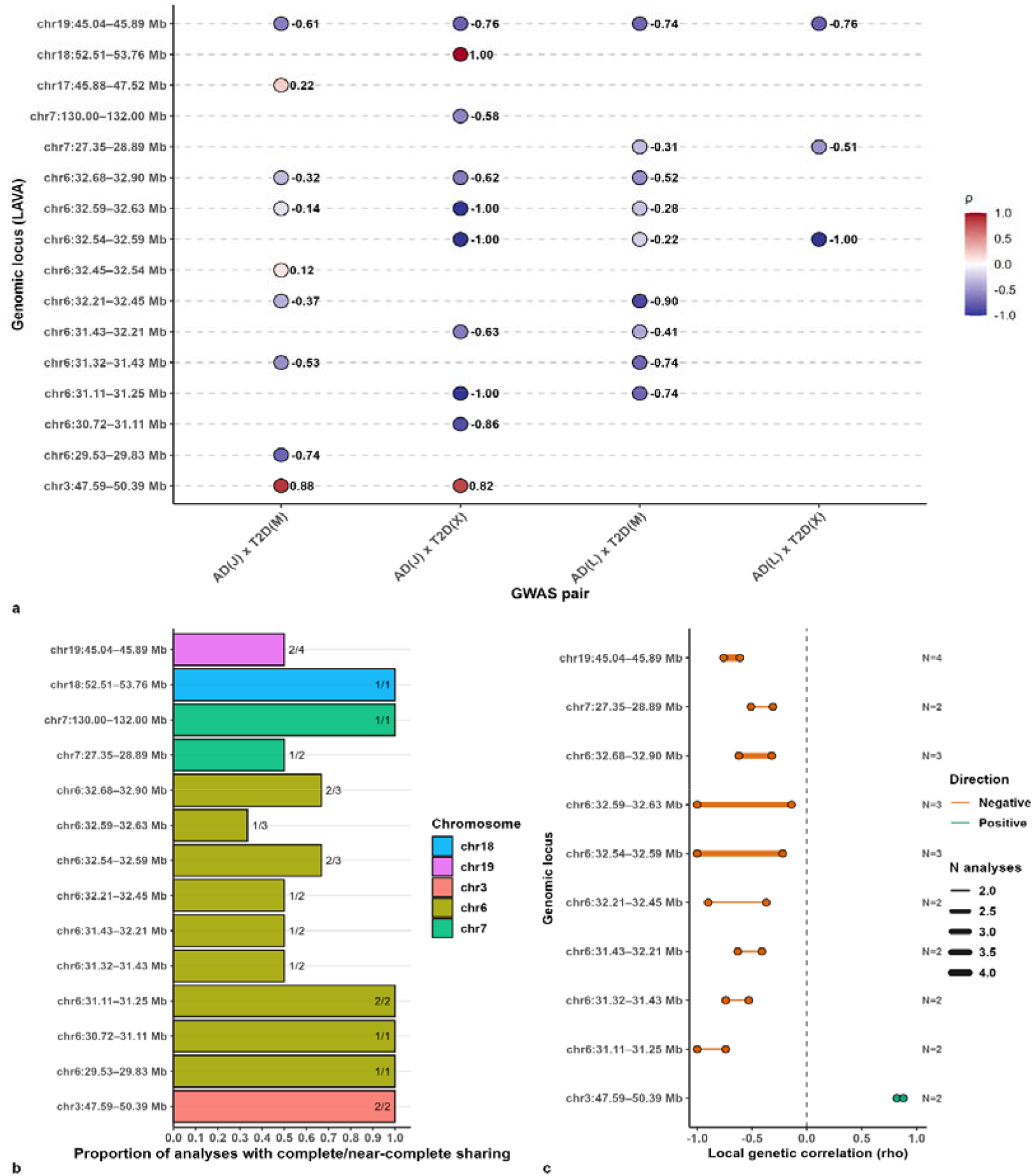
Local genetic correlation between AD and T2D across loci and phenotype pairs. **Panel a (top):** Local genetic correlation (ρ) per locus and GWAS pair. Each point represents the estimated local genetic correlation (ρ) for a given genomic locus (y axis; chr: start–stop, Mb) and GWAS pair (x axis). Points are coloured according to ρ using a diverging scale (blue, negative; white, ∼0; red, positive; range −**1** to +**1**), with numeric ρ values shown adjacent to each point. This panel summarises locus-specific genetic correlation estimates across all AD and T2D GWAS pairings. Legend indicates ρ (local genetic correlation). **Panel b (bottom left):** Proportion of analyses with complete or near-complete sharing. Horizontal bars show, for each locus, the proportion of analyses indicating complete or near-complete sharing of local genetic effects, defined as ρ = ±1 or a 95% confidence interval including ±1 in the LAVA framework. Bars are coloured by chromosome (legend: Chromosome). Fraction labels at the bar termini indicate the number of analyses supporting sharing relative to the total number of analyses per locus (N-shared / N-analyses). The physical length of each genomic interval (Mb) is annotated to the left of each row. **Panel c (bottom right):** Range of local genetic correlation (ρ) estimates by locus. Dumbbell plots summarise the minimum and maximum ρ values observed across analyses for each locus, restricted to loci with at least two significant results. Horizontal lines span from ρ_min to ρ_max, with endpoints shown as filled points. Line colour encodes the direction of correlation (positive or negative), and line thickness scales with the number of contributing analyses (legend: N analyses). Bold annotations to the right indicate the total number of analyses contributing to each locus. The dashed vertical line denotes ρ = 0. **Abbreviations and conventions:** AD, Alzheimer’s disease; T2D, type 2 diabetes; GWAS, genome-wide association study; ρ (rho), local genetic correlation; LAVA, Local Analysis of [co]Variant Association; Mb, megabases. GWAS pairs are abbreviated as AD(J) × T2D(M), AD(J) × T2D(X), AD(L) × T2D(M), and AD(L) × T2D(X), corresponding to Alzheimer’s disease GWAS by Jansen *et al*. or Lambert *et al*. and type 2 diabetes GWAS by Mahajan *et al*. or Xue *et al*., respectively. Genomic loci are displayed as chr: start–stop (Mb). Direction (Panel c): positive indicates ρ > 0; negative indicates ρ < 0. N analyses denotes the number of independent LAVA analyses contributing to each locus summary.

The most prominent and consistent signal was observed at the *APOE* locus on chromosome 19 (approximately 45.0 - 45.9 Mb). This region exhibited a strong and highly significant negative local genetic correlation across all AD-T2D pairings, including analyses using AD GWASs from Jansen et al. and Lambert et al., in combination with multiple T2D datasets (Mahajan et al. and Xue et al.). Local correlation coefficients were consistently large in magnitude, ranging from approximately ρ = -0.61 to -0.74, with corresponding P values reaching extreme levels of significance (as low as ∼ 10^-30^, Figure 3 and Supplementary Table 6). Notably, the CIs for the local genetic correlation estimates at this locus were narrow and did not overlap zero, indicating highly precise estimates. Moreover, in at least two separate AD-T2D analysis pairs, the CIs encompassed -1 (Figure 3 and Supplementary Tables 6 and 7), consistent with near-complete local genetic sharing in an inverse direction. This finding provides strong evidence of robust locus-specific putatively antagonistic pleiotropy at *APOE*, whereby genetic variants conferring increased risk for one disorder are associated with reduced liability for the other.

Beyond *APOE*, we identified additional reproducible signals within the MHC region on chromosome six. Two adjacent loci, spanning approximately 31.3 - 31.4 Mb and 32.2 - 32.4 Mb, showed consistent inverse local genetic correlations across at least two separate AD-T2D analysis pairs (Figure 3 and Supplementary Tables 6 and 8). These regions exhibited moderate to robust negative local genetic correlations, with ρ values ranging from approximately -0.37 to -0.9, and with nominal to Bonferroni-corrected statistical support (Figure 3 and Supplementary Table 6). Several additional loci on chromosomes six and seven showed a similar pattern of reproducible negative local genetic correlation across independent analysis pairs. Importantly, for many of these loci, the correlation coefficient or CIs encompassed -1, suggesting complete or near-complete local genetic sharing in an inverse direction (Figure 3 and Supplementary Tables 6 and 7). These findings imply that, within the identified regions, the same underlying genetic variants may be contributing to both disorders but with opposing effects, consistent with shared immune-mediated or inflammatory mechanisms acting antagonistically across disease contexts.

In contrast to the predominance of putative antagonistic effects, a smaller number of loci demonstrated reproducible positive local genetic correlations indicative of concordant genetic effects (Figure 3 and Supplementary Tables 6 and 8). A notable example was a locus on chromosome 3 (approximately 47.6 - 50.3 Mb), which showed a strong positive local genetic correlation (ρ > 0.8; P ≈ 10^-2^) across two AD–T2D dataset pairings. At this locus, the CIs for the local genetic correlation included +1, indicating a high degree of local genetic sharing between AD and T2D. This finding is consistent with the pleiotropic effects of shared variants influencing both disorders in the same direction. Together, our analyses demonstrate that the genetic relationship between AD and T2D is highly locus-dependent. While genome-wide analyses using LDSC and SECA indicate a broadly shared polygenic architecture with predominantly concordant directions of effect, local analyses using LAVA reveal that this global signal masks substantial regional heterogeneity. Importantly, the presence of loci with CIs encompassing ±1 provides strong evidence that some regions are driven by almost entirely shared sets of genetic variants, either acting concordantly or antagonistically across the two disorders. This combination of widespread polygenic sharing and a subset of loci exhibiting near-complete local genetic overlap may underscore the complexity of the biological mechanisms linking AD and T2D.

### No evidence of a causal relationship between AD and T2D

Our analyses reveal no convincing evidence of a significant causal association between AD and T2D, regardless of the direction of analysis: AD or T2D as the exposure or outcome variables (Supplementary Table 9). Specifically, when examining AD as the exposure and T2D as the outcome, the IVW model suggests that genetic liability to AD did not significantly affect the risk of T2D (OR = 0.98, 95%CI: 0.86 – 1.13, Supplementary Table 9). Sensitivity testing using other MR approaches, including the weighted median, MR Egger, and MR pleiotropy residual sum and outlier (MR-PRESSO), reveals consistent results of no significant causal effect of AD on T2D. Conversely, when T2D was assessed as the exposure and AD as the outcome, we found a marginally positive association (OR = 1.01, P = 1.42 × 10^-2^), which is non-convincing. The MR-PRESSO method supported this finding, suggesting a probable but non-convincing causal effect of T2D on AD. Cochran’s Q statistic revealed no significant heterogeneity, which suggests that the MR assumptions were unlikely to have been violated. The MR-Egger intercept also did not significantly deviate from zero, indicating no unbalanced pleiotropy (Supplementary Table 9). However, the 95% CI for the estimated OR ranges from 1.00 to 1.01 (for both IVW and MR-PRESSO models), indicating that the finding is not convincing. Consistent with this premise, the weighted median and MR Egger models found no evidence of a significant causal effect of T2D on AD. Notably, the findings of no causal association were consistent across validating T2D GWAS, regardless of whether AD or T2D was assessed as the exposure or outcome variables (Supplementary Table 9).

### Meta-analysis of AD and T2D GWAS

We performed cross-trait GWAS meta-analyses of AD and T2D with priority for the Han and Eskin RE2 model, which is robust to between-trait heterogeneity. We conducted primary assessments aimed at: (1) identifying SNPs and loci that were GWS (sentinel) in one or both traits and showed evidence of association with the other following meta-analysis, and (2) characterising SNPs and loci not GWS in either trait but reaching this threshold under meta-analysis.

### AD and T2D sentinel SNPs and loci showing association in GWAS meta-analysis

We first examined SNPs that were already GWS in both the AD and T2D GWAS to determine whether these signals represented potentially shared loci in both disorders. Several AD and T2D GWS SNPs showed strengthened evidence of association under meta-analysis (P_AD_ < 5 × 10^-8^ > P_RE2-meta-analysis_ < P_T2D_ < 5 × 10^-8^). Significant associations were confined to two genomic regions: on chromosome 6, within the extended HLA region, and on chromosome 19, encompassing the *APOE* locus (Supplementary Table 10). The chromosome 6 locus (chr6:32,340,070–32,667,957) comprised two lead SNPs, rs9275095 and rs9271548. The chromosome 19 locus spanned chr19:45,387,459–45,422,946 with a single lead SNP, rs12972156. Highly significant RE2 p-values indicated increased statistical support at loci already GWS in both disorders, consistent with improved detection under a heterogeneity-aware model. However, evaluation of BE p-values and m-values revealed limited support for pleiotropy at these loci. For instance, at most loci, m-values supported effect in only one trait (m-value ≥ 0.9) with absence in the other (m-value ≤ 0.1), indicating trait-specific associations at the individual SNP level. Specifically, variants within the HLA region showed strong evidence of association with T2D but minimal evidence in AD, whereas variants within the *APOE* region were AD-specific (Supplementary Table 10). BE p-values were generally weaker than the corresponding RE2 p-values, suggesting that these signals were not driven by shared single-variant effects acting concordantly across traits.

### AD sentinel SNPs and loci showing association with T2D

We next examined SNPs and loci that were GWS in the AD (sentinel) and at least nominally associated in T2D, to assess whether they showed evidence of association under cross-trait meta-analysis. Based on the improved RE2 p-values, multiple AD sentinel SNPs showed evidence of association with T2D (Supplementary Table 11). These SNPs clustered into three loci: chromosomes 6 (chr6:32,340,070–32,667,957), 11 (chr11:121,435,587–121,451,813), and 19 (within the *APOE* region). On chromosome 6, the SNPs collapsed into a single locus, with rs9271548 and rs9275095 as lead SNPs, consistent with extensive LD and allelic heterogeneity. On chromosome 19, the SNPs mapped to two distinct loci. The first (chr19:45,241,638– 45,422,946) comprised multiple SNPs with rs2927437 as the lead. A second, independent locus was at chr19:45,734,751 (lead SNP: rs62118504). Both chromosome 19 loci showed highly significant RE2 p-values, driven primarily by AD with weak evidence of association in T2D. Across chromosomes 6 and 19, BE p-values and m-values indicated limited concordant pleiotropy at the individual SNP level. Variants within the *APOE* loci were AD-specific, whereas the extended HLA region exhibited marked heterogeneity, with subsets of SNPs showing AD-specific effects, T2D-specific effects, or intermediate evidence in one trait alongside strong evidence in the other. BE p-values were generally weaker than RE2 p-values.

Notably, SNPs on chromosome 11 formed a single locus with rs11218343 as the lead. This locus showed robust evidence of association under the RE2 model, with m-values indicating strong evidence of effects in both AD and T2D, supported by significant BE p-values. This region, therefore, represents a clear example of a putatively shared genetic effect identified in the analysis. Our cross-trait meta-analysis identified multiple AD sentinel loci with evidence of association in T2D; however, robust shared genetic effects were restricted to specific regions, most notably the locus on chromosome 11, while other loci reflected heterogeneous (chromosome 6) and largely trait-specific genetic architectures (chromosomes 6 and 19).

### T2D sentinel SNPs and loci showing association with AD

The T2D sentinel variants (that are at least nominally associated with AD) mapped predominantly to the extended HLA region on chromosome 6, with additional loci identified across multiple chromosomes, including 1, 2, 3, 4, 5, 8, 10, 15, 16, 17, and 19 (Supplementary Table 12). Within the HLA region (approximately chr6:31.1–32.7 Mb), a dense cluster of SNPs collapsed into a single broad locus, reflecting extensive LD and allelic heterogeneity. Multiple established T2D lead variants were represented, including rs3130931, rs35840219, rs9271548, and rs9275095. Although RE2 meta-analytic p-values were highly significant across this region, BE p-values and m-values indicated nearly all variants showed strong evidence of effect presence in T2D (m-value ≈ 1.0) with little to no evidence of effect in AD, consistent with T2D-specific genetic effects. One HLA variant (rs4335021) exhibited intermediate m-values in AD and T2D, suggesting limited or uncertain cross-trait sharing rather than robust pleiotropy. Outside chromosome 6, the SNPs showed a consistent pattern of T2D-specific effects with minimal evidence of association in AD. Across loci on chromosomes 1, 2, 3, 4, 5, 8, 10, 15, 16, 17, and 19, RE2 significance was driven primarily by strong T2D associations, while AD signals remained weak or absent, supported by uniformly low AD m-values (≤ 0.1) and generally non-significant BE p-values. The findings suggest that the meta-analytic signals were not attributable to shared single-variant effects acting across both traits. Overall, cross-trait meta-analysis of T2D sentinel SNPs mirrored the AD sentinel pattern: RE2 identified multiple loci with cross-trait association, but signals were largely trait-specific, with little evidence of robust sharing.

### SNPs and loci reaching genome-wide significance under cross-trait meta-analysis

We examined SNPs and loci that were nominally associated with both AD and T2D in the individual GWAS but did not reach GWS in either trait alone, and instead attained significance under the RE2 cross-trait meta-analysis (Figure 4; Supplementary Table 13). These loci represent candidate novel signals emerging from cross-trait integration (based on our data). Assessment of BE p-values and m-values revealed heterogeneous support across loci; a subset showed strong evidence of shared effects, with high posterior probability of effect in both traits (m-value ≥ 0.9). Specifically, loci tagged by rs34016387 (chr5), rs35366052 (chr6), rs7131432 (chr11) and rs150775861 (chr16) demonstrated concordant support across RE2, BE p-values and m-values, consistent with putative pleiotropy, and thus represent the most compelling examples of shared genetic risk between AD and T2D among the candidate signals.

**Figure 4:**
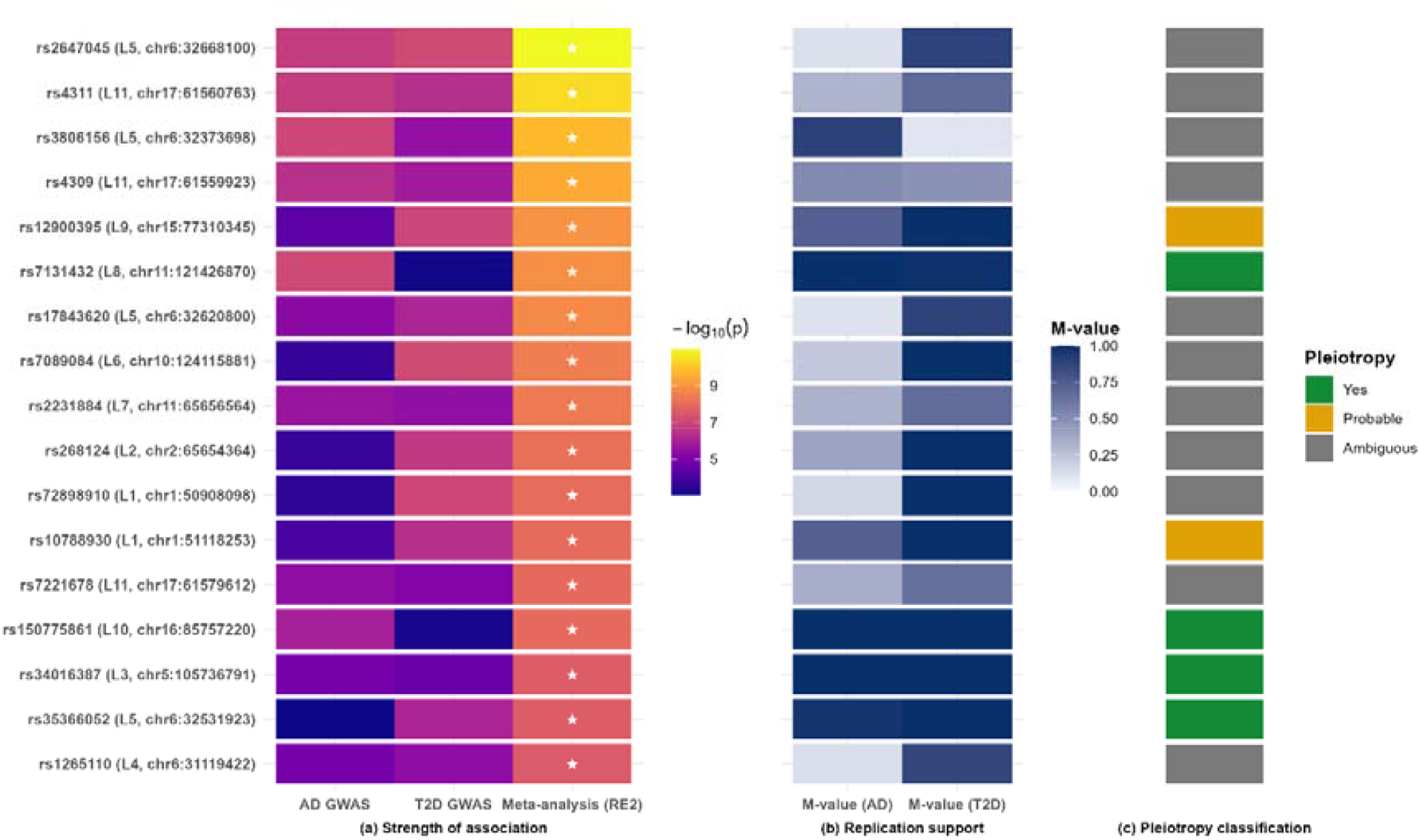
Summary of candidate ‘novel’ loci shared by AD and T2D. The figure displays independent SNPs that did not reach genome-wide significance (GWS) in the individual AD or T2D GWAS but attained significance under the RE2 model of cross-trait meta-analysis (labelled putatively ‘novel’ based on our data). SNP labels are shown as rsID (Lx, CHR: BP), where L denotes the genomic locus index defined by physical proximity and linkage disequilibrium structure, and CHR: BP indicates the chromosome and base pair position of the SNP. **(a)** Heatmap of association strength showing −log_10_(p-values) from the AD GWAS, T2D GWAS, and RE2 meta-analysis. Stars (⍰) indicate genome-wide significance (p < 5 × 10^-8^). **(b)** Trait-specific m-values (range 0– 1), representing posterior support for the presence of an effect in AD and T2D, respectively. Values close to 1 indicate strong support for association, whereas intermediate or low values indicate heterogeneous or trait-dominant architectures. **(c)** Summary pleiotropy classification integrating RE2 significance, binary effect testing, and m-value patterns. Loci classified as Yes exhibit concordant evidence of shared genetic effects across traits, Probable indicates partial but consistent support, and Ambiguous reflects discordant or intermediate support. SNPs are ordered by decreasing RE2 meta-analysis significance. SNPs showing significant improvement under meta-analysis, supported by low BE P-values (all listed are supported) and high m-values (≥0.9) for both traits, were considered strong candidates for pleiotropy. Note: These loci were designated as candidate novel loci shared by AD and T2D based on our data, as they did not reach genome-wide significance in the primary analyses but achieved genome-wide significance following meta-analysis. **Abbreviations:** AD: Alzheimer’s disease, BP: base pair position, CHR: chromosome, T2D: type 2 diabetes; GWAS: genome-wide association study; RE2: random-effects meta-analysis; SNP: single-nucleotide polymorphism; LD: linkage disequilibrium; GWS: genome-wide significance.

Using LDtrait with a European reference, a ±500 kb window and an LD threshold (R^2^ ≥ 0.6), none of the four RE2-significant pleiotropic SNPs and loci, rs34016387, rs35366052, rs7131432 or rs150775861, are in strong LD with previously reported AD or T2D GWAS index variants (Supplementary Table 14). Specifically, rs150775861 is in high LD (R^2^ ≈ 0.62) with a variant reported for colour-vision deficiency but not with AD or T2D. In contrast, rs7131432 shows only moderate LD with the established AD variant rs11218343 (R^2^ ≈ 0.34), which is below our threshold for strong tagging (Supplementary Table 14). Further assessment of rs7131432 shows that it maps to *SORL1* (g: SNPense^59^), a gene repeatedly implicated in AD^29,60^ but not in T2D (suggesting it is at least putatively novel for T2D). Further, rs150775861 maps to *C16orf74*, where nearby variants have been reported for T2D (Japanese^61^: rs377457-A, P = 6×10^-7^; European^62^: rs439967-A, P = 2×10^-6^), though none reached genome-wide significance or were associated with AD, making them putatively novel for both disorders. For the remaining SNPs and loci (rs34016387, rs35366052), we have no evidence of their association with AD and T2D, suggesting they are putatively novel for both disorders. Several additional loci show probable pleiotropy, characterised by near-certain evidence in T2D (m-value ≈ 1.0) and moderate-to-high support in AD (e.g. loci tagged by rs10788930 [chr1] and rs12900395 [chr15]), suggesting further shared effects that merit follow-up conditional and ancestry-matched LD analyses (Figure 4; Supplementary Table 13).

In contrast, many loci exhibited ambiguous support for a robust effect in both disorders. These loci reached GWS under RE2 but showed discordant or intermediate m-values between traits, indicating heterogeneous or trait-dominant architectures. This pattern was particularly evident across multiple loci on chromosomes 1, 2, 6, 10, and 11, including variants within the extended HLA region, where allelic heterogeneity and trait-specific effects were prominent. Consistent with this premise, BE p-values for some of the ambiguous SNPs were weaker than the corresponding RE2 p-values, suggesting that the meta-analytic signals were not driven by single-variant effects acting across both traits. Our cross-trait meta-analysis, thus, uncovered several GWS SNPs and loci, but robust putative pleiotropy was confined to a limited subset, highlighting the predominance of heterogeneous or trait-specific effects.

### Putatively shared AD–T2D loci and variants identified in colocalisation analyses

Using GWAS-PW, we identified multiple genomic regions associated with both AD and T2D that predominantly supported Model 4, consistent with distinct causal variants influencing each trait within the same locus. These regions were characterised by high logBF4 values and substantial PPA4, indicating strong support for locus-level sharing with variant-level heterogeneity, while recognising that high LD may limit complete discrimination from shared-variant models. In total, 18 loci met this criterion, spanning chromosomes 1, 2, 4, 6, 7, 8, 10, 11, 14, 15, 17, and 19 (Figure 5; Supplementary Table 15). These loci showed strong associations for both traits (max Z: 14.0 for AD, 11.8 for T2D), indicating Model 4 was supported by robust evidence rather than differential power (Z ≥ 5.5 = strong evidence). These results suggest that a substantial component of the genetic overlap between AD and T2D reflects locus-level convergence mediated by distinct causal variants, rather than single-variant pleiotropy.

**Figure 5:**
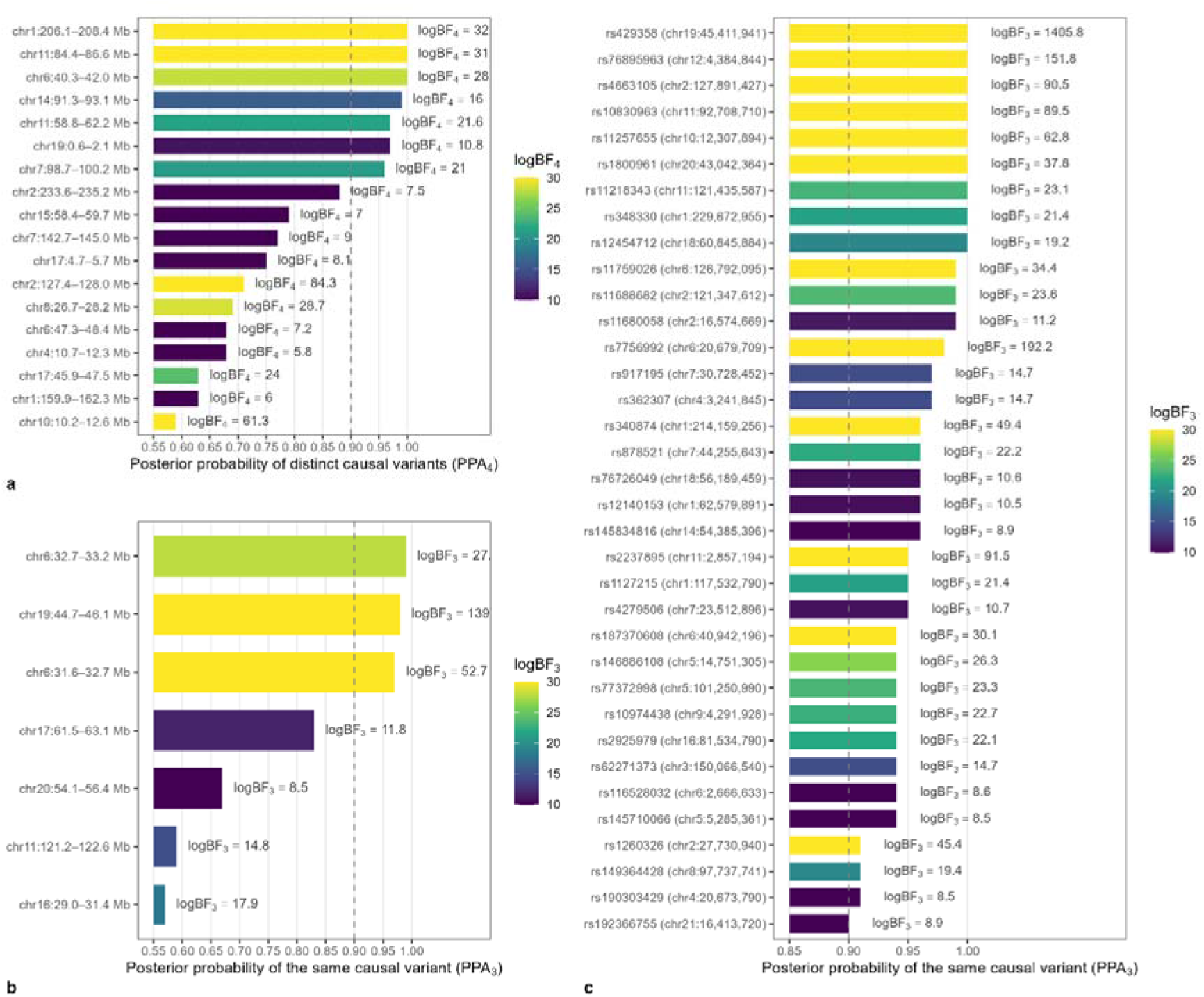
Candidate loci shared by AD and T2D with evidence of distinct, shared, or putative causal variants. This figure summarises genomic loci and representative SNPs jointly associated with AD and T2D based on Bayesian colocalisation using GWAS-PW. (a) Loci with distinct causal variants. Horizontal bars show the regional posterior probability for Model⍰4 (PPA4), corresponding to loci most consistent with distinct causal variants influencing AD and T2D within the same genomic region. Bar colour reflects the regional logBF4, with higher values indicating stronger relative support. A dashed vertical line at PPA4⍰= ⍰0.9 marks high-confidence regional evidence. Loci are ordered by decreasing PPA4. (b) Loci with shared genetic effects. Horizontal bars show the regional posterior probability for Model⍰3 (PPA3), corresponding to loci most consistent with a shared causal variant influencing both AD and T2D. Colour intensity reflects the regional logBF3, using the same scale as panel (a) for comparability. Loci are ordered by decreasing PPA3. (c) Representative SNPs within Model⍰3-supported regions. Each bar represents a SNP selected post hoc from genomic regions with high regional support for Model⍰3, prioritised based on high conditional SNP-level posterior probability (PPA3⍰≥ ⍰0.9) and contribution to the regional Bayes factor. These SNPs are shown as descriptive representatives of regional shared-variant signals rather than independent causal assignments. Colour indicates SNP-level logBF3, and bars are spaced to improve visibility at high PPA3 values. SNPs may reside in loci that do not reach conventional genome-wide significance thresholds. Across all panels, posterior probabilities (PPA3, PPA4) quantify the probability of the corresponding regional causal configuration, while Bayes factors (logBF3, logBF4) indicate the strength of relative evidence between competing models. LogBF values are interpreted using established guidelines: values ≥⍰20 indicate very strong evidence, values of approximately 8–20 indicate strong support, and smaller values are interpreted with caution, consistent with Kass and Raftery’s Bayes factor scale. Abbreviations: AD, Alzheimer’s disease; T2D, type⍰2 diabetes; SNP, single-nucleotide polymorphism; PPA, posterior probability of association; logBF, log Bayes factor; BP, base-pair position; CHR, chromosome; GWAS, genome-wide association study; GWS, genome-wide significant.

In contrast, a smaller subset of regions showed strong support for Model 3, consistent with shared causal variants influencing both AD and T2D. Based on regional posterior probabilities for Mode⍰3, we identified seven genomic regions across chromosomes 6, 11, 16, 17, 19, and 20 exhibiting high (PPA3 > 0.9) or moderate-to-high (0.5 < PPA3 < 0.9) posterior probability support (Figure 5; Supplementary Table 16). These regions were characterised by high logBF3 and PPA3 values, indicating strong relative evidence in favour of the shared-variant configuration compared with alternative models. Notably, a locus on chromosome⍰19 showed overwhelming regional support for Mode⍰3 (logBF3 = 1,397). Across these regions, association signals were strong for both traits (maximum Z⍰= ⍰52.5 for AD and 9.7 for T2D), supporting the inference that shared-variant signals are driven by robust evidence in each disease rather than being artefacts of asymmetric association strength. These findings provide evidence for a subset of genomic regions in which AD and T2D risk is influenced by the same underlying genetic signal, consistent with shared genetic effects at the locus level and potentially overlapping molecular pathways.

GWAS-PW reports Bayes factors and posterior probabilities for every SNP; hence, we selected SNPs post hoc within regions showing high regional posterior support for Mode⍰3, applying a SNP-level threshold of PPA3⍰> ⍰0.90. These SNPs were used as descriptive representatives of the regional shared-variant signal, reflecting their conditional contribution to the regional Bayes factors. Accordingly, we identified 35 SNPs across 18 chromosomes in Mode⍰3-supported regions, serving as high-confidence representatives of the regional shared-variant signal (Figure 5; Supplementary Table 17). Several loci exhibited exceptionally strong regional evidence for shared effects, with variants contributing substantially to the regional Bayes factors. Notably, rs429358 at the *APOE* locus resided within a region showing maximal posterior support for Mode⍰3 and extremely large regional logBF3, alongside strong association with AD (Z⍰= ⍰52.5) and robust association with T2D (Z⍰= ⍰8.7). Additional regions with strong Mode⍰3 support included those tagged by rs7756992 (chromosome⍰6), rs2237895 and rs10830963 (chromosome⍰11), rs4663105 (chromosome⍰2), and rs76895963 (chromosome⍰12), each characterised by large regional Bayes factors and strong evidence in at least one of the two traits. Across regions, the relative contribution of AD and T2D association signals varied: some shared-signal regions were driven primarily by AD associations (e.g. the *APOE* locus), others by T2D, and several by substantial evidence in both diseases (Supplementary Table 17). These results identify a robust set of genomic regions with strong support for shared genetic effects on AD and T2D, consistent with partial overlap in their genetic architecture.

### GWS genes shared by AD and T2D in gene-based cross-trait meta-analysis

Based on a Bonferroni-corrected threshold for selecting GWS (P_gene_ < 2.64 × 10^−6^, accounting for the total number of genes), we identified four genes that were independently significant in both disorders and exhibited a strengthened association following p-value aggregation (Supplementary Table 18). Three of these genes, *APOC1, APOE*, and *TOMM40*, clustered within the *APOE* locus, while *ACE* on chromosome 17 also demonstrated consistent evidence of association across traits. For all shared genes, the combined Stouffer’s Z-scores exceeded those observed in the individual GWAS, yielding highly significant combined p-values (as low as P ≤ 4.2 × 10^-28^, Supplementary Table 18). We observed multiple entries for *ACE*, which may reflect distinct gene definitions or transcript models used in the gene-based association analysis (Supplementary Table 18). Gene-based analyses here suggest shared signals between AD and T2D at several loci, most notably within the *APOE* region. However, locus-resolved analyses using LAVA showed that this shared gene-level signal does not imply concordant effects. Instead, local genetic correlations at *APOE* were strongly negative, demonstrating that although the same region contributes to both traits, the underlying genetic effects likely act in opposite directions.

### Shared genes reaching genome-wide significance for AD and T2D in gene-based analysis

We extended our gene-based cross-trait analysis using SCP to identify genes shared by AD and T2D in three categories: (1) genes GWS for AD (P_gene-AD_ < 2.64 × 10^-6^) and at least showing evidence of association with T2D at the nominal level (P_gene-T2D_ < 0.05); (2) genes GWS for T2D (P_gene-T2D_ < 2.64 × 10^-6^) and at least nominally associated with AD (P_gene-AD_ < 0.05); and (3) genes nominal in both individual gene-based tests (2.64 × 10^-6^ < P_gene_ < 0.05 for each) but attaining GWS in the combined (SCP) test (P_SCP_ < 2.64 × 10^-6^), representing potential candidate novel shared genes. Using these criteria, we observed 10 genes in the first category (Table 1), 51 genes in the second category (Supplementary Table 19), and 45 genes in the third category (Supplementary Table 20). The shared genes include multiple *APOE*-region genes (*APOC1, APOE*, and *TOMM40*), *ACE* (two transcript models reported), *C6orf10, PLEKHA1, INO80E, DOC2A, PPP4C, YPEL3, FIBP, FOSL1, ZNF668, CAMTA2, and C16orf92*, among others.

**Table 1:** AD Genome-wide significant genes associated with T2D.

| GENE | CHR | AD<sup>a</sup> |  | T2D<sup>b</sup> |  | Stouffer's Method (equal weights) |  |  |  | Stouffer's P < the individual GWAS p |
| --- | --- | --- | --- | --- | --- | --- | --- | --- | --- | --- |
|  |  | Sample size | P-value | Sample size | P-value | Z-score AD | Z-score T2D | Stouffer's Z-score | P-value |  |
| RP11-196G11.1 | 16: 31094760 - 31106277 | 356072 | $2.49 \times 10^{-6}$ | 898130 | $9.63 \times 10^{-3}$ | -4.57 | -2.34 | -4.88 | $5.21 \times 10^{-7}$ | Yes |
| VKORC1 | 16: 31102163 - 31107301 | 378741 | $1.32 \times 10^{-6}$ | 898130 | $2.22 \times 10^{-3}$ | -4.70 | -2.84 | -5.33 | $4.84 \times 10^{-8}$ | Yes |
| BCKDK | 16: 31117428 - 31124110 | 408235 | $5.29 \times 10^{-8}$ | 898130 | $1.15 \times 10^{-3}$ | -5.32 | -3.05 | -5.91 | $1.66 \times 10^{-9}$ | Yes |
| KAT8 | 16: 31127075 - 31142714 | 409306 | $1.91 \times 10^{-7}$ | 898130 | $3.02 \times 10^{-4}$ | -5.08 | -3.43 | -6.02 | $8.97 \times 10^{-10}$ | Yes |
| ACE | 17: 61554422 - 61599205 | 376655 | $9.10 \times 10^{-7}$ | 898130 | $1.50 \times 10^{-6}$ | -4.77 | -4.67 | -6.68 | $1.21 \times 10^{-11}$ | Yes |
| ACE | 17: 61562184 - 61599209 | 381061 | $2.17 \times 10^{-6}$ | 898130 | $1.75 \times 10^{-6}$ | -4.59 | -4.64 | -6.53 | $3.31 \times 10^{-11}$ | Yes |
| PVRL2 | 19: 45349432 - 45392485 | 386816 | $1.01 \times 10^{-12}$ | 898130 | $1.27 \times 10^{-5}$ | -7.03 | -4.21 | -7.95 | $9.33 \times 10^{-16}$ | Yes |
| TOMM40 | 19: 45393826 - 45406946 | 385101 | $7.20 \times 10^{-14}$ | 898130 | $1.15 \times 10^{-10}$ | -7.39 | -6.34 | -9.71 | $1.37 \times 10^{-22}$ | Yes |
| APOE | 19: 45409011 - 45412650 | 409171 | $2.56 \times 10^{-14}$ | 898130 | $1.43 \times 10^{-11}$ | -7.53 | -6.65 | -10.03 | $5.71 \times 10^{-24}$ | Yes |
| APOC1 | 19: 45417504 - 45422606 | 264790 | $3.11 \times 10^{-15}$ | 898130 | $9.60 \times 10^{-15}$ | -7.80 | -7.66 | -10.93 | $4.21 \times 10^{-28}$ | Yes |
| CD33 | 19: 51728320 - 51747115 | 390954 | $1.73 \times 10^{-6}$ | 898130 | $1.70 \times 10^{-2}$ | -4.64 | -2.12 | -4.78 | $8.72 \times 10^{-7}$ | Yes |

AD<sup>a</sup>: Alzheimer's disease (Jansen et al), GWAS: genome-wide association studies, T2D<sup>b</sup>: type 2 diabetes (Mahajan et al), GWS: genome-wide significance. Stouffer's Z-scores were calculated using equal weights across studies to avoid dominance of the larger T2D GWAS sample size. Multiple entries for ACE reflect distinct gene definitions or transcript models used in the gene-based association analysis. Genome-wide gene-based significance: $P_{\text{gene}} < 2.64 \times 10^{-6}$ (Bonferroni correction for 18,965 tested genes). Gene-based Z-score signs summarise aggregate evidence and should not be interpreted as definitive per-variant directional concordance; locus-level signed genetic covariance is assessed with LAVA.

### Genes prioritised for expression-driven links between Alzheimer’s disease and type 2 diabetes

We performed SMR to test whether genetically predicted gene expression influences the risk of AD and T2D. Results reported here were restricted to associations surviving FDR correction, supported by at least 10 cis eQTL instruments per gene, and demonstrating no evidence of heterogeneity (all pHEIDI > 0.01; nHEIDI ≥ 10) (Figure 6, Supplementary Table 21). Across eQTL resources (GTEx, eQTLGen, BrainMeta), 17 genes (12 unique genes) exhibited significant SMR associations with both traits, including *FIBP, SNX32, ACE, PLEKHA1, YPEL3, PPP4C, KAT8, INO80E, BCKDK, ZNF668, HSD3B7*, and *GALNT10*. All these genes (except *HSD3B7* and *SNX32*) are supported in gene-based analysis findings (Supplementary Tables 19 and 20). Several genes were supported across multiple tissues or independent eQTL platforms, most notably *KAT8, INO80E, BCKDK*, and *HSD3B7*, strengthening evidence for robust shared regulatory effects (Figure 6).

**Figure 6:**
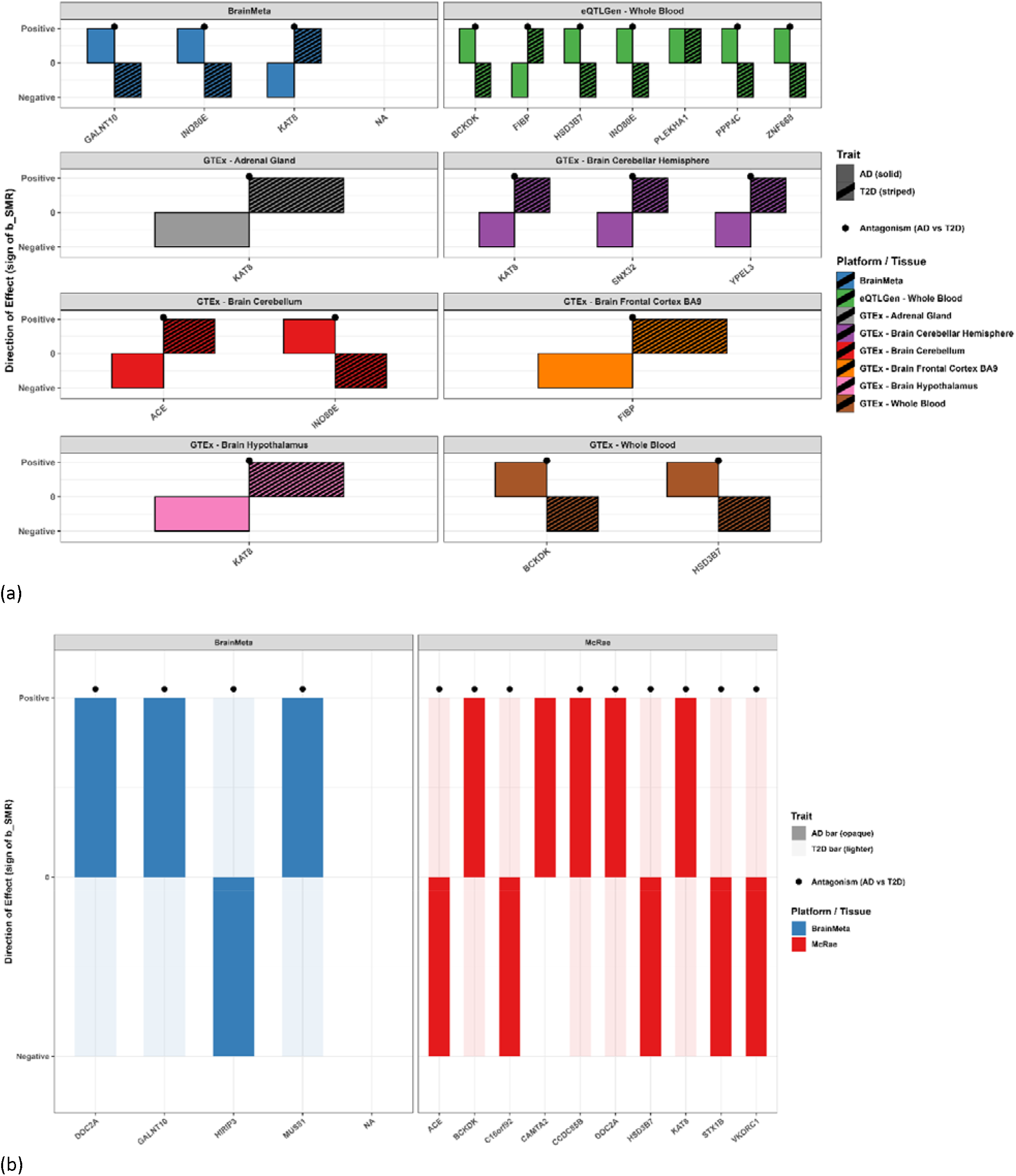
Directional AD and T2D effects for eQTL and mQTL genes showing concordant and putatively antagonistic patterns. Panel a: eQTL directional effects (AD vs T2D). This panel summarises the direction of SMR effects for each gene–platform pair across Alzheimer’s disease (AD) and Type 2 diabetes (T2D). Bars represent the sign of the SMR effect: Positive = increased expression associated with higher disease risk, while Negative = decreased expression associated with higher disease risk. AD = opaque bars, T2D = lighter bars; both shown side-by-side per gene. Coloured panels correspond to different eQTL platforms/tissues, allowing cross-tissue comparison. A black dot above a gene indicates antagonism, meaning the AD and T2D effects point in opposite directions (e.g., AD positive, T2D negative), highlighting genes where expression has divergent implications for the two diseases. Panel b: mQTL directional effects (AD vs T2D). This panel shows the corresponding methylation-QTL (mQTL) directional effects for the same AD and T2D traits. As in Panel a, bars show positive or negative SMR effect directions, with opaque = AD and lighter = T2D. The two mQTL data sources (BrainMeta and McRae) are displayed in separate facets for clarity. Antagonistic effects (black dot above the bar) again mark genes where AD and T2D methylation effects move in opposite directions. Comparison with Panel a helps identify whether expression-level antagonism is mirrored (or not) at the methylation-regulation level, which may indicate multi-layer regulatory divergence.

The direction of SMR effects revealed a predominance of discordance, consistent with antagonistic pleiotropy between AD and T2D. Increased expression of *INO80E, BCKDK, ZNF668, HSD3B7, PPP4C*, and *GALNT10* was associated with higher AD risk but lower T2D risk, whereas *FIBP, SNX32, ACE, YPEL3*, and *KAT8* showed the opposite pattern, with lower AD risk but higher T2D risk. In contrast, *PLEKHA1* (also identified in gene-based analysis) was the only gene demonstrating concordant positive associations with both AD and T2D, suggesting a shared regulatory effect operating in the same direction across AD and T2D. For several genes, the same top cis eQTL variant instrumented expression for both traits (e.g., FIBP, *SNX32, ACE, INO80E, BCKDK*, and *HSD3B7*), yet with opposing effect directions, supporting shared causal regulatory variants exerting divergent phenotypic consequences rather than trait-specific linkage. This finding notably strengthens the evidence for antagonism where the same top cis-eQTL variant instrumented expression for both traits yet produced opposing effect directions, consistent with shared causal regulatory variation driving divergent phenotypic outcomes. Together, these results identify a focused set of cis-regulated genes whose expression levels contribute to both AD and T2D susceptibility, dominated by putatively antagonistic but also including a concordant regulatory effect, supporting complex shared molecular architectures potentially linking neurodegeneration and metabolic dysfunction.

### Methylation-driven shared associations between AD and T2D

Integrative SMR analyses of brain- and blood-derived mQTLs identified a focused set of 13 unique genes, represented by 14 CpG sites, exhibiting shared methylation-driven associations with both AD and T2D, all supported by non-heterogeneous HEIDI results (pHEIDI⍰> ⍰0.01; nHEIDI⍰≥ ⍰10) [Supplementary Table 22]. Most loci displayed opposite directions of effect between the two traits, suggesting predominantly putative antagonistic pleiotropy. Notably, increased methylation at *GALNT10, MUS81, CCDC85B, DOC2A, BCKDK*, and *KAT8* was associated with higher AD risk but lower T2D risk, whereas *HIRIP3, C16orf92, HSD3B7, STX1B, VKORC1*, and *ACE* showed the opposite pattern. *CAMTA2* was the only gene demonstrating concordant positive associations with both AD and T2D. For *DOC2A*, consistency across multiple CpG probes strengthened its prioritisation. Although several CpGs within the *APOE* locus (*APOE, APOC1, NECTIN2*, and *DMPK*) were significantly associated with both disorders, all failed the HEIDI test, suggesting likely linkage rather than a shared causal methylation signal (Supplementary Table 23). These genes were excluded from our prioritised results, yet they exhibited opposite effects on AD and T2D, consistent with findings from other methods, which may further reinforce the biological plausibility of antagonistic relationships in this region. These findings highlight a set of methylation-regulated genes with divergent biological effects on AD and T2D risk, underscoring the complex molecular architecture potentially linking the two disorders.

### Summary of overlapping eQTL and mQTL genes/loci and alignment with gene-based findings

This study identifies robust cross-omics evidence that AD and T2D share genetic regulatory architecture at both gene and locus levels, with multiple genes prioritised across independent eQTL- and mQTL-SMR analyses. The genes identified in eQTL and mQTL were largely supported in the gene-based and Stouffer’s p-value aggregation analyses. Specifically, five genes (*GALNT10, HSD3B7, BCKDK, KAT8*, and *ACE*) were consistently supported across expression and methylation layers, and many shared signals concentrated within the 16p11.2 region, highlighting a regulatory hotspot implicated in both disorders. However, the shared architecture is predominantly characterised by putatively antagonistic pleiotropy, where the same genetically regulated expression or methylation changes are associated with increased risk of one trait but decreased risk of the other. The observation that identical top cis-QTL instruments can underlie both traits while yielding opposite SMR directions further supports shared putatively causal regulatory variants with divergent phenotypic consequences rather than trait-specific linkage. Only a limited number of signals showed concordant directions (e.g., *PLEKHA1* in *eQTL-SMR* and *CAMTA2* in mQTL-SMR), indicating that while some mechanisms may increase susceptibility to both conditions, most shared regulatory effects appear to involve trade-offs between neurodegenerative and metabolic risk pathways. Overall, these findings support a model in which the AD-T2D relationship arises from shared loci operating through complex, multi-layer regulation that is more often directionally opposed than aligned.

## Discussion

We applied multi-omic analyses integrating genomic, transcriptomic, and epigenomic data to investigate the relationship between AD and T2D. We found a modest but statistically significant positive genome-wide correlation, replicated across multiple datasets and persisting after exclusion of the *APOE* region. SNP-effect concordance analyses similarly supported robust and reproducible shared polygenicity across traits. However, higher-resolution analyses revealed pronounced locus-specific heterogeneity in both magnitude and direction of sharing. The *APOE* region showed consistently strong negative local genetic correlations across all GWAS pairings, while the extended MHC exhibited a mixture of positive and negative sub-regional effects. These opposing signals potentially explain why genome-wide correlation estimates can appear attenuated or near zero despite underlying biological overlap, a pattern consistent with prior reports of discordant allelic effects at *APOE* and *HLA*^33,63-65^. Our direction-aware local correlation analyses quantify this phenomenon, demonstrating that robust negative local covariance can coexist alongside a modest positive global correlation. These findings indicate that locus-level sharing does not necessarily reflect concordant biology and that genome-wide concordance largely arises from many small aligned polygenic effects, while a small number of high-impact loci contribute strong antagonistic covariance.

Against this backdrop, we revisited previous global genetic correlation findings^32,33^. For example, in apparent contrast to our results, Hardy and colleagues reported no significant global AD-T2D genetic correlation and concluded that shared susceptibility was limited^33^. This difference is unlikely to reflect power: well-powered GWAS were used in both studies, but rather the sensitivity of LDSC to mixtures of positive and negative local effects and to implementation-level decisions. In architectures where strong putative antagonistic pleiotropy at high-leverage loci (such as *APOE* and segments of the MHC) coexists with dispersed polygenic concordance elsewhere, modest changes in SNP inclusion, LD reference, baseline model, genomic-control correction, handling of *APOE* or MHC regions, or intercept constraints can shift estimates from modestly positive to near zero. Thus, our findings refine rather than contradict earlier work: global AD–T2D genetic correlation appears modest but statistically detectable in our analyses, while the underlying architecture is directionally heterogeneous.

Cross-trait GWAS meta-analysis^47^ and Bayesian colocalisation^53^ refined our understanding of shared loci. The heterogeneity-aware RE2 model identified genome-wide significant signals at known loci (*APOE*, HLA) and several other regions. However, posterior diagnostics (BE P-values and m-values)^48^ indicated that many of these were trait-dominant, with variants contributing primarily to one disorder. Bayesian colocalisation broadly supported this pattern: most windows favoured Mode⍰4 (distinct causal variants), with only a minority of regions showing moderate-to-high support for Mode⍰3 (shared causal variant). Even Mode⍰3 regions do not necessarily imply concordant effects, as shared causal variants may still act in opposite directions. Overall, these results show that apparent cross-trait loci often reflect locus-level convergence rather than single-variant pleiotropy. Our findings advance earlier gene-set and locus-enrichment observations^32,34,36^ by demonstrating that many previously interpreted ‘shared’ signals are better explained by variant-level heterogeneity. The extended MHC region illustrates this complexity. The RE2 meta-analysis yielded highly significant cross-trait signals consistent with immune involvement in both disorders^26,28,32^. However, variant-level posterior diagnostics showed that most contributing SNPs were T2D-dominant. Colocalisation again favoured distinct causal variants, and direction-aware local correlation revealed both positive and negative local associations, indicating a mixture of shared, trait-specific, and putatively antagonistic effects. These results refine interpretations equating immune enrichment with a unified immune mechanism and underscore the need to account for effect-direction and heterogeneity when inferring shared biology.

Despite the heterogeneity observed across most loci, a small subset showed consistent cross-trait support across RE2 meta-analysis, BE P-values, and m-values, consistent with robust pleiotropy rather than trait-dominant effects. Four loci: rs34016387, rs35366052, rs7131432, and rs150775861, met this criterion. Among these, rs7131432 tags SORL1, previously implicated in AD^29,60,66^ but not T2D, while rs150775861 lies near C16orf74, a region with sub-threshold T2D associations. The remaining two loci have no prior reported associations with either disorder (to our knowledge), suggesting potentially novel cross-trait signals. These results refine earlier enrichment-based interpretations of overlapping AD–T2D risk variants^13,67^ by showing that only a small minority of loci demonstrate statistically robust cross-trait involvement once heterogeneity-aware meta-analysis and posterior diagnostics are applied.

Gene-based and p-value aggregation analyses provided complementary evidence for cross-trait convergence, identifying genes such as *VKORC1, ACE, BCKDK, KAT8, PVRL2, TOMM40, APOE, CD33*, and *APOC1* with strengthened combined signals. Shared genome-wide significant gene-level associations were observed at the *APOE/APOC1/TOMM40* cluster and *ACE*, consistent with previously implicated lipid, immune, and vascular pathways^13,32^. However, gene-level aggregation reflects shared association burden rather than effect direction: strong negative local correlation at *APOE* illustrates how antagonistic pleiotropy can coexist with strengthened gene-based signals. Similarly, although convergence at *ACE* is biologically plausible given its vascular and endocrine roles^68^, much of the underlying architecture does not support concordant effects in AD-T2D association^33^. Together, current results show that apparent gene-level sharing frequently arises from distinct variant-level mechanisms and directionally divergent effects.

The SMR analyses identified several genes whose expression or methylation contributed to both disorders, but most exhibited opposing regulatory effects, reinforcing putative antagonistic pleiotropy. Only two genes, *PLEKHA1* (eQTL) and *CAMTA2* (mQTL), showed concordant positive effects on AD and T2D, indicating that the same-direction of regulatory pleiotropy is rare but present. Many SMR-highlighted putatively causal genes overlapped with those identified in the gene-based analyses, including *ACE, KAT8*, and several genes located at 16p11.2 (e.g., *VKORC1*), suggesting convergence on shared pathways even when causal variants may differ or act in divergent directions. These findings support a model in which AD and T2D converge on shared genes through fragmented regulatory architecture rather than a uniformly shared causal mechanism. We also identify a cross-omic regulatory hotspot at 16p11.2, supported across expression and methylation layers. These observations align with prior reports of opposite-direction effects at *APOE* and *HLA* ^33,45,46^, consistent with antagonistic pleiotropy. Our study extends these earlier findings by integrating cross-omic evidence to show that regulatory convergence frequently coexists with directional discordance, delineating a broader multi-layer architecture, including putatively novel cross-omic genes and the 16p11.2 hotspot, beyond what SNP-level analyses alone can reveal.

Lastly, MR analyses provided no evidence for a direct causal effect of AD liability on T2D, nor of T2D on AD. Combined with our cross-omic results, this finding suggests that the epidemiological association between the two disorders does not reflect a single genetically mediated causal pathway. Instead, modest shared polygenicity, locus-specific heterogeneity, locus-level convergence, and small pockets of concordant pleiotropy contribute to a complex cross-trait architecture. Strong antagonistic effects at loci such as *APOE* and segments of the MHC further indicate that cross-trait overlap often arises from pathways acting in opposite directions. Overlapping biological contexts, particularly vascular, metabolic, and immune pathways, may also generate shared vulnerability through mechanisms not captured by MR, including metabolic dysfunction, vascular injury, inflammation, lifestyle exposures, or medication effects. We note, as an additional non-genetic explanation, that survival (competing-risk) bias may contribute to the apparent antagonistic effects observed at high-impact loci. Because T2D and related metabolic conditions shorten life expectancy, individuals at high metabolic risk may be under-represented among older AD cases, making metabolic risk alleles appear protective in cross-sectional or age-unadjusted analyses. Future work using age-stratified or incident-case designs, survival-GWAS, or competing-risk models will be important for quantifying the contribution of survivorship to these locus-level patterns

### Strengths and limitations

A major strength of this study is its comprehensive multi-omic design, integrating genome-wide association data with transcriptional and epigenomic evidence to characterise the AD–T2D relationship across biological layers. By combining genome-wide LDSC and SECA, direction-aware local genetic correlation, heterogeneity-aware cross-trait meta-analysis, Bayesian colocalisation, gene-based aggregation, and eQTL/mQTL-informed SMR, we provide detailed evaluations of cross-trait architecture between AD and T2D. This integrative framework enables discrimination between concordant pleiotropy, putative antagonistic pleiotropy, trait-specific effects, and locus-level convergence, patterns that cannot be resolved by a single method or genome-wide focused approaches. Another strength is our conservative cross-omic matching strategy, which requires overlap within the same QTL resource and regulatory feature, thereby prioritising high-confidence shared regulatory signals. Although this approach may under-detect broader tissue-shared effects, it strengthens confidence in our conclusions. The identification of cross-omic candidates and regulatory hotspots such as 16p11.2 further provides mechanistically meaningful targets for functional follow-up.

Several limitations should also be noted. First, despite improved resolution from colocalisation and SMR, most shared loci favoured distinct causal variants, and regulatory inference is constrained by the tissue composition of available QTL datasets, which may not capture all disease-relevant cell types or contexts. Second, Mendelian randomisation captures genetically proxied lifetime effects and may not adequately resolve time-dependent, tissue-specific, or environmentally contingent mechanisms, such that context-specific causal pathways may remain undetected. Third, the reliance on European-ancestry datasets limits generalisability to other ancestries. Finally, loci such as *APOE* and regions within the extended HLA exhibit complex LD and likely harbour multiple independent signals. Our colocalisation analyses adopt single-signal assumptions that may be violated in these regions; conditional analyses and multi-ancestry fine-mapping will may be needed to distinguish multiple causal variants from genuinely shared effects. Accordingly, we interpret locus-level directional discordance cautiously and view these regions as priorities for targeted follow-up rather than definitive evidence of biological antagonism.

### Conclusion, implications for the observed epidemiological association and the broader field

This study shows that AD and T2D share a complex, multilayered genetic architecture in which modest genome-wide correlation coexists with pronounced locus-specific heterogeneity. Although we observed a reproducible positive genome-wide genetic correlation and widespread concordant SNP-effect directions, this diffuse polygenic alignment masks substantial local variation: several loci, most notably *APOE* and regions within the MHC, exhibit strong inverse effects consistent with antagonistic pleiotropy, while only a minority of regions show concordant associations. Cross-trait meta-analysis further indicates that most genome-wide significant signals are trait-specific, and Bayesian colocalisation demonstrates that most overlapping loci are driven by distinct causal variants. Consistently, eQTL- and mQTL-based SMR analyses reveal that most shared regulatory signals act in opposite directions, with only a small set of concordant cross-omic candidates, such as *PLEKHA1* and *CAMTA2*.

In relation to the long-standing epidemiological association between AD and T2D, the scarcity of concordant genetic effects, together with the lack of evidence for bidirectional genetically proxied causal effects in MR, suggests that their co-occurrence is unlikely to reflect a single shared genetic susceptibility. Instead, the overlap appears to arise from broad polygenic influences, shared regulatory architecture, and convergence on partially overlapping biological pathways. Although we observe a modest genome-wide polygenic correlation, the overall architecture is dominated by marked locus-specific heterogeneity and frequent directional discordance, including putative antagonistic pleiotropy. This position indicates that cross-trait overlap reflects a mixture of concordant and opposing effects across loci rather than a unified inherited mechanism or a simple shared-risk model.

More broadly, our integrative findings refine how cross-trait genetic overlap between AD and T2D should be interpreted. Modest genome-wide correlation can coexist with strong local antagonistic pleiotropy, and gene- or pathway-level convergence does not imply shared causal variants or concordant biological effects. These insights have important translational implications: gene-level sharing should not be assumed to represent shared therapeutic targets, particularly at loci exhibiting antagonistic regulation, where disease-specific interventions may be required. This is particularly relevant for efforts to repurpose T2D therapies for AD, or AD-related interventions for T2D. Conversely, the few loci showing consistent concordant pleiotropy, such as *PLEKHA1* and *CAMTA2*, and the variant-level findings, represent promising candidates for functional follow-up. Overall, AD and T2D appear genetically and mechanistically distinct at most loci. The combination of modest genome-wide overlap, diffuse polygenic concordance, strong local antagonistic pleiotropy, and a lack of evidence for causal effects in MR provides a coherent explanation for the observed patterns of genetic overlap between the disorders, despite limited evidence for shared genetic causality. These results highlight the importance of locus-aware, direction-sensitive, and multi-omic approaches when interpreting cross-trait relationships in complex disease biology.

## Methods

### Data sources

GWAS summary statistics were sourced from large, publicly available datasets generated by international research collaborations. For AD, we used summary data from a comprehensive GWAS meta-analysis, including 71,880 individuals with AD and 383,378 unaffected controls^26^. This resource combined genetic data from both clinically ascertained AD cases and individuals classified as AD based on proxy phenotypes. The clinically diagnosed AD subset comprised several independent case-control studies. It included approximately 24,000 individuals with a confirmed AD diagnosis and more than 55,000 control participants, yielding a total sample of 79,145 individuals. Case definitions were based on clinician assessment or documented medical diagnoses, while controls were characterised as lacking a diagnosis of AD or dementia at the time of evaluation^26^.

Proxy-defined AD cases were largely drawn from the UK Biobank and identified through reported parental history of AD, with additional information on parental age at diagnosis or death incorporated into the phenotype definition. Previous analyses have demonstrated a high degree of genetic similarity between clinically diagnosed AD and AD-by-proxy phenotypes (genetic correlation r = 0.81)^26,27^, providing a strong rationale for their joint analysis in genetic studies. For validation, we additionally analysed GWAS summary statistics restricted to cohorts with clinically confirmed AD diagnoses. These data were obtained from a large meta-analysis aggregating results from four major collaborative efforts: the European AD Initiative (EADI), the Cohorts for Heart and Aging Research in Genomic Epidemiology (CHARGE), the AD Genetics Consortium (ADGC), and the Genetic and Environmental Risk in AD (GERAD) consortium. The combined dataset comprised participants of European ancestry and included 17,008 individuals diagnosed with AD and 37,154 control subjects^29^. Detailed descriptions of cohort recruitment, phenotyping, and quality control procedures have been reported elsewhere^29^.

Summary statistics for T2D were obtained from multiple high-quality GWAS resources, including large international consortia and population-based biobanks. The primary T2D dataset was drawn from the DIAGRAM consortium and reported by Mahajan *et al*., comprising 74,124 cases and 824,006 controls^28^. Additional T2D GWAS data were sourced from independent studies and resources, including Xue *et al*.,^30^ and phenotype-defined data from the UK Biobank, consisting of white British participants. To further assess the robustness and reproducibility of our findings, we incorporated an independent T2D GWAS from the FinnGen consortium (release R10)^58^. FinnGen is a large-scale Finnish population study that integrates genetic data with nationwide health registry information from over 412,000 participants, enabling the investigation of genetic susceptibility across a broad range of complex diseases.

We obtained cis-expression QTL summary statistics from GTEx v8 (multiple tissues), eQTLGen (whole blood), and BrainMeta (brain-derived datasets), providing complementary coverage across central nervous system and peripheral regulatory contexts. These resources include large-scale, well-curated transcriptomic datasets with extensive sample sizes and standardised quality control, enabling robust identification of regulatory variants influencing gene expression across diverse tissues. For methylation QTL analyses, we used summary statistics from BrainMeta and the McRae blood mQTL resource, which provide genome-wide maps of genetic influences on DNA methylation across brain and blood tissues. These datasets offer high-resolution coverage of CpG sites and well-powered cis-mQTL associations, facilitating integrative regulatory analyses across transcriptional and epigenomic layers. Only cis-QTL associations (within ±1 Mb of gene transcription start sites or CpG sites) were considered. These resources were selected to maximise regulatory coverage across disease-relevant tissues while enabling cross-platform validation of shared regulatory effects.

### Genome-wide genetic correlation analysis

We applied LDSC^31^ to estimate genome-wide genetic correlations between AD and T2D. LDSC enables separation of polygenic signal from confounding due to population stratification, cryptic relatedness, or model misspecification. GWAS summary statistics were pre-processed according to LDSC recommendations, including removal of duplicate variants, ambiguous SNPs, non-SNP variants, and poorly imputed markers. Cross-trait (bivariate) LDSC was then used to estimate genetic correlations, with LD scores derived from the 1000 Genomes Project reference panel for individuals of European ancestry. Analyses were conducted with the genetic covariance intercept constrained, as unconstrained analyses indicated no evidence of significant sample overlap. Genetic correlation analysis was performed using the main AD and T2D GWAS datasets, and results were subsequently evaluated for consistency using four validation datasets for T2D. For validation on the side of AD, we used the clinically diagnosed GWAS. In each set, five separate AD–T2D genetic correlation tests were conducted. Accordingly, statistical significance was defined as P < 0.05, with a Bonferroni-corrected threshold of P < 0.01 (0.05/5) applied to account for multiple testing. Genetic correlation estimates range from −1 to 1, with positive values suggesting shared genetic effects that potentially increase risk for both traits, and negative values indicating opposing genetic effects. Given the well-established and potentially disproportionate influence of the *APOE* locus on AD risk, we conducted sensitivity analyses both including and excluding the *APOE* region to examine its effect on the observed genome-wide genetic correlations.

### Effect concordance and genetic overlap analysis

To further assess the genetic relationship between AD and T2D across multiple cohorts, we performed analyses using the SECA framework^51^. SECA is designed to assess shared genetic architecture by evaluating both concordance in SNP effect directions and evidence of SNP-level genetic overlap (pleiotropy), with the additional advantage of enabling bidirectional analyses^51^. We restricted the current analysis to only two AD GWAS (one comprising clinically diagnosed and proxy AD cases^26^, for the main investigation, and the other consisting of only clinically diagnosed cases^29^, for validation testing. Each of these AD GWAS data was assessed against the T2D GWAS from two studies (Mahajan et al^28^, for the main investigation, and Xue et al^30^, for validation).

Before analysis, GWAS summary statistics underwent quality control, including removal of duplicate and non-rsID variants and harmonisation of effect alleles to ensure consistency across datasets. Independent SNPs were identified using LD clumping implemented in PLINK v1.9^69^, applying an r^2^ threshold of <0.1 within a 1 Mb window, followed by a broader 10 Mb window to capture longer-range LD structure. SNPs were stratified into subsets according to their association P-values in the reference dataset, and concordance across subsets was assessed using Fisher’s exact tests^51^. Statistical significance was evaluated using permutation-based correction with 1,000 replicates. Bidirectional analyses were conducted by alternately designating AD and T2D as the reference dataset to account for potential asymmetry in genetic effects. To account for the potentially outsized influence of the *APOE* locus on AD risk, we performed SECA analyses with and without the *APOE* region.

Beyond concordance in effect direction, we further examined SNP-level genetic overlap between AD and T2D using SECA to assess evidence of potential pleiotropy^51^, defined as genetic variants influencing both traits. Specifically, a binomial test was applied to determine whether the number of SNPs jointly associated with both traits exceeded that expected by chance (P_binomial_ < 0.05). This analysis was conducted across three P-value threshold combinations (P1: dataset 1; P2: dataset 2): (i) P1 < 0.05 and P2 < 0.05 (nominal significance), (ii) P1 < 1 × 10^-5^ and P2 < 0.05 (suggestive genome-wide significance in dataset 1), and (iii) P1 < 5 × 10^-8^ and P2 < 0.05 (genome-wide significance in dataset 1). The analyses provided complementary evidence for shared genetic effects and pleiotropy between AD and T2D across independent datasets.

### Regional genetic correlation analysis

To identify specific genomic regions that contribute disproportionately to the shared genetic basis between AD and T2D, we performed local genetic correlation analyses using the LAVA framework^52^. Unlike genome-wide approaches such as LDSC, which provide an average estimate of genetic correlation across the genome^31^, LAVA enables high-resolution, locus-by-locus assessment of shared genetic influences^52^. In the present study, we conducted analyses using the semi-independent LD blocks defined in the original LAVA publication, with LD estimated from the 1000 Genomes Project Phase 3 European reference panel, including variants with minor allele frequency >0.5%. Following allele harmonisation and standard quality control, LAVA first estimated local SNP-based heritability for each trait within each LD block. Only loci exhibiting sufficient univariate signal were subsequently tested for bivariate local genetic correlation. Consistent with recommendations from the LAVA developers and prior studies, we applied a univariate screening threshold of *P* < 0.05 in our analyses. This threshold maintains appropriate control of type I error while allowing a broader set of loci to be interrogated for region-specific shared genetic effects.

We performed local genetic correlation analyses across multiple GWAS dataset pairings (AD Jansen vs. T2D Mahajan and T2D Xue; AD Lambert vs. T2D Mahajan and T2D Xue), enabling locus-specific evaluation of shared genetic architecture and cross-dataset validation. Results were interpreted based on the magnitude and direction of the local genetic correlation coefficient, statistical significance, and confidence interval estimates. We considered loci for which the confidence intervals (CIs) included a value of 1 as consistent with near-complete sharing of genetic effects between traits at that region. Nominal significance was defined as *P* < 0.05, with Bonferroni correction applied to account for multiple testing across all loci evaluated within each dataset pairing.

### Bidirectional Mendelian randomisation

We performed bidirectional two-sample MR analyses^54,55^ to assess potential causal relationships between AD and T2D. MR analyses are predicated on three key assumptions: that genetic instruments are robustly associated with the exposure, affect the outcome solely through the exposure, and are not correlated with confounders^54^. We applied multiple complementary MR methods to estimate causal effects in both directions, with careful instrument selection and sensitivity analyses to mitigate bias from horizontal pleiotropy and heterogeneity. Analyses were restricted to individuals of European ancestry and conducted in accordance with STROBE-MR reporting guidelines^54,70,71^. We selected genetic instruments from relevant GWAS summary statistics using a genome-wide significance threshold (P < 5 × 10^-8^) and F-statistics greater than 10 to minimise weak instrument bias^54^. To ensure instrument independence, variants were clumped using stringent LD criteria (R^2^ < 0.001 within a 10 Mb window). Palindromic SNPs with intermediate allele frequencies were excluded to avoid strand ambiguity, variants absent from reference datasets were removed, and exposure and outcome data were harmonised to ensure consistent allele orientation and effect direction.

We used the inverse variance weighted (IVW) method as the primary MR approach, implemented with a multiplicative random-effects model to account for heterogeneity across instruments. Additional analyses included the weighted median estimator and MR-Egger regression to assess robustness under different assumptions regarding instrument validity and horizontal pleiotropy^72,73^. Robustness was evaluated using Cochran’s Q statistic to assess heterogeneity, leave-one-out analyses to examine the influence of individual instruments, and funnel plots to detect potential bias. Directional horizontal pleiotropy was assessed using the MR-Egger intercept test. The MR-PRESSO framework was applied to identify and remove pleiotropic outliers^74^. The framework detects horizontal pleiotropy through a global test, identifies individual pleiotropic variants using an outlier test, and re-estimates causal effects after exclusion of these variants^74^. In addition, the distortion test evaluates whether removing outlier SNPs significantly alters the original causal estimate, thereby assessing potential bias introduced by pleiotropy and enhancing the robustness of MR inference^74^. We defined nominal significance as P < 0.05, with a Bonferroni-corrected threshold of P < 0.017 applied to account for three unique traits tested. All analyses were conducted in R (version 4.5.2) using the TwoSampleMR package.

### Cross-disorder GWAS meta-analysis and functional annotation

We conducted a cross-disorder GWAS meta-analysis of AD and T2D summary statistics to identify potential shared genetic associations. Meta-analyses were performed using METASOFT, applying both the fixed-effect (FE) model and the modified random-effects model (RE2)^47^. The FE model assumes a common underlying genetic effect across disorders and maximises power under homogeneity, whereas the RE2 model accommodates between-trait heterogeneity in effect sizes while retaining sensitivity to shared associations, making the combination well-suited for cross-disorder analyses. SNPs common to both GWAS datasets were meta-analysed. In line with prior cross-trait studies^27,75,76^, we prioritised variants that did not reach genome-wide significance (GWS; P_meta-analysis_ < 5 × 10^-8^) in either individual GWAS (5 × 10^-8^ < P_meta-analysis_ < 0.001) but attained GWS following meta-analysis, as these may reflect novel shared susceptibility loci. We also examined SNPs that are already GWS in one disorder (sentinel) that showed evidence of association with the other. Genome-wide significance was defined as P < 5 × 10^-8^, with P < 1 × 10^-5^ considered suggestive. To evaluate whether association signals were supported across disorders, we applied the binary effect (BE) p-value and m-value framework^48^. The BE p-value tests for evidence of association in at least one dataset, while m-values estimate the posterior probability that a SNP has an effect in each disorder, with values >0.9 indicating strong evidence and <0.1 indicating no effect^48^.

Independent significant SNPs were identified using LD thresholds of r^2^ < 0.6, and lead SNPs were defined using r^2^ < 0.1. Genomic loci were delineated as ±250 kb regions centred on lead SNPs, with overlapping regions merged into a single locus (it is possible to have more than one lead SNP). Loci reaching GWS in the cross-disorder meta-analysis, including those not significant in either individual GWAS, were carried forward for annotation. Functional mapping and annotation were performed using Functional Mapping and Annotation (FUMA) ^77^, and identified loci were cross-referenced with the GWAS Catalog to assess prior associations with AD and T2D.

### Assessing shared genetic signals between AD and T2D using GWAS-PW colocalisation

To investigate shared genetic signals between AD and T2D, we performed a Bayesian analysis of shared and distinct causal configurations using the GWAS pair-wise (GWAS-PW) framework^53^. GWAS-PW is a hierarchical Bayesian method designed to scan the genome for loci that are statistically consistent with either shared or distinct causal configurations between two traits (sometimes referred to as pleiotropy, although we follow Pickrell et al. in avoiding this terminology here)^53^. For each approximately independent genomic region, the method estimates posterior probabilities for four mutually exclusive models: (1) the region contains a genetic variant influencing AD only (Model 1; PPA1), (2) the region contains a variant influencing T2D only (Model 2; PPA2), (3) the region contains a single variant influencing both AD and T2D (Model 3; PPA3), or (4) the region contains two distinct variants influencing AD and T2D independently (Model 4; PPA4)^53^. GWAS summary statistics for AD and T2D were harmonised to ensure consistent allelic orientation across traits. Variants were matched by rsID and allele information, and effect estimates were aligned to the same reference allele. Signed Z-scores and corresponding variances for each SNP were then used as input to the GWAS-PW model.

We conducted analyses across approximately independent genomic regions defined based on LD patterns derived from European-ancestry reference data, the 1000 Genomes Project. Because GWAS-PW explicitly models correlation in summary statistics arising from overlapping samples, we assessed potential sample overlap between the AD and T2D GWAS. No correction for sample overlap was required, where overlap was unlikely. Our primary interest was in regions showing posterior support for Model 3 or Model 4. Regions with PPA3 > 0.9 were considered as having strong support for a shared-variant configuration between AD and T2D, whereas values of 0.5 < PPA3 ≤ 0.9 were considered to indicate moderate support. Conversely, regions with PPA4 > 0.9 were interpreted as being more consistent with distinct causal variants affecting each trait within the same locus, with 0.5 < PPA4 ≤ 0.9 indicating moderate support for this configuration. Within the GWAS-PW framework, Bayes factors are derived at both the SNP and regional levels. SNP-level Bayes factors (logBF3) quantify the relative compatibility of individual variants with a shared-variant configuration, conditional on the region and model, and are used to prioritise representative variants within regions. Regional Bayes factors (logBF3, logBF4) are obtained by integrating SNP-level evidence across approximately independent genomic regions and quantify the relative support for shared versus distinct causal configurations. PPA3 and PPA4, derived from these regional Bayes factors, were used for primary model inference.

### Gene-based association analysis

To complement SNP-level analyses and improve biological interpretability, we conducted gene-based association analyses using MAGMA^56^, on the FUMA platform^77^ (https://fuma.ctglab.nl). This approach aggregates the effects of multiple SNPs within a gene into a single test statistic, thereby increasing power to detect modest but coordinated genetic effects that may be missed at the single-variant level^27,41,42^. Gene-based analyses were performed separately for AD (Jansen et al.) and T2D (Mahajan et al.) using GWAS summary statistics from each study. Following the practice in previous studies^75,78-81^, only SNPs overlapping between the respective GWAS datasets and the MAGMA reference panel were included. SNPs were assigned to genes based on physical position using strict gene boundaries (±0 kb from annotated transcription start and end sites). Gene-level association P-values (P_gene_) were computed independently for AD and T2D and subsequently used for downstream cross-trait analyses.

### Identification of shared and pleiotropic genes

To identify genes with evidence of shared genetic association between AD and T2D, we applied a cross-trait gene-based p-value aggregation using Stouffer’s Z-score method. This approach combines gene-level association evidence across traits by converting P_gene_ values into Z-scores and averaging them, rather than summing log-transformed P-values as in Fisher’s method^57^. Compared with Fisher’s method, Stouffer’s approach is less influenced by extremely small P-values and preferentially highlights genes showing evidence of association across traits^57^.

Gene-level Z-scores derived independently from AD and T2D MAGMA analyses outputs were combined using equal weights. Equal weighting was deliberately chosen to avoid dominance of one trait over the other and to ensure balanced contribution from both traits. Under this framework, genes were required to show evidence of association in both AD and T2D to achieve statistical significance in the combined analysis, rendering the approach conservative but favouring robust shared signals. Statistical significance for gene-based analyses was defined using a Bonferroni-corrected threshold of P_gene_ < 2.64 × 10^−6^, corresponding to correction for 18,965 tested genes. Using this framework, we classified shared genes into three categories: (i) genes reaching GWS in AD and nominal significance in T2D, (ii) genes reaching GWS in T2D and nominal significance in AD, and (iii) genes not GWS in either trait individually but achieving this status through the Stouffer’s combined P-value analysis, representing potentially novel shared genes based entirely on the joint evidence across both disorders GWAS.

### Summary-data–based Mendelian randomisation (SMR) and regulatory prioritisation

We applied SMR^49,50^ to test whether genetically predicted gene expression or DNA methylation levels influence AD and T2D. SMR integrates GWAS summary statistics with cis-expression QTL or methylation QTL data to prioritise genes whose regulatory variation may mediate disease associations through shared causal variants rather than linkage^49^. For eQTL-based SMR analyses, we used cis-eQTL summary statistics from GTEx v8 (multiple tissues), eQTLGen (whole blood), and BrainMeta (brain-specific datasets)^50^. For each gene, we selected the top cis-eQTL variant (within ±1 Mb of the transcription start site) as the instrumental variable^49^. SMR was performed separately using GWAS summary statistics for AD and T2D. We retained gene–trait associations that survived FDR correction (q < 0.05), were supported by at least 10 independent cis-eQTL instruments, and showed no evidence of heterogeneity using the HEIDI test (pHEIDI > 0.01; nHEIDI ≥ 10), indicating consistency with a shared causal variant model rather than linkage. We then intersected genes showing significant SMR associations for both AD and T2D within the same eQTL resource and regulatory feature. That is, overlap required concordance at the gene–eQTL dataset (tissue/platform) level. Directionality of effects was assessed by comparing the sign of SMR estimates across traits, allowing classification into concordant (same-direction) versus antagonistic (opposite-direction) regulatory effects.

For mQTL-based SMR analyses, we conducted analogous analyses using cis-mQTL summary statistics from BrainMeta and the McRae blood mQTL resource^50^. CpG sites within ±1 Mb of annotated genes were tested as instruments. We applied the same filtering criteria as for eQTL-SMR, retaining associations that passed FDR correction, were supported by at least 10 cis-mQTL instruments, and showed no evidence of heterogeneity (pHEIDI > 0.01; nHEIDI ≥ 10). Prioritised genes required concordance at the gene–mQTL dataset–CpG level, ensuring shared regulatory signals were evaluated within identical regulatory contexts across traits. Effect directions were compared across traits to classify concordant versus antagonistic methylation-mediated effects.

To prioritise high-confidence shared regulatory mechanisms, we adopted a conservative matching strategy requiring overlap within the same QTL resource and regulatory feature (gene–eQTL dataset for expression and gene–mQTL dataset–CpG site for methylation). This approach minimises false convergence arising from tissue- or platform-specific regulatory heterogeneity and strengthens inference that shared signals reflect common regulatory contexts rather than distinct mechanisms acting on the same gene. While this strategy may be conservative for broader tissue-shared effects, it enhances the specificity and mechanistic interpretability of the prioritised loci. Genes prioritised independently by eQTL-SMR and mQTL-SMR were intersected to identify candidates supported across multiple regulatory layers. We also examined genomic clustering of prioritised genes to identify regulatory hotspots. Loci with multiple independently prioritised genes across expression and methylation analyses were considered regions of convergent regulatory influence. For canonical regions, including *APOE* and the MHC, HEIDI-filtered results were interpreted cautiously due to extensive linkage disequilibrium and complex regulatory architecture. All analyses were conducted using the SMR software^50^ with default settings unless otherwise specified. Multiple testing was controlled using the Benjamini–Hochberg FDR procedure within each QTL resource and trait.

## Data Availability

The study used only openly available human data from:
1: https://www.diagram-consortium.org
2. https://www.ebi.ac.uk/gwas/
3. https://www.leelabsg.org/resources
4. https://ctg.cncr.nl/documents/p1651/AD_sumstats_Jansenetal_2019sept.txt.gz
All datasets used in this study are described in the main text and in the Supplementary Materials. Summary statistics were sourced from publicly accessible repositories and international consortia, as detailed in the Data Sources section. A complete list of datasets, including accession identifiers and direct access links, is provided in Supplementary Table 1.

## Statistical Analysis and Reproducibility

All primary statistical analyses were conducted within Unix-based environments and the R statistical platform (https://www.r-project.org/, version 4.5.2). Additional analyses were performed with other Unix-compatible tools, including Python (https://www.python.org/) and PLINK (https://www.cog-genomics.org/plink/^69^), and with locally implemented programs such as SECA^51^. Functional and genomic annotation procedures also made use of online platforms such as G: Profiler (https://biit.cs.ut.ee/gprofiler^59^) and FUMA (https://fuma.ctglab.nl^77^). Reproducibility was evaluated by replicating key findings across independent datasets, where available, and by applying complementary analytic approaches to confirm the robustness and consistency of results.

## Ethical Approval and Participant Consent

This work is a secondary analysis of publicly available, de-identified data derived from international research consortia and open-access repositories. As the study utilised only aggregated summary statistics with no identifiable information, no additional ethics approval or participant consent was required. All analyses adhered to relevant guidelines governing the secondary use and sharing of research data.

## Data Availability

All datasets used in this study are described in the main text and in the Supplementary Materials. Summary statistics were sourced from publicly accessible repositories and international consortia, as detailed in the Data Sources section. A complete list of datasets and appropriate references is provided in Supplementary Table 1.

## Code availability

All analyses were performed using publicly available software and online resources. URLs and documentation for each tool are as follows: G:Profiler: https://biit.cs.ut.ee/gprofiler, GWAS Catalogue: https://www.ebi.ac.uk/gwas/home, Open Targets Genetics: https://genetics.opentargets.org/, LDSC: https://github.com/bulik/ldsc, LAVA: https://github.com/josefin-werme/LAVA/tree/main/vignettes/data, Two-Sample MR: https://mrcieu.github.io/TwoSampleMR/articles/introduction.html, SMR: https://yanglab.westlake.edu.cn/software/smr/#Overview, FUMA: https://fuma.ctglab.nl, SECA: https://research.qut.edu.au/sgel/software/seca-local-version

## Acknowledgments

We acknowledge the research consortia and publicly available databases that provided access to the various omics data used in this work. We thank the investigators and participants of the FinnGen study. We are also grateful to the broader research community whose efforts enabled these data resources. Appreciation is extended to the developers and maintainers of the computational tools and platforms employed in this study. We thank Master John Adewuyi for assistance with figure preparation.

## Author contributions

EOA conceived and designed the study, curated datasets, conducted statistical analyses, and drafted the initial manuscript. EO, DRN, GP, and AA contributed to the interpretation of the results. AA contributed to the study design. DRN provided methodological and conceptual insights. OO assisted with data management and table preparation. AA, GP, KG and KKS provided cross-disciplinary insight. EO, AA, OO, KG, KKS, DRN, and GP contributed to critical revisions. All authors reviewed and approved the final manuscript for submission.

## Additional information

Supplementary Tables 1 – 23 are available as a Word document file.

## References

1 2023 Alzheimer’s disease facts and figures. Alzheimers Dement 19, 1598–1695 (2023).

2 Nichols, E. et al. Estimation of the global prevalence of dementia in 2019 and forecasted prevalence in 2050: an analysis for the Global Burden of Disease Study 2019. The Lancet Public Health 7, e105–e125 (2022).

3 Livingston, G. et al. Dementia prevention, intervention, and care: 2020 report of the Lancet Commission. The lancet 396, 413–446 (2020).

4 Roden, M. & Shulman, G. I. The integrative biology of type 2 diabetes. Nature 576, 51–60 (2019). 10.1038/s41586-019-1797-8

5 Galicia-Garcia, U. et al. Pathophysiology of Type 2 Diabetes Mellitus. International Journal of Molecular Sciences 21, 6275 (2020).

6 Sun, H. et al. IDF Diabetes Atlas: Global, regional and country-level diabetes prevalence estimates for 2021 and projections for 2045. Diabetes research and clinical practice 183, 109119 (2022).

7 Genitsaridi, I. et al. 11th edition of the IDF Diabetes Atlas: global, regional, and national diabetes prevalence estimates for 2024 and projections for 2050. The Lancet Diabetes & Endocrinology 14, 149–156 (2026). 10.1016/S2213-8587(25)00299-2

8 Duncan, B. B., Magliano, D. J. & Boyko, E. J. IDF Diabetes Atlas 11th edition 2025: global prevalence and projections for 2050. Nephrol Dial Transplant 41, 7–9 (2025). 10.1093/ndt/gfaf177

9 Gudala, K., Bansal, D., Schifano, F. & Bhansali, A. Diabetes mellitus and risk of dementia: A meta-analysis of prospective observational studies. Journal of diabetes investigation 4, 640–650 (2013).

10 Chatterjee, S. et al. Type 2 diabetes as a risk factor for dementia in women compared with men: a pooled analysis of 2.3 million people comprising more than 100,000 cases of dementia. Diabetes care 39, 300–307 (2016).

11 Biessels, G. J., Staekenborg, S., Brunner, E., Brayne, C. & Scheltens, P. Risk of dementia in diabetes mellitus: a systematic review. The Lancet Neurology 5, 64–74 (2006).

12 Cao, F. et al. The relationship between diabetes and the dementia risk: a meta-analysis. Diabetology & Metabolic Syndrome 16, 101 (2024).

13 Karmakar, A., Dutta, S. & Mukhopadhyay, D. in Diabetes and Neurodegeneration (eds Debasis Bagchi, Samudra Prosad Banik, & Rameshwar Nath Chaurasia) 407–431 (Academic Press, 2026).

14 Hassing, L. B. et al. Diabetes Mellitus Is a Risk Factor for Vascular Dementia, but Not for Alzheimer’s Disease: A Population-Based Study of the Oldest Old. International Psychogeriatrics 14, 239–248 (2002). 10.1017/S104161020200844X

15 MacKnight, C., Rockwood, K., Awalt, E. & McDowell, I. Diabetes mellitus and the risk of dementia, Alzheimer’s disease and vascular cognitive impairment in the Canadian Study of Health and Aging. Dement Geriatr Cogn Disord 14, 77–83 (2002). 10.1159/000064928

16 Santiago, J. A., Karthikeyan, M., Lackey, M., Villavicencio, D. & Potashkin, J. A. Diabetes: a tipping point in neurodegenerative diseases. Trends in molecular medicine 29, 1029–1044 (2023).

17 Hamzé, R. et al. Type 2 Diabetes Mellitus and Alzheimer’s Disease: Shared Molecular Mechanisms and Potential Common Therapeutic Targets. International Journal of Molecular Sciences 23, 15287 (2022).

18 Kellar, D. & Craft, S. Brain insulin resistance in Alzheimer’s disease and related disorders: mechanisms and therapeutic approaches. The Lancet Neurology 19, 758–766 (2020).

19 Arnold, S. E. et al. Brain insulin resistance in type 2 diabetes and Alzheimer disease: concepts and conundrums. Nature Reviews Neurology 14, 168–181 (2018).

20 Lynn, J., Park, M., Ogunwale, C. & Acquaah-Mensah, G. K. A Tale of Two Diseases: Exploring Mechanisms Linking Diabetes Mellitus with Alzheimer’s Disease. Journal of Alzheimer’s Disease 85, 485–501 (2022). 10.3233/jad-210612

21 Talbot, K. et al. Demonstrated brain insulin resistance in Alzheimer’s disease patients is associated with IGF-1 resistance, IRS-1 dysregulation, and cognitive decline. The Journal of clinical investigation 122, 1316–1338 (2012).

22 Craft, S. Insulin resistance and Alzheimer’s disease pathogenesis: potential mechanisms and implications for treatment. Current Alzheimer Research 4, 147–152 (2007).

23 Abner, E. L. et al. Diabetes is associated with cerebrovascular but not Alzheimer’s disease neuropathology. Alzheimer’s & Dementia 12, 882–889 (2016).

24 Arvanitakis, Z. et al. Diabetes is related to cerebral infarction but not to AD pathology in older persons. Neurology 67, 1960–1965 (2006).

25 dos Santos Matioli, M.N.P. et al. Diabetes is not associated with Alzheimer’s disease neuropathology. Journal of Alzheimer’s disease 60, 1035–1043 (2017).

26 Jansen, I. E. et al. Genome-wide meta-analysis identifies new loci and functional pathways influencing Alzheimer’s disease risk. Nature genetics 51, 404–413 (2019).

27 Adewuyi, E. O., O’Brien, E. K., Nyholt, D. R., Porter, T. & Laws, S. M. A large-scale genome-wide crosstrait analysis reveals shared genetic architecture between Alzheimer’s disease and gastrointestinal tract disorders. Communications biology 5, 691 (2022).

28 Mahajan, A. et al. Fine-mapping type 2 diabetes loci to single-variant resolution using high-density imputation and islet-specific epigenome maps. Nature Genetics 50, 1505–1513 (2018). 10.1038/s41588-018-0241-6

29 Lambert, J. C. et al. Meta-analysis of 74,046 individuals identifies 11 new susceptibility loci for Alzheimer’s disease. Nat Genet 45, 1452–1458 (2013). 10.1038/ng.2802

30 Xue, A. et al. Genome-wide association analyses identify 143 risk variants and putative regulatory mechanisms for type 2 diabetes. Nature Communications 9, 2941 (2018). 10.1038/s41467-018-04951-w

31 Bulik-Sullivan, B. K. et al. LD Score regression distinguishes confounding from polygenicity in genome-wide association studies. Nature genetics 47, 291–295 (2015). 10.1038/ng.3211

32 Zhu, Z., Lin, Y., Li, X., Driver, J. A. & Liang, L. Shared genetic architecture between metabolic traits and Alzheimer’s disease: a large-scale genome-wide cross-trait analysis. Human Genetics 138, 271–285 (2019). 10.1007/s00439-019-01988-9

33 Hardy, J., de Strooper, B. & Escott-Price, V. Diabetes and Alzheimer’s disease: shared genetic susceptibility? The Lancet Neurology 21, 962–964 (2022). 10.1016/S1474-4422(22)00395-7

34 Hu, Z. et al. Shared Causal Paths underlying Alzheimer’s dementia and Type 2 Diabetes. Scientific Reports 10, 4107 (2020). 10.1038/s41598-020-60682-3

35 Shu, J., Li, N., Wei, W. & Zhang, L. Detection of molecular signatures and pathways shared by Alzheimer’s disease and type 2 diabetes. Gene 810, 146070 (2022). 10.1016/j.gene.2021.146070

36 Hao, K. et al. Shared genetic etiology underlying Alzheimer’s disease and type 2 diabetes. Molecular aspects of medicine 43, 66–76 (2015).

37 Adewuyi, E. O., Porter, T., Verdile, G. & Laws, S. M. Assessing genetic overlap of Alzheimer’s disease with type 2 diabetes. Alzheimer’s & Dementia 20, e090165 (2024). 10.1002/alz.090165

38 Chornenkyy, Y., Wang, W. X., Wei, A. & Nelson, P. T. Alzheimer’s disease and type 2 diabetes mellitus are distinct diseases with potential overlapping metabolic dysfunction upstream of observed cognitive decline. Brain Pathology 29, 3–17 (2019).

39 Han, S., Lelieveldt, T., Sturkenboom, M., Biessels, G. J. & Ahmadizar, F. Evaluating the Causal Association Between Type 2 Diabetes and Alzheimer’s Disease: A Two-Sample Mendelian Randomization Study. Biomedicines 13, 1095 (2025).

40 Qi, G. et al. Genome-wide large-scale multi-trait analysis characterizes global patterns of pleiotropy and unique trait-specific variants. Nature communications 15, 6985 (2024).

41 Adewuyi, E. O. & Laws, S. M. Genomic Characterisation of the Relationship and Causal Links Between Vascular Calcification, Alzheimer’s Disease, and Cognitive Traits. Biomedicines 13, 618 (2025).

42 Adewuyi, E. O. et al. Genome-wide cross-disease analyses highlight causality and shared biological pathways of type 2 diabetes with gastrointestinal disorders. Communications Biology 7, 643 (2024). 10.1038/s42003-024-06333-z

43 Zhang, Z. et al. A scalable approach to characterize pleiotropy across thousands of human diseases and complex traits using GWAS summary statistics. The American Journal of Human Genetics 110, 1863–1874 (2023). 10.1016/j.ajhg.2023.09.015

44 Xu, Q., Yang, C. & Pei, Y.-F. Editorial: Genetic Pleiotropy in Complex Traits and Diseases. Frontiers in Genetics Volume 13 -2022 (2022). 10.3389/fgene.2022.897383

45 Kulminski, A. M. et al. Prevailing antagonistic risks in pleiotropic associations with Alzheimer’s disease and diabetes. Journal of Alzheimer’s Disease 94, 1121–1132 (2023).

46 Loika, Y., Loiko, E., Culminskaya, I. & Kulminski, A. M. Exome-Wide Association Study Identified Clusters of Pleiotropic Genetic Associations with Alzheimer’s Disease and Thirteen Cardiovascular Traits. Genes 14, 1834 (2023).

47 Han, B. & Eskin, E. Random-effects model aimed at discovering associations in meta-analysis of genome-wide association studies. The American Journal of Human Genetics 88, 586–598 (2011).

48 Han, B. & Eskin, E. Interpreting meta-analyses of genome-wide association studies. PLoS genetics 8, e1002555 (2012).

49 Zhu, Z. et al. Integration of summary data from GWAS and eQTL studies predicts complex trait gene targets. Nature Genetics 48, 481–487 (2016). 10.1038/ng.3538

50 Guo, Y. et al. SMR-Portal: an online platform for integrative analysis of GWAS and xQTL data to identify complex trait genes. Nature Methods 22, 220–222 (2025). 10.1038/s41592-024-02561-7

51 Nyholt, D. R. SECA: SNP effect concordance analysis using genome-wide association summary results. Bioinformatics 30, 2086–2088 (2014). 10.1093/bioinformatics/btu171

52 Werme, J., van der Sluis, S., Posthuma, D. & de Leeuw, C. A. An integrated framework for local genetic correlation analysis. Nature Genetics 54, 274–282 (2022). 10.1038/s41588-022-01017-y

53 Pickrell, J. K. et al. Detection and interpretation of shared genetic influences on 42 human traits. Nat Genet 48, 709–717 (2016). 10.1038/ng.3570

54 Davies, N. M., Holmes, M. V. & Davey Smith, G. Reading Mendelian randomisation studies: a guide, glossary, and checklist for clinicians. BMJ 362, k601 (2018). 10.1136/bmj.k601

55 Sekula, P., Del Greco M, F., Pattaro, C. & Köttgen, A. Mendelian Randomization as an Approach to Assess Causality Using Observational Data. J Am Soc Nephrol 27, 3253–3265 (2016). 10.1681/ASN.2016010098

56 de Leeuw, C. A., Mooij, J. M., Heskes, T. & Posthuma, D. MAGMA: Generalized Gene-Set Analysis of GWAS Data. PLOS Computational Biology 11, e1004219 (2015). 10.1371/journal.pcbi.1004219

57 Whitlock, M. C. Combining probability from independent tests: the weighted Z-method is superior to Fisher’s approach. Journal of evolutionary biology 18, 1368–1373 (2005).

58 Kurki, M. I. et al. FinnGen provides genetic insights from a well-phenotyped isolated population. Nature 613, 508–518 (2023). 10.1038/s41586-022-05473-8

59 Raudvere, U. et al. g:Profiler: a web server for functional enrichment analysis and conversions of gene lists (2019 update). Nucleic Acids Research 47, W191–W198 (2019). 10.1093/nar/gkz369

60 Marioni, R. E. et al. GWAS on family history of Alzheimer’s disease. Transl Psychiatry 8, 99 (2018). 10.1038/s41398-018-0150-6

61 Imamura, M. et al. Genome-wide association studies in the Japanese population identify seven novel loci for type 2 diabetes. Nat Commun 7, 10531 (2016). 10.1038/ncomms10531

62 Mansour Aly, D. et al. Genome-wide association analyses highlight etiological differences underlying newly defined subtypes of diabetes. Nat Genet 53, 1534–1542 (2021). 10.1038/s41588-021-00948-2

63 Hao, K. et al. Shared genetic etiology underlying Alzheimer’s disease and type 2 diabetes. Mol Aspects Med 43-44, 66–76 (2015). 10.1016/j.mam.2015.06.006

64 Chen, D. W., Shi, J. K., Li, Y., Yang, Y. & Ren, S. P. Association between ApoE Polymorphism and Type 2 Diabetes: A Meta-Analysis of 59 Studies. Biomed Environ Sci 32, 823–838 (2019). 10.3967/bes2019.104

65 Sapkota, B. et al. Association of APOE polymorphisms with diabetes and cardiometabolic risk factors and the role of APOE genotypes in response to anti-diabetic therapy: results from the AIDHS/SDS on a South Asian population. J Diabetes Complications 29, 1191–1197 (2015). 10.1016/j.jdiacomp.2015.07.025

66 Kim, J. P. et al. Cross-ancestry genome-wide association study identifies implications of SORL1 in cerebral beta-amyloid deposition. Nat Commun 16, 3150 (2025). 10.1038/s41467-025-57751-4

67 Wang, X.-F. et al. Linking Alzheimer’s disease and type 2 diabetes: Novel shared susceptibility genes detected by cFDR approach. Journal of the Neurological Sciences 380, 262–272 (2017). 10.1016/j.jns.2017.07.044

68 Khurana, V. & Goswami, B. Angiotensin converting enzyme (ACE). Clinica Chimica Acta 524, 113–122 (2022). 10.1016/j.cca.2021.10.029

69 Purcell, S. et al. PLINK: a tool set for whole-genome association and population-based linkage analyses. The American journal of human genetics 81, 559–575 (2007).

70 Richmond, R. C. & Davey Smith, G. Mendelian Randomization: Concepts and Scope. Cold Spring Harb Perspect Med 12 (2022). 10.1101/cshperspect.a040501

71 Skrivankova, V. W. et al. Strengthening the reporting of observational studies in epidemiology using Mendelian randomization: the STROBE-MR statement. Jama 326, 1614–1621 (2021).

72 Bowden, J., Davey Smith, G., Haycock, P. C. & Burgess, S. Consistent estimation in Mendelian randomization with some invalid instruments using a weighted median estimator. Genetic epidemiology 40, 304–314 (2016).

73 Bowden, J., Davey Smith, G. & Burgess, S. Mendelian randomization with invalid instruments: effect estimation and bias detection through Egger regression. International Journal of Epidemiology 44, 512–525 (2015). 10.1093/ije/dyv080

74 Verbanck, M., Chen, C.-y., Neale, B. & Do, R. Detection of widespread horizontal pleiotropy in causal relationships inferred from Mendelian randomization between complex traits and diseases. Nature genetics 50, 693–698 (2018).

75 Adewuyi, E. O., Mehta, D. & Nyholt, D. Genetic overlap analysis of endometriosis and asthma identifies shared loci implicating sex hormones and thyroid signalling pathways. Human Reproduction 37, 366–383 (2022).

76 Islam, M. R., Nyholt, D. R. & The International Headache Genetics, C. Cross-trait analyses identify shared genetics between migraine, headache, and glycemic traits, and a causal relationship with fasting proinsulin. Human Genetics (2023). 10.1007/s00439-023-02532-6

77 Watanabe, K., Taskesen, E., van Bochoven, A. & Posthuma, D. Functional mapping and annotation of genetic associations with FUMA. Nature Communications 8, 1826 (2017). 10.1038/s41467-017-01261-5

78 Adewuyi, E. O. et al. Genetic analysis of endometriosis and depression identifies shared loci and implicates causal links with gastric mucosa abnormality. Human Genetics 140, 529–552 (2021). 10.1007/s00439-020-02223-6

79 Adewuyi, E. O., O’Brien, E. K., Porter, T. & Laws, S. M. Relationship of Cognition and Alzheimer&rsquo;s Disease with Gastrointestinal Tract Disorders: A Large-Scale Genetic Overlap and Mendelian Randomisation Analysis. International Journal of Molecular Sciences 23, 16199 (2022).

80 Adewuyi, E. O. et al. Shared Molecular Genetic Mechanisms Underlie Endometriosis and Migraine Comorbidity. Genes 11, 268 (2020).

81 Yang, Y. et al. Molecular genetic overlap between migraine and major depressive disorder. European Journal of Human Genetics 26, 1202–1216 (2018). 10.1038/s41431-018-0150-2

