## Supplementary Tables for "Cross-omic dissection reveals locus-specific heterogeneity and antagonistic pleiotropy between Alzheimer’s disease and type 2 diabetes"

### Supplementary Table 1: Description of GWAS data analysed in this study with links to the source

| **Data** | **Cases** | **control** | **Sample size** | **Ancestry** | **Phenotype source/definition** |
| --- | --- | --- | --- | --- | --- |
| T2D Mahajan et al 2018 | 74,124 | 824,006 | 898130 | European | T2D data from the DIAGRAM consortium |
| T2D Xue et al 2018 | 62,832 | 596,424 | 659256 |  | Meta-analysis of genome-wide association studies (GWAS) in 62,892 T2D cases and 596,424 controls of European ancestry, by combining three GWAS data sets: DIAbetes Genetics Replication And Meta-analysis (DIAGRAM), Genetic Epidemiology Research on Aging (GERA) and the full cohort release of the UK Biobank (UKB). |
| UKB T2D Phecode 250.2 (Lee Lab) | 18,945 | 596,424 | 615369 |  | Full White British samples from the Lee Lab |
| T2D FinnGen-R10 | 65085 | 335112 | 400197 |  | T2D (FinnGen) refers to the FinnGen Type 2 diabetes phenotype constructed using combined registry-based diagnostic and medication definitions |
| AD Jansen et al 2019 | 71,880 | 383,378 | 455,258 |  | Summary data for GWAS of Alzheimer's diseases, comprising clinically diagnosed cases and AD-by-proxy cases |
| AD Lambert et al 2013 | 17,008 | 37,154 | 54,162 |  | Summary data for GWAS of Alzheimer's disease comprising clinically diagnosed cases |

### Supplementary Table 2: Global genetic correlation between Alzheimer’s disease and type 2 diabetes

| Trait 1 | Trait 2 | rG | SE | P |
| --- | --- | --- | --- | --- |
| Including the *APOE* region | | | | |
| ^a^AD Jansen et al ^18^ | T2D Mahajan et al ^20^ | 0.14 | 0.04 | 1.43 x 10^-04^ |
|  | T2D adjusted for BMI ^20^ | 0.13 | 0.03 | 2.44 x 10^-04^ |
|  | T2D UKB Phecode 250.2 | 0.19 | 0.05 | 7.68 x 10^-05^ |
|  | T2D Xue et al ^27^ | 0.17 | 0.03 | 3.81 x 10^-08^ |
|  | T2D FinnGen-R10 ^28^ | 0.10 | 0.03 | 1.05 x 10^-04^ |
| ^b^AD Lambert et al ^26^ | T2D Mahajan et al ^20^ | 0.09 | 0.03 | 2.39 x 10^-03^ |
|  | T2D adjusted for BMI ^20^ | 0.09 | 0.03 | 1.57 x 10^-03^ |
|  | T2D UKB Phecode 250.2 | 0.06 | 0.04 | 1.02 x 10^-01^ |
|  | T2D Xue et al ^27^ | 0.07 | 0.03 | 1.30 x 10^-02^ |
|  | T2D FinnGen-R10 ^28^ | 0.05 | 0.02 | 1.44 x 10^-02^ |
| Excluding the *APOE* region | | | | |
| ^a^AD Jansen et al ^18^ | T2D Mahajan et al ^20^ | 0.17 | 0.03 | 2.07 x 10^-10^ |
|  | T2D adjusted for BMI ^20^ | 0.15 | 0.03 | 1.41 x 10^-07^ |
|  | T2D UKB Phecode 250.2 | 0.22 | 0.04 | 1.80 x 10^-09^ |
|  | T2D Xue et al ^27^ | 0.18 | 0.03 | 3.03 x 10^-09^ |
|  | T2D FinnGen-R10 ^28^ | 0.11 | 0.02 | 2.73 x 10^-08^ |
| ^b^AD Lambert et al ^26^ | T2D Mahajan et al ^20^ | 0.10 | 0.03 | 7.01 x 10^-05^ |
|  | T2D adjusted for BMI ^20^ | 0.10 | 0.03 | 3.06 x 10^-04^ |
|  | T2D UKB Phecode 250.2 | 0.07 | 0.03 | 4.30 x 10^-02^ |
|  | T2D Xue et al ^27^ | 0.07 | 0.03 | 7.80 x 10^-03^ |
|  | T2D FinnGen-R10 ^28^ | 0.06 | 0.02 | 3.77 x 10^-03^ |

AD: Alzheimer’s disease, T2D: type 2 diabetes, rG: genetic correlation, SE: standard error, P: p-value, UKB: United Kingdom Biobank, ^a^AD: combining clinically diagnosed with proxy cases, ^b^AD: comprising only clinically diagnosed cases.

### Supplementary Table 3: pairwise genetic correlations between different T2D GWAS

| **Trait 1** | **Trait 2** | **rG** | **SE** | **p** | **Gcov int** | **Gcov int se** |
| --- | --- | --- | --- | --- | --- | --- |
| T2D Mahajan et al ^20^ | T2Dbmiadj.sumstats.gz | 0.91 | 0.01 | 0.00E+00 | 0.92 | 0.02 |
|  | T2-diabetes.sumstats.gz | 1.01 | 0.01 | 0.00E+00 | 0.43 | 0.01 |
|  | T2D_Xue-2018.sumstats.gz | 1.02 | 0.01 | 0.00E+00 | 0.56 | 0.02 |
| T2D Xue et al ^27^ | T2-diabetes.sumstats.gz | 1.01 | 0.01 | 0.00E+00 | 0.72 | 0.01 |
| T2Dbmiadj Mahajan et al ^20^ | T2-diabetes.sumstats.gz | 0.89 | 0.02 | 0.00E+00 | 0.49 | 0.01 |
|  | T2D_Xue-2018.sumstats.gz | 0.93 | 0.01 | 0.00E+00 | 0.48 | 0.02 |

### Supplementary Table 4: SECA Primary test for SNP effects concordance and effect direction association

| **Trait 1** | **Trait 2** | **Test for SNP effect concordance** | | | **Test for the association between effect direction** | |
| --- | --- | --- | --- | --- | --- | --- |
|  |  | **Direction** | **SNP sets ratio ^c^** | **P_permuted_ ^d^** | **OR (95%CI)** | **P _Fisher's-exact-test_^e^** |
| Including the APOE region | | | | | | |
| AD Jansen et al ^18^ | T2D Mahajan et al ^20^ | + | 143/144 | 0.001 | 1.49 (1.26 – 1.78) | 4.06 x 10^-06^ |
|  | T2D Xue et al ^27^ | + | 144/144 | 0.001 | 1.74 (1.30 – 2.32) | 1.56 x 10^-04^ |
| T2D Mahajan et al ^20^ | AD Jansen et al ^18^ | + | 132/144 | 0.001 | 1.36 (1.11 – 1.67) | 2.80 x 10^-03^ |
| T2D Xue et al ^27^ |  | + | 133/144 | 0.001 | 1.49 (1.15 – 1.92) | 1.82 x 10^-03^ |
| AD Lambert et al ^26^ | T2D Mahajan et al ^20^ | + | 142/144 | 0.001 | 1.38 (1.15 – 1.64) | 3.46 x 10^-04^ |
|  | T2D Xue et al ^27^ | + | 123/144 | 0.001 | 1.53 (1.16 – 2.02) | 2.52 x 10^-03^ |
| T2D Mahajan et al ^20^ | AD Lambert et al ^26^ | + | 128/144 | 0.001 | 1.23 (0.99 – 1.56) | 7.49 x 10^-02^ |
| T2D Xue et al ^27^ |  | + | 123/144 | 0.001 | 1.41 (1.09 – 1.82) | 9.05 x 10^-03^ |
| Excluding the APOE region | | | | | | |
| AD Jansen et al ^18^ | T2D Mahajan et al ^20^ | + | 144/144 | 0.001 | 1.57 (1.32 – 1.87) | 2.67 x 10^-07^ |
|  | T2D Xue et al ^27^ | + | 144/144 | 0.001 | 1.78 (1.31 – 2.35) | 1.14 x 10^-04^ |
| T2D Mahajan et al ^20^ | AD Jansen et al ^18^ | + | 135/144 | 0.001 | 1.41 (1.15 – 1.73) | 9.32 x 10^-04^ |
| T2D Xue et al ^27^ |  | + | 138/144 | 0.001 | 1.50 (1.17 – 1.94) | 1.42 x 10^-03^ |
| AD Lambert et al ^26^ | T2D Mahajan et al ^20^ | + | 143/144 | 0.001 | 1.42 (1.19 – 1.70) | 8.17 x 10^-05^ |
|  | T2D Xue et al ^27^ | + | 131/144 | 0.001 | 1.58 (1.19 – 2.09) | 1.23 x 10^-03^ |
| T2D Mahajan et al ^20^ | AD Lambert et al ^26^ | + | 131/144 | 0.001 | 1.26 (1.00 – 1.59) | 4.98 x 10^-02^ |
| T2D Xue et al ^27^ |  | + | 125/144 | 0.001 | 1.43 (1.11 – 1.86) | 6.09 x 10^-03^ |

AD: Alzheimer’s disease, T2D: type 2 diabetes, P: p-value, UKB: United Kingdom Biobank, ^c^ The ratio of the number of SNP subsets with significant concordant effects between AD and T2D traits at P_SNP_ < 0.05. ^d^ Permuted P for observing significant SNP subsets as the observation. SECA calculates the permuted P value for the number of significant associations with adjustment for testing 144 associations (based on permutations of 1000 replicates). ^e^Testing for the association between effect direction in Trait 1 SNPs and Trait 2 SNPs at P_SNP_ < 0.05 using the Fisher’s exact test (two-sided). Note: The SNP effects in trait 1 and trait 2 were POSITIVELY correlated. If significant, the results indicate the presence of allelic effects that increase the risk for both traits.

### Supplementary Table 5: SECA results: primary test for genetic overlap between AD and T2D

| Trait 1 (P1) | Trait 2 (P2) | Expected proportion of overlap | Overlap test at P1 < 0.05 and P2 < 0.05 | | Overlap test at P1 < 1e-05 and P2 < 0.05 | | Overlap test at P1 < 5e-08 and P2 < 0.05 | |
| --- | --- | --- | --- | --- | --- | --- | --- | --- |
|  |  |  | **Observed proportion of overlap (%)** | **P_binomial-test (two-sided)_** | **Observed proportion of overlap (%)** | **P_binomial-test (two-sided)_** | **Observed proportion of overlap (%)** | **P_binomial-test (two-sided)_** |
| Including the APOE region | | | | | | | | |
| AD Jansen et al ^18^ | T2D Mahajan et al ^20^ | 9.47% | 9.94% | 1.93 x 10^-02^ | 25.64% | 4.22 x 10^-09^ | 32.43% | 4.15 x 10^-08^ |
|  | T2D Xue et al ^27^ | 9.60% | 10.46% | 1.24 x 10^-02^ | 22.41% | 2.96 x 10^-03^ | 17.39% | 2.72 x 10^-01^ |
| T2D Mahajan et al ^20^ | AD Jansen et al ^18^ | 5.62% | 5.98% | 1.19 x 10^-02^ | 9.01% | 6.73 x 10^-06^ | 12.72% | 1.22 x 10^-07^ |
| T2D Xue et al ^27^ |  | 5.86% | 6.43% | 2.46 x 10^-03^ | 11.67% | 2.92 x 10^-06^ | 14.29% | 7.22 x 10^-05^ |
| AD Lambert et al ^26^ | T2D Mahajan et al ^20^ | 9.94% | 10.26% | 1.33 x 10^-01^ | 30.95% | 8.99 x 10^-08^ | 33.33% | 6.17 x 10^-05^ |
|  | T2D Xue et al ^27^ | 9.26% | 9.68% | 1.79 x 10^-01^ | 17.65% | 1.26 x 10^-01^ | 23.08% | 1.13 x 10^-01^ |
| T2D Mahajan et al ^20^ | AD Lambert et al ^26^ | 5.77% | 5.93% | 3.30 x 10^-01^ | 8.59% | 4.08 x 10^-04^ | 11.02% | 1.34 x 10^-04^ |
| T2D Xue et al ^27^ |  | 6.08% | 6.36% | 1.44 x 10^-01^ | 9.38% | 6.54 x 10^-03^ | 12.50% | 3.10 x 10^-03^ |
| Excluding the APOE region | | | | | | | | |
| AD Jansen et al ^18^ | T2D Mahajan et al ^20^ | 9.45% | 9.89% | 2.58 x 10^-02^ | 20.56% | 4.10 x 10^-04^ | 21.21% | 3.19 x 10^-02^ |
|  | T2D Xue et al ^27^ | 9.62% | 10.43% | 1.71 x 10^-02^ | 19.23% | 3.00 x 10^-02^ | 11.11% | 6.90 x 10^-01^ |
| T2D Mahajan et al ^20^ | AD Jansen et al ^18^ | 5.59% | 5.94% | 1.63 x 10^-02^ | 8.84% | 1.57 x 10^-05^ | 12.72% | 1.14 x 10^-07^ |
| T2D Xue et al ^27^ |  | 5.83% | 6.40% | 2.40 x 10^-03^ | 11.67% | 1.85 x 10^-06^ | 14.29% | 6.69 x 10^-05^ |
| AD Lambert et al ^26^ | T2D Mahajan et al ^20^ | 9.92% | 10.23% | 1.45 x 10^-01^ | 25.86% | 4.16 x 10^-04^ | 21.43% | 1.56 x 10^-01^ |
|  | T2D Xue et al ^27^ | 9.25% | 9.66% | 1.85 x 10^-01^ | 13.33% | 3.54 x 10^-01^ | 11.11% | 5.82 x 10^-01^ |
| T2D Mahajan et al ^20^ | AD Lambert et al ^26^ | 5.75% | 5.89% | 3.75 x 10^-01^ | 8.50% | 5.16 x 10^-04^ | 11.02% | 1.28 x 10^-04^ |
| T2D Xue et al ^27^ |  | 6.06% | 6.34% | 1.34 x 10^-01^ | 9.38% | 6.41 x 10^-03^ | 12.50% | 3.03 x 10^-03^ |

SECA: Single nucleotide polymorphism (SNP) effect concordant analysis, AD: Alzheimer’s disease, T2D: type 2 diabetes. Note: The analysis was performed in SECA, using independent SNPs for Trait 1. The proportion of overlap refers to the independent SNPs overlapping between the two traits at the stated p-value threshold for each of the two traits. The exact binomial test (two-sided) was used to assess whether the proportion of SNP overlap between the two traits was more than expected by chance, comparing the expected with the observed proportion of overlap. Two-sided indicates that the test assessed whether the observed proportion of overlap was significantly more (the first direction) or less (the second direction) than the expected proportion of overlap. When the number of SNPs associated with both datasets was significantly more than expected (P_binomial-test_ < 0.05), the results indicate the presence of pleiotropy, i.e., some genetic variants influence both traits) at the stated threshold.

### Supplementary Table 6: Local genetic correlation between AD and T2D

| **Phen1** | **Phen2** | **locus** | **chr** | **start** | **stop** | **rho** | **Rho lower** | **Rho upper** | **r2** | **r2 lower** | **r2 upper** | **P** |
| --- | --- | --- | --- | --- | --- | --- | --- | --- | --- | --- | --- | --- |
| AD (Jansen et al) | T2D (Mahajan et al) | 2351 | 19: | 45040933 | 45893307 | -0.61 | -0.70 | -0.52 | 0.37 | 0.27 | 0.49 | 4.89 x 10^-33^ |
|  |  | 962 | 6 | 32208902 | 32454577 | -0.37 | -0.57 | -0.17 | 0.14 | 0.03 | 0.32 | 4.18 x 10^-04^ |
|  |  | 965 | 6 | 32586785 | 32629239 | -0.14 | -0.21 | -0.06 | 0.02 | 0.00 | 0.04 | 6.14 x 10^-04^ |
|  |  | 960 | 6 | 31320269 | 31427209 | -0.53 | -0.86 | -0.22 | 0.28 | 0.05 | 0.75 | 1.64 x 10^-03^ |
|  |  | 464 | 3 | 47588462 | 50387742 | 0.88 | 0.32 | 1.00 | 0.78 | 0.10 | 1.00 | 3.92 x 10^-03^ |
|  |  | 963 | 6 | 32454578 | 32539567 | 0.12 | 0.04 | 0.20 | 0.01 | 0.00 | 0.04 | 4.07 x 10^-03^ |
|  |  | 954 | 6 | 29529756 | 29833843 | -0.74 | -1.00 | -0.19 | 0.55 | 0.04 | 1.00 | 1.50 x 10^-02^ |
|  |  | 2209 | 17 | 45883902 | 47516224 | 0.22 | 0.04 | 0.40 | 0.05 | 0.00 | 0.16 | 2.12 x 10^-02^ |
|  |  | 967 | 6 | 32682214 | 32897998 | -0.32 | -0.63 | -0.03 | 0.10 | 0.00 | 0.39 | 2.78 x 10^-02^ |
| AD (Jansen et al) | T2D (Xue et al) | 2351 | 19 | 45040933 | 45893307 | -0.76 | -1.00 | -0.52 | 0.59 | 0.27 | 1.00 | 1.01 x 10^-07^ |
|  |  | 964 | 6 | 32539568 | 32586784 | -1.00 | -1.00 | -0.90 | 1.00 | 0.81 | 1.00 | 1.22 x 10^-07^ |
|  |  | 965 | 6 | 32586785 | 32629239 | -1.00 | -1.00 | -0.79 | 1.00 | 0.62 | 1.00 | 2.13 x 10^-04^ |
|  |  | 2281 | 18 | 52512524 | 53762996 | 1.00 | 0.47 | 1.00 | 1.00 | 0.22 | 1.00 | 3.05 x 10^-04^ |
|  |  | 958 | 6 | 31106494 | 31250556 | -1.00 | -1.00 | -0.60 | 0.99 | 0.36 | 1.00 | 1.98 x 10^-03^ |
|  |  | 1209 | 7 | 1.3E+08 | 1.32E+08 | -0.58 | -1.00 | -0.23 | 0.34 | 0.05 | 1.00 | 2.15 x 10^-03^ |
|  |  | 967 | 6 | 32682214 | 32897998 | -0.62 | -1.00 | -0.26 | 0.38 | 0.07 | 1.00 | 2.47 x 10^-03^ |
|  |  | 957 | 6 | 30715007 | 31106493 | -0.86 | -1.00 | -0.38 | 0.75 | 0.14 | 1.00 | 5.88 x 10^-03^ |
|  |  | 961 | 6 | 31427210 | 32208901 | -0.63 | -1.00 | -0.14 | 0.40 | 0.02 | 1.00 | 1.95 x 10^-02^ |
|  |  | 464 | 3 | 47588462 | 50387742 | 0.82 | 0.19 | 1.00 | 0.67 | 0.04 | 1.00 | 2.08 x 10^-02^ |
| AD (Lambert et al) | T2D (Mahajan et al) | 2351 | 19 | 45040933 | 45893307 | -0.74 | -0.85 | -0.63 | 0.54 | 0.40 | 0.72 | 9.26 x 10^-30^ |
|  |  | 965 | 6 | 32586785 | 32629239 | -0.28 | -0.41 | -0.15 | 0.08 | 0.02 | 0.17 | 4.32 x 10^-05^ |
|  |  | 960 | 6 | 31320269 | 31427209 | -0.74 | -1.00 | -0.29 | 0.55 | 0.09 | 1.00 | 4.76 x 10^-03^ |
|  |  | 962 | 6 | 32208902 | 32454577 | -0.90 | -1.00 | -0.36 | 0.81 | 0.13 | 1.00 | 4.77 x 10^-03^ |
|  |  | 961 | 6 | 31427210 | 32208901 | -0.41 | -0.72 | -0.14 | 0.17 | 0.02 | 0.52 | 4.87 x 10^-03^ |
|  |  | 967 | 6 | 32682214 | 32897998 | -0.52 | -1.00 | -0.14 | 0.27 | 0.02 | 1.00 | 1.19 x 10^-02^ |
|  |  | 964 | 6 | 32539568 | 32586784 | -0.22 | -0.39 | -0.05 | 0.05 | 0.00 | 0.15 | 1.24 x 10^-02^ |
|  |  | 958 | 6 | 31106494 | 31250556 | -0.74 | -1.00 | -0.17 | 0.54 | 0.03 | 1.00 | 2.14 x 10^-02^ |
|  |  | 1126 | 7 | 27351287 | 28890886 | -0.31 | -0.66 | -0.03 | 0.09 | 0.00 | 0.44 | 3.41 x 10^-02^ |
| AD (Lambert et al) | T2D (Xue et al) | 2351 | 19 | 45040933 | 45893307 | -0.76 | -1.00 | -0.50 | 0.58 | 0.25 | 1.00 | 4.07 x 10^-06^ |
|  |  | 964 | 6 | 32539568 | 32586784 | -1.00 | -1.00 | -0.86 | 1.00 | 0.74 | 1.00 | 7.35 x 10^-04^ |
|  |  | 1126 | 7 | 27351287 | 28890886 | -0.51 | -1.00 | -0.16 | 0.26 | 0.03 | 1.00 | 4.85 x 10^-03^ |

Local genetic correlations between Alzheimer’s disease (AD) and type 2 diabetes (T2D) across genomic loci. The local genetic correlation coefficient (ρ) and shared variance (r²) quantify region-specific genetic sharing, with 95% confidence intervals and *p*-values testing ρ ≠ 0. Positive ρ indicates concordant effect directions within a locus, whereas negative ρ indicates opposing effects.

### Supplementary Table 7: Genomic loci showing evidence of local genetic sharing between AD and T2D

| **Locus** | **N analyses** | **N of loci completely/near completely shared** |
| --- | --- | --- |
| chr3:47.59–50.39 Mb | 2 | 2 |
| chr6:29.53–29.83 Mb | 1 | 1 |
| chr6:30.72–31.11 Mb | 1 | 1 |
| chr6:31.11–31.25 Mb | 2 | 2 |
| chr6:31.32–31.43 Mb | 2 | 1 |
| chr6:31.43–32.21 Mb | 2 | 1 |
| chr6:32.21–32.45 Mb | 2 | 1 |
| chr6:32.54–32.59 Mb | 3 | 2 |
| chr6:32.59–32.63 Mb | 3 | 1 |
| chr6:32.68–32.90 Mb | 3 | 2 |
| chr7:27.35–28.89 Mb | 2 | 1 |
| chr7:130.00–132.00 Mb | 1 | 1 |
| chr18:52.51–53.76 Mb | 1 | 1 |
| chr19:45.04–45.89 Mb | 4 | 2 |

N analyses indicate the number of AD–T2D GWAS pair analyses in which each locus exhibited significant local genetic correlation. N of loci completely/near completely shared indicates the number of analyses in which most local genetic variance at the locus was attributable to variants shared between AD and T2D, indicating strong regional genetic overlap.

### Supplementary Table 8: Genomic loci showing reproducible local genetic correlation between AD and T2D

| **locus** | **N analyses** | **direction** | **Rho (min)** | **Rho (max)** |
| --- | --- | --- | --- | --- |
| chr3:47.59–50.39 Mb | 2 | Positive | 0.82 | 0.88 |
| chr6:31.11–31.25 Mb | 2 | Negative | -1 | -0.74 |
| chr6:31.32–31.43 Mb | 2 | Negative | -0.74 | -0.53 |
| chr6:31.43–32.21 Mb | 2 | Negative | -0.63 | -0.41 |
| chr6:32.21–32.45 Mb | 2 | Negative | -0.9 | -0.37 |
| chr6:32.54–32.59 Mb | 3 | Negative | -1 | -0.22 |
| chr6:32.59–32.63 Mb | 3 | Negative | -1 | -0.14 |
| chr6:32.68–32.90 Mb | 3 | Negative | -0.62 | -0.32 |
| chr7:27.35–28.89 Mb | 2 | Negative | -0.51 | -0.31 |
| chr19:45.04–45.89 Mb | 4 | Negative | -0.76 | -0.61 |

Loci were included if they exhibited significant local genetic correlation in at least two independent AD–T2D GWAS analyses. *N analyses* indicate the number of analyses in which the locus was significant. *Direction* indicates whether local genetic effects were concordant or discordant between traits. The range of local genetic correlation estimates is summarised by ρ(min) and ρ(max).

### Supplementary Table 9: Bidirectional MR analyses between AD and T2D

| Exposure | Outcome | Method | OR (95%CI) | nIV | (P) pleiotropy * | P (heterogeneity)& | P# |
| --- | --- | --- | --- | --- | --- | --- | --- |
| AD (Jansen et al) | T2D (Mahajan et al) | IVW | 0.98 (0.86 – 1.13) | 22 | 4.97 x 10^-01^ | 9.76 x 10^-01^ | 8.05 x 10^-01^ |
|  |  | Weighted median | 0.96 (0.80 – 1.15) | 22 | 4.97 x 10^-01^ | 9.76 x 10^-01^ | 6.32 x 10^-01^ |
|  |  | MR Egger | 0.92 (0.73 – 1.16) | 22 | 4.97 x 10^-01^ | 9.76 x 10^-01^ | 4.89 x 10^-01^ |
|  |  | MR-PRESSO (Raw) | 0.98 (0.89 – 1.08) | 22 |  |  | 7.27 x 10^-01^ |
|  |  | MR-PRESSO (Corrected) |  | 22 |  |  |  |
| T2D (Mahajan et al) | AD (Jansen et al) | IVW | 1.01 (1.00 – 1.01) | 173 | 6.37 x 10^-01^ | 4.92 x 10^-01^ | 1.42 x 10^-02^ |
|  |  | Weighted median | 1.01 (1.00 – 1.02) | 173 | 6.37 x 10^-01^ | 4.92 x 10^-01^ | 5.23 x 10^-02^ |
|  |  | MR Egger | 1.00 (0.99 – 1.02) | 173 | 6.37 x 10^-01^ | 4.92 x 10^-01^ | 6.31 x 10^-01^ |
|  |  | MR-PRESSO (Raw) | 1.01 (1.00 – 1.01) | 173 |  |  | 1.52 x 10^-02^ |
|  |  | MR-PRESSO (Corrected) |  | 173 |  |  |  |
| AD (Jansen et al) | T2D (Xue et al) | IVW | 0.97 (0.76 – 1.23) | 15 | 1.98 x 10^-01^ | 5.74 x 10^-01^ | 7.91 x 10^-01^ |
|  |  | Weighted median | 0.91 (0.65 – 1.28) | 15 | 1.98 x 10^-01^ | 5.74 x 10^-01^ | 5.86 x 10^-01^ |
|  |  | MR Egger | 0.56 (0.24 – 1.28) | 15 | 1.98 x 10^-01^ | 5.74 x 10^-01^ | 1.92 x 10^-01^ |
|  |  | MR-PRESSO (Raw) | 0.97 (0.77 – 1.21) | 15 |  |  | 7.82 x 10^-01^ |
|  |  | MR-PRESSO (Corrected) |  | 15 |  |  |  |
| T2D (Xue et al) | AD (Jansen et al) | IVW | 1.00 (1.00 – 1.01) | 98 | 9.43 x 10^-01^ | 8.87 x 10^-01^ | 2.60 x 10^-01^ |
|  |  | Weighted median | 1.01 (1.00 – 1.02) | 98 | 9.43 x 10^-01^ | 8.87 x 10^-01^ | 1.47 x 10^-01^ |
|  |  | MR Egger | 1.00 (0.98 – 1.03) | 98 | 9.43 x 10^-01^ | 8.87 x 10^-01^ | 7.62 x 10^-01^ |
|  |  | MR-PRESSO (Raw) | 1.00 (1.00 – 1.01) | 98 |  |  | 2.19 x 10^-01^ |
|  |  | MR-PRESSO (Corrected) |  | 98 |  |  |  |

AD: Alzheimer’s disease, T2D: type 2 diabetes, IVW, inverse variance weighted, MR-PRESSO: Mendelian randomisation pleiotropy residual sum and outlier, P: P-value, OR: odds ratio, CI: confidence interval, nIV: number of instrumental variables, * MR-Egger intercept p-value, &: P-value for heterogeneity testing, # The p-value for each of the model.

### Supplementary Table 10: Genome-wide significant independent SNPs and loci shared by AD and T2D

| **Independent SNPs** | **Unique ID** | **Genomic**  **loci** | **Lead**  **SNPs** | **Individual GWAS P-value** | | **Meta-analysis** | **Binary effect P-value** | **M-value** | |
| --- | --- | --- | --- | --- | --- | --- | --- | --- | --- |
|  |  |  |  | **AD** | **T2D** | **P-value (RE2)** |  | **AD** | **T2D** |
| GWS AD and T2D Independent SNPs and loci in the GWAS meta-analysis | | | | | | | | | |
| rs1980496 | 6:32340070:C:T | 1 | rs9275095,  rs9271548 | 3.81 x 10^-08^ | 6.90 x 10^-13^ | 2.68 x 10^-17^ | 1.06 x 10^-12^ | 0.00 | 1.00 |
| rs2395163 | 6:32387809:C:T |  |  | 6.15 x 10^-09^ | 1.70 x 10^-14^ | 4.57 x 10^-19^ | 9.48 x 10^-14^ | 0.00 | 1.00 |
| rs9268838 | 6:32428715:A:G |  |  | 4.02 x 10^-08^ | 4.30 x 10^-14^ | 3.29 x 10^-18^ | 1.11 x 10^-13^ | 0.00 | 1.00 |
| rs482044 | 6:32576064:C:G |  |  | 2.03 X 10^-08^ | 5.70 x 10^-11^ | 2.12 x 10^-15^ | 3.09 x 10^-10^ | 0.00 | 1.00 |
| rs646984 | 6:32576592:A:C |  |  | 1.78 X 10^-08^ | 2.00 x 10^-10^ | 3.25 x 10^-15^ | 8.35 x 10^-10^ | 0.00 | 1.00 |
| rs532965 | 6:32577973:G:T |  |  | 1.42 X 10^-10^ | 1.80 x 10^-20^ | 2.77 x 10^-26^ | 1.49 x 10^-19^ | 0.00 | 1.00 |
| rs1846190 | 6:32583813:A:G |  |  | 8.86 X 10^-11^ | 2.30 x 10^-13^ | 5.51 x 10^-20^ | 1.13 x 10^-12^ | 0.00 | 1.00 |
| rs4335021 | 6:32386619:C:T |  |  | 4.29 X 10^-09^ | 2.10 x 10^-08^ | 1.03 x 10^-13^ | 1.46 x 10^-07^ | 0.62 | 0.38 |
| rs138002663 | 6:32473542:C:T |  |  | 1.52 X 10^-08^ | 1.50 x 10^-12^ | 3.12 x 10^-17^ | 5.00 x 10^-12^ | 0.00 | 1.00 |
| rs9271260 | 6:32580507:A:C |  |  | 3.12 X 10^-08^ | 1.30 x 10^-10^ | 3.27 x 10^-15^ | 2.54 x 10^-10^ | 0.00 | 1.00 |
| rs9271548 | 6:32590234:A:T |  |  | 4.74 X 10^-08^ | 3.80 x 10^-14^ | 2.26 x 10^-18^ | 6.34 x 10^-14^ | 0.00 | 1.00 |
| rs116457146 | 6:32578632:C:T |  |  | 1.30 X 10^-10^ | 1.50 x 10^-12^ | 4.24 x 10^-19^ | 2.03 x 10^-11^ | 0.00 | 1.00 |
| rs9275095 | 6:32649088:C:G |  |  | 2.65 X 10^-10^ | 9.00 x 10^-21^ | 3.83 x 10^-28^ | 9.77 x 10^-22^ | 0.00 | 1.00 |
| rs9275150 | 6:32651617:A:G |  |  | 2.28 X 10^-08^ | 6.50 x 10^-09^ | 1.26 x 10^-13^ | 8.01 x 10^-02^ | 0.06 | 0.94 |
| rs9275362 | 6:32667957:C:T |  |  | 7.24 X 10^-10^ | 1.10 x 10^-14^ | 2.41 x 10^-20^ | 3.99 x 10^-14^ | 0.00 | 1.00 |
| rs12972156 | 19:45387459:C:G | 2 | rs12972156 | 0 | 7.60 x 10^-15^ | 0.00E+00 | 0 | 1.00 | 0.00 |
| rs157582 | 19:45396219:C:T |  |  | 0 | 2.50 x 10^-10^ | 0.00E+00 | 0 | 1.00 | 0.00 |
| rs10119 | 19:45406673:A:G |  |  | 0 | 3.50 x 10^-09^ | 0.00E+00 | 0 | 1.00 | 0.00 |
| rs7256200 | 19:45415935:G:T |  |  | 0 | 1.70 x 10^-14^ | 0.00E+00 | 0 | 1.00 | 0.00 |
| rs5117 | 19:45418790:C:T |  |  | 0 | 1.40 x 10^-13^ | 0.00E+00 | 0 | 1.00 | 0.00 |
| rs4420638 | 19:45422946:A:G |  |  | 0 | 4.00 x 10^-16^ | 0.00E+00 | 0 | 1.00 | 0.00 |

AD: Alzheimer’s disease, GWAS: genome-wide association studies, T2D: type 2 diabetes, SNP: single-nucleotide polymorphism, RE2: Han–Eskin modified random-effects model implemented in the Metasoft GWAS meta-analysis. We applied LD clumping using a threshold of r² < 0.6 to define independent SNPs and further identified lead SNPs as those with r² < 0.1 relative to others in the same region. Genomic loci were defined as regions within ±250 kb of each lead SNP, and overlapping regions were collapsed into a single locus; hence, there can be more than one lead SNP in a locus.

### Supplementary Table 11: AD genome-wide significant SNPs and loci showing association with T2D

| **Independent SNPs** | **Unique ID** | **Genomic**  **loci** | **Lead**  **SNPs** | **Individual GWAS P-value** | | **Meta-analysis** | **Binary effect P-value** | **M-value** | |
| --- | --- | --- | --- | --- | --- | --- | --- | --- | --- |
|  |  |  |  | **AD** | **T2D** | **P-value (RE2)** |  | **AD** | **T2D** |
| rs1980496 | 6:32340070:C:T | 1 | rs9271548, rs9275095 | 3.81 × 10^-08^ | 6.90 × 10^-13^ | 2.68 × 10^-17^ | 1.06 × 10^-12^ | 0.00 | 1.00 |
| rs3806157 | 6:32373801:G:T |  |  | 2.01 × 10^-08^ | 6.80 × 10^-07^ | 9.57 × 10^-12^ | 8.31 × 10^-08^ | 0.90 | 0.10 |
| rs9268494 | 6:32375352:A:C |  |  | 4.41 × 10^-08^ | 8.30 × 10^-07^ | 2.10 × 10^-11^ | 2.59 × 10^-07^ | 0.80 | 0.20 |
| rs2395163 | 6:32387809:C:T |  |  | 6.15 × 10^-09^ | 1.70 × 10^-14^ | 4.57 × 10^-19^ | 9.48 × 10^-14^ | 0.00 | 1.00 |
| rs3135344 | 6:32395036:C:T |  |  | 5.79 × 10^-10^ | 6.70 × 10^-08^ | 5.35 × 10^-14^ | 1.85 × 10^-09^ | 0.98 | 0.02 |
| rs9268838 | 6:32428715:A:G |  |  | 4.02 × 10^-08^ | 4.30 × 10^-14^ | 3.29 × 10^-18^ | 1.11 × 10^-13^ | 0.00 | 1.00 |
| rs138002663 | 6:32473542:C:T |  |  | 1.52 × 10^-08^ | 1.50 × 10^-12^ | 3.12 × 10^-17^ | 5.00 × 10^-12^ | 0.00 | 1.00 |
| rs9270599 | 6:32561656:A:G |  |  | 4.88 × 10^-08^ | 3.00 × 10^-07^ | 8.20 × 10^-12^ | 1.71 × 10^-06^ | 0.53 | 0.47 |
| rs482044 | 6:32576064:C:G |  |  | 2.03 × 10^-08^ | 5.70 × 10^-11^ | 2.12 × 10^-15^ | 3.09 × 10^-10^ | 0.00 | 1.00 |
| rs646984 | 6:32576592:A:C |  |  | 1.78 × 10^-08^ | 2.00 × 10^-10^ | 3.25 × 10^-15^ | 8.35 × 10^-10^ | 0.00 | 1.00 |
| rs532965 | 6:32577973:G:T |  |  | 1.42 × 10^-10^ | 1.80 × 10^-20^ | 2.77 × 10^-26^ | 1.49 × 10^-19^ | 0.00 | 1.00 |
| rs116457146 | 6:32578632:C:T |  |  | 1.30 × 10^-10^ | 1.50 × 10^-12^ | 4.24 × 10^-19^ | 2.03 × 10^-11^ | 0.00 | 1.00 |
| rs9271260 | 6:32580507:A:C |  |  | 3.12 × 10^-08^ | 1.30 × 10^-10^ | 3.27 × 10^-15^ | 2.54 × 10^-10^ | 0.00 | 1.00 |
| rs1846190 | 6:32583813:A:G |  |  | 8.86 × 10^-11^ | 2.30 × 10^-13^ | 5.51 × 10^-20^ | 1.13 × 10^-12^ | 0.00 | 1.00 |
| rs9271548 | 6:32590234:A:T |  |  | 4.74 × 10^-08^ | 3.80 × 10^-14^ | 2.26 × 10^-18^ | 6.34 × 10^-14^ | 0.00 | 1.00 |
| rs9275095 | 6:32649088:C:G |  |  | 2.65 × 10^-10^ | 9.00 × 10^-21^ | 3.83 × 10^-28^ | 9.77 × 10^-22^ | 0.00 | 1.00 |
| rs9275150 | 6:32651617:A:G |  |  | 2.28 × 10^-08^ | 6.50 × 10^-09^ | 1.26 × 10^-13^ | 08.01 × 10^-2^ | 0.06 | 0.94 |
| rs9275362 | 6:32667957:C:T |  |  | 7.24 × 10^-10^ | 1.10 × 10^-14^ | 2.41 × 10^-20^ | 3.99 × 10^-14^ | 0.00 | 1.00 |
| rs11218343 | 11:121435587:C:T | 2 | rs11218343 | 8.12 × 10^-12^ | 8.90 × 10^-04^ | 9.60 × 10^-14^ | 1.62 × 10^-13^ | 1.00 | 1.00 |
| rs1790211 | 11:121451813:G:T |  |  | 1.91 × 10^-08^ | 8.90 × 10^-04^ | 3.97 × 10^-10^ | 7.68 × 10^-10^ | 1.00 | 0.99 |
| rs2927437 | 19:45241638:A:G | 3 | rs2927437, rs41289512, rs11666329, rs6859, rs157580, rs1081105, rs3925681, rs60049679 | 4.18 × 10^-56^ | 6.20 × 10^-04^ | 6.62 × 10^-56^ | 1.23 × 10^-55^ | 1.00 | 0.00 |
| rs28399664 | 19:45324756:C:G |  |  | 1.43 × 10^-24^ | 9.00 × 10^-06^ | 1.10 × 10^-25^ | 4.66 × 10^-24^ | 1.00 | 0.00 |
| rs10412413 | 19:45327309:C:T |  |  | 6.97 × 10^-115^ | 7.50 × 10^-05^ | 2.87 × 10^-115^ | 2.08 × 10^-114^ | 1.00 | 0.00 |
| rs41289512 | 19:45351516:C:G |  |  | 1.46 × 10^-278^ | 1.20 × 10^-07^ | 5.19 × 10^-281^ | 4.39 × 10^-278^ | 1.00 | 0.00 |
| rs1871047 | 19:45351746:A:G |  |  | 2.35 × 10^-22^ | 3.00 × 10^-05^ | 1.04 × 10^-23^ | 7.49 × 10^-22^ | 1.00 | 0.00 |
| rs12974942 | 19:45352487:A:T |  |  | 8.12 × 10^-54^ | 5.20 × 10^-04^ | 8.22 × 10^-54^ | 2.60 × 10^-53^ | 1.00 | 0.00 |
| rs11666329 | 19:45354296:A:G |  |  | 2.07 × 10^-49^ | 1.20 × 10^-06^ | 1.06 × 10^-51^ | 6.76 × 10^-49^ | 1.00 | 0.00 |
| rs73050205 | 19:45356464:A:T |  |  | 2.05 × 10^-135^ | 2.00 × 10^-06^ | 3.95 × 10^-137^ | 5.42 × 10^-135^ | 1.00 | 0.00 |
| rs6859 | 19:45382034:A:G |  |  | 4.92 × 10^-154^ | 1.20 × 10^-04^ | 5.66 × 10^-154^ | 1.36 × 10^-153^ | 1.00 | 0.00 |
| rs157580 | 19:45395266:A:G |  |  | 1.88 × 10^-221^ | 8.80 × 10^-04^ | 1.98 × 10^-220^ | 4.95 × 10^-221^ | 1.00 | 0.00 |
| rs1081105 | 19:45412955:A:C |  |  | 1.16 × 10^-232^ | 4.40 × 10^-05^ | 1.21 × 10^-232^ | 3.06 × 10^-232^ | 1.00 | 0.00 |
| rs439401 | 19:45414451:C:T |  |  | 7.70 × 10^-167^ | 4.90 × 10^-04^ | 2.49 × 10^-166^ | 2.81 × 10^-166^ | 1.00 | 0.00 |
| rs3925681 | 19:45421100:A:G |  |  | 2.09 × 10^-125^ | 5.90 × 10^-05^ | 1.45 × 10^-125^ | 6.51 × 10^-125^ | 1.00 | 0.00 |
| rs60049679 | 19:45429708:C:G |  |  | 8.28 × 10^-16^ | 2.20 × 10^-04^ | 1.09 × 10^-16^ | 2.70 × 10^-15^ | 1.00 | 0.00 |
| rs62118504 | 19:45734751:A:G | 4 | rs62118504 | 4.49 × 10^-13^ | 9.60 × 10^-05^ | 3.33 × 10^-14^ | 1.32 × 10^-12^ | 1.00 | 0.00 |

### Supplementary Table 12: T2D genome-wide significant SNPs and loci showing association with AD

| **Independent SNPs** | **Unique ID** | **Genomic**  **loci** | | **Lead**  **SNPs** | **Individual GWAS P-value** | | **Meta-analysis** | **Binary effect P-value** | **M-value** | |
| --- | --- | --- | --- | --- | --- | --- | --- | --- | --- | --- |
|  |  |  |  |  | **AD** | **T2D** | **P-value (RE2)** |  | **AD** | **T2D** |
| rs60290409 | 1:51404041:A:C | | 1 | rs60290409 | 3.50 × 10^-04^ | 9.70 × 10^-10^ | 7.75 × 10^-11^ | 9.71 × 10^-10^ | 0.01 | 1.00 |
| rs6432613 | 2:161145612:A:G | | 2 | rs6432613 | 4.72 × 10^-04^ | 1.30 × 10^-10^ | 2.77 × 10^-11^ | 1.93 × 10^-10^ | 0.00 | 1.00 |
| rs1994077 | 3:12117220:A:G | | 3 | rs1994077 | 7.82 × 10^-04^ | 2.70 × 10^-17^ | 6.41 × 10^-18^ | 2.48 × 10^-17^ | 0.00 | 1.00 |
| rs879779 | 4:185704199:A:G | | 4 | rs879779 | 7.60 × 10^-04^ | 4.10 × 10^-09^ | 2.90 × 10^-09^ | 1.68 × 10^-08^ | 0.00 | 1.00 |
| rs467367 | 5:55811346:A:C | | 5 | rs467367 | 7.43 × 10^-04^ | 4.30 × 10^-08^ | 6.96 × 10^-09^ | 2.53 × 10^-09^ | 0.07 | 1.00 |
| rs3130931 | 6:31134888:C:T | | 6 | rs3130931,  rs35840219 | 6.34 × 10^-05^ | 1.40 × 10^-12^ | 4.71 × 10^-14^ | 1.74 × 10^-12^ | 0.00 | 1.00 |
| rs35840219 | 6:31249267:C:G | |  |  | 1.91 × 10^-04^ | 4.10 × 10^-09^ | 2.72 × 10^-10^ | 5.77 × 10^-09^ | 0.00 | 1.00 |
| rs28381348 | 6:31708910:C:T | |  |  | 6.33 × 10^-04^ | 3.10 × 10^-08^ | 3.57 × 10^-09^ | 2.91 × 10^-08^ | 0.00 | 1.00 |
| rs4335021 | 6:32386619:C:T | | 7 | rs3129882,  rs111637026,  rs9271548,  rs9275095 | 4.29 × 10^-09^ | 2.10 × 10^-08^ | 1.03 × 10^-13^ | 1.46 × 10^-07^ | 0.62 | 0.38 |
| rs9268492 | 6:32375280:C:G | |  |  | 7.44 × 10^-09^ | 5.00 × 10^-09^ | 2.73 × 10^-14^ | 7.62 × 10^-1^ | 0.12 | 0.88 |
| rs116457146 | 6:32578632:C:T | |  |  | 1.30 × 10^-10^ | 1.50 × 10^-12^ | 4.24 × 10^-19^ | 2.03 × 10^-11^ | 0.00 | 1.00 |
| rs7743662 | 6:32425049:A:G | |  |  | 2.61 × 10^-06^ | 2.00 × 10^-08^ | 2.96 × 10^-11^ | 5.74 × 10^-08^ | 0.00 | 1.00 |
| rs1846190 | 6:32583813:A:G | |  |  | 8.86 × 10^-11^ | 2.30 × 10^-13^ | 5.51 × 10^-20^ | 1.13 × 10^-12^ | 0.00 | 1.00 |
| rs9275205 | 6:32657560:C:T | |  |  | 2.62 × 10^-08^ | 3.40 × 10^-11^ | 1.48 × 10^-15^ | 1.18 × 10^-10^ | 0.00 | 1.00 |
| rs1967688 | 6:32340068:C:T | |  |  | 4.74 × 10^-07^ | 1.70 × 10^-09^ | 6.04 × 10^-13^ | 4.06 × 10^-09^ | 0.00 | 1.00 |
| rs532965 | 6:32577973:G:T | |  |  | 1.42 × 10^-10^ | 1.80 × 10^-20^ | 2.77 × 10^-26^ | 1.49 × 10^-19^ | 0.00 | 1.00 |
| rs9275095 | 6:32649088:C:G | |  |  | 2.65 × 10^-10^ | 9.00 × 10^-21^ | 3.83 × 10^-28^ | 9.77 × 10^-22^ | 0.00 | 1.00 |
| rs9275362 | 6:32667957:C:T | |  |  | 7.24 × 10^-10^ | 1.10 × 10^-14^ | 2.41 × 10^-20^ | 3.99 × 10^-14^ | 0.00 | 1.00 |
| rs2395163 | 6:32387809:C:T | |  |  | 6.15 × 10^-09^ | 1.70 × 10^-14^ | 4.57 × 10^-19^ | 9.48 × 10^-14^ | 0.00 | 1.00 |
| rs138002663 | 6:32473542:C:T | |  |  | 1.52 × 10^-08^ | 1.50 × 10^-12^ | 3.12 × 10^-17^ | 5.00 × 10^-12^ | 0.00 | 1.00 |
| rs1980496 | 6:32340070:C:T | |  |  | 3.81 × 10^-08^ | 6.90 × 10^-13^ | 2.68 × 10^-17^ | 1.06 × 10^-12^ | 0.00 | 1.00 |
| rs9271548 | 6:32590234:A:T | |  |  | 4.74 × 10^-08^ | 3.80 × 10^-14^ | 2.26 × 10^-18^ | 6.34 × 10^-14^ | 0.00 | 1.00 |
| rs2858861 | 6:32580331:C:T | |  |  | 6.44 × 10^-08^ | 9.40 × 10^-12^ | 6.84 × 10^-16^ | 1.86 × 10^-11^ | 0.00 | 1.00 |
| rs9271375 | 6:32587067:A:G | |  |  | 1.22 × 10^-07^ | 3.20 × 10^-12^ | 3.28 × 10^-16^ | 4.30 × 10^-12^ | 0.00 | 1.00 |
| rs5026743 | 6:32439964:G:T | |  |  | 1.25 × 10^-07^ | 6.80 × 10^-13^ | 1.14 × 10^-16^ | 1.41 × 10^-12^ | 0.00 | 1.00 |
| rs9272546 | 6:32606941:C:T | |  |  | 1.96 × 10^-07^ | 5.70 × 10^-11^ | 2.03 × 10^-14^ | 2.06 × 10^-10^ | 0.00 | 1.00 |
| rs9275611 | 6:32683763:A:G | |  |  | 4.77 × 10^-07^ | 5.50 × 10^-15^ | 8.69 × 10^-18^ | 2.76 × 10^-14^ | 0.00 | 1.00 |
| rs17191234 | 6:32564681:A:C | |  |  | 6.58 × 10^-07^ | 5.40 × 10^-13^ | 4.79 × 10^-16^ | 1.30 × 10^-12^ | 0.00 | 1.00 |
| rs522308 | 6:32581922:C:T | |  |  | 2.49 × 10^-06^ | 1.10 × 10^-16^ | 2.50 × 10^-19^ | 1.28 × 10^-16^ | 0.00 | 1.00 |
| rs3104412 | 6:32585967:A:G | |  |  | 3.86 × 10^-06^ | 2.20 × 10^-11^ | 6.84 × 10^-14^ | 3.83 × 10^-11^ | 0.00 | 1.00 |
| rs9270949 | 6:32572975:C:T | |  |  | 1.23 × 10^-05^ | 1.30 × 10^-13^ | 8.34 × 10^-16^ | 2.17 × 10^-13^ | 0.00 | 1.00 |
| rs2273017 | 6:32337630:A:G | |  |  | 1.23 × 10^-05^ | 1.90 × 10^-13^ | 2.98 × 10^-15^ | 4.58 × 10^-13^ | 0.00 | 1.00 |
| rs6927022 | 6:32612397:A:G | |  |  | 1.36 × 10^-05^ | 2.40 × 10^-15^ | 2.40 × 10^-17^ | 5.16 × 10^-15^ | 0.00 | 1.00 |
| rs6910071 | 6:32282854:A:G | |  |  | 1.49 × 10^-05^ | 1.10 × 10^-11^ | 1.08 × 10^-13^ | 1.80 × 10^-11^ | 0.00 | 1.00 |
| rs35445101 | 6:32546879:A:G | |  |  | 1.80 × 10^-05^ | 1.30 × 10^-10^ | 1.76 × 10^-12^ | 4.89 × 10^-10^ | 0.00 | 1.00 |
| rs9272293 | 6:32603742:A:G | |  |  | 2.09 × 10^-05^ | 7.10 × 10^-11^ | 1.71 × 10^-12^ | 2.09 × 10^-10^ | 0.00 | 1.00 |
| rs9268045 | 6:32229655:C:T | |  |  | 3.04 × 10^-05^ | 8.20 × 10^-11^ | 1.26 × 10^-12^ | 1.12 × 10^-10^ | 0.00 | 1.00 |
| rs7751376 | 6:32605437:C:G | |  |  | 3.69 × 10^-05^ | 3.90 × 10^-14^ | 9.84 × 10^-16^ | 8.68 × 10^-14^ | 0.00 | 1.00 |
| rs4321864 | 6:32399187:A:C | |  |  | 3.92 × 10^-05^ | 1.10 × 10^-10^ | 2.91 × 10^-12^ | 1.99 × 10^-10^ | 0.00 | 1.00 |
| rs111365964 | 6:32517646:G:T | |  |  | 4.04 × 10^-05^ | 8.50 × 10^-13^ | 3.16 × 10^-14^ | 3.17 × 10^-12^ | 0.00 | 1.00 |
| rs9273508 | 6:32628439:A:G | |  |  | 4.66 × 10^-05^ | 4.80 × 10^-12^ | 2.67 × 10^-13^ | 1.40 × 10^-11^ | 0.00 | 1.00 |
| rs17843693 | 6:32622122:A:G | |  |  | 1.10 × 10^-04^ | 9.50 × 10^-09^ | 6.24 × 10^-10^ | 4.36 × 10^-08^ | 0.00 | 1.00 |
| rs9271364 | 6:32586787:A:G | |  |  | 2.03 × 10^-04^ | 1.40 × 10^-09^ | 1.21 × 10^-10^ | 2.22 × 10^-09^ | 0.00 | 1.00 |
| rs3129882 | 6:32409530:A:G | |  |  | 2.66 × 10^-04^ | 6.80 × 10^-13^ | 1.55 × 10^-13^ | 1.43 × 10^-12^ | 0.00 | 1.00 |
| rs111637026 | 6:32471012:A:G | |  |  | 2.77 × 10^-04^ | 1.90 × 10^-10^ | 2.64 × 10^-11^ | 5.94 × 10^-10^ | 0.00 | 1.00 |
| rs3129883 | 6:32410137:C:T | |  |  | 5.59 × 10^-04^ | 9.00 × 10^-13^ | 6.25 × 10^-13^ | 3.14 × 10^-12^ | 0.00 | 1.00 |
| rs3117125 | 6:32316016:G:T | |  |  | 6.27 × 10^-04^ | 2.90 × 10^-13^ | 2.09 × 10^-13^ | 9.12 × 10^-13^ | 0.00 | 1.00 |
| rs9273526 | 6:32628617:A:C | |  |  | 8.56 × 10^-04^ | 2.20 × 10^-11^ | 1.98 × 10^-11^ | 7.63 × 10^-11^ | 0.00 | 1.00 |
| rs9273368 | 6:32626475:A:G | |  |  | 6.87 × 10^-04^ | 2.80 × 10^-16^ | 2.70 × 10^-16^ | 7.64 × 10^-16^ | 0.00 | 1.00 |
| rs10808671 | 8:95967372:A:G | | 8 | rs10808671 | 3.79 × 10^-04^ | 1.70 × 10^-15^ | 3.36 × 10^-16^ | 1.79 × 10^-15^ | 0.00 | 1.00 |
| rs7911784 | 10:94139488:C:T | | 9 | rs7894946 | 1.10 × 10^-04^ | 1.80 × 10^-13^ | 1.31 × 10^-14^ | 2.66 × 10^-13^ | 0.00 | 1.00 |
| rs7084673 | 10:94167087:A:G | |  |  | 5.12 × 10^-04^ | 3.20 × 10^-12^ | 8.09 × 10^-13^ | 4.35 × 10^-12^ | 0.00 | 1.00 |
| rs7894946 | 10:94171083:A:G | |  |  | 8.33 × 10^-04^ | 5.50 × 10^-17^ | 3.61 × 10^-17^ | 8.42 × 10^-17^ | 0.00 | 1.00 |
| rs10510109 | 10:124120457:G:T | | 10 | rs2280141 | 1.93 × 10^-04^ | 3.40 × 10^-08^ | 1.29 × 10^-09^ | 6.59 × 10^-11^ | 0.17 | 1.00 |
| rs2280141 | 10:124193181:G:T | |  |  | 4.72 × 10^-05^ | 2.00 × 10^-13^ | 2.80 × 10^-15^ | 2.67 × 10^-13^ | 0.00 | 1.00 |
| rs35134156 | 15:77315432:A:G | | 11 | rs35134156 | 2.14 × 10^-04^ | 4.10 × 10^-10^ | 2.34 × 10^-11^ | 4.29 × 10^-10^ | 0.01 | 1.00 |
| rs3814877 | 16:30042677:G:T | | 12 | rs3814877 | 2.32 × 10^-06^ | 1.40 × 10^-10^ | 2.96 × 10^-13^ | 3.01 × 10^-10^ | 0.00 | 1.00 |
| rs8067439 | 17:17698254:A:G | | 13 | rs8067439 | 1.56 × 10^-04^ | 9.00 × 10^-10^ | 7.00 × 10^-11^ | 1.47 × 10^-09^ | 0.00 | 1.00 |
| rs112972879 | 19:46165082:A:G | | 14 | rs112972879 | 1.66 × 10^-05^ | 2.50 × 10^-17^ | 6.52 × 10^-19^ | 5.70 × 10^-17^ | 0.00 | 1.00 |

### Supplementary Table 13: SNPs and loci reaching genome-wide significance under cross-trait meta-analysis: potentially novel based on data

| **Independent SNPs** | **Unique ID** | **Genomic**  **loci** | **Lead**  **SNPs** | **Individual GWAS P-value** | | **Meta-analysis** | **Binary effect P-value** | **M-value** | | **Pleiotropy support** |
| --- | --- | --- | --- | --- | --- | --- | --- | --- | --- | --- |
|  |  |  |  | **AD** | **T2D** | **P-value (RE2)** |  | **AD** | **T2D** |  |
| rs72898910 | 1:50908098:C:G | 1 | rs72898910 | 3.06 × 10^-04^ | 9.30 × 10^-08^ | 9.36 × 10^-09^ | 4.81 × 10^-10^ | 0.16 | 1.00 | Ambiguous |
| rs10788930 | 1:51118253:C:T |  |  | 1.02 × 10^-04^ | 4.20 × 10^-07^ | 1.03 × 10^-08^ | 1.02 × 10^-07^ | 0.73 | 1.00 | Probable |
| rs268124 | 2:65654364:C:T | 2 | rs268124 | 2.01 × 10^-04^ | 2.20 × 10^-07^ | 7.33 × 10^-09^ | 4.91 × 10^-09^ | 0.40 | 1.00 | Ambiguous |
| rs34016387 | 5:105736791:A:G | 3 | rs34016387 | 1.32 × 10^-05^ | 2.40 × 10^-05^ | 2.39 × 10^-08^ | 9.21 × 10^-08^ | 1.00 | 1.00 | Yes |
| rs1265110 | 6:31119422:C:T | 4 | rs1265110 | 1.26 × 10^-05^ | 3.90 × 10^-06^ | 2.74 × 10^-08^ | 9.23 × 10^-01^ | 0.13 | 0.87 | Ambiguous |
| rs3806156 | 6:32373698:G:T | 5 | rs35366052, rs2647045 | 9.18 × 10^-08^ | 3.10 × 10^-06^ | 1.63 × 10^-10^ | 3.49 × 10^-07^ | 0.90 | 0.10 | Ambiguous |
| rs35366052 | 6:32531923:A:G |  |  | 8.69 × 10^-04^ | 8.60 × 10^-07^ | 2.44 × 10^-08^ | 7.57 × 10^-08^ | 0.97 | 1.00 | Yes |
| rs17843620 | 6:32620800:A:G |  |  | 5.11 × 10^-06^ | 8.20 × 10^-07^ | 1.84 × 10^-09^ | 3.41 × 10^-04^ | 0.11 | 0.89 | Ambiguous |
| rs2647045 | 6:32668100:A:G |  |  | 1.79 × 10^-07^ | 7.30 × 10^-08^ | 1.05 × 10^-11^ | 8.08 × 10^-01^ | 0.12 | 0.89 | Ambiguous |
| rs7089084 | 10:124115881:A:T | 6 | rs7089084 | 2.29 × 10^-04^ | 7.50 × 10^-08^ | 3.38 × 10^-09^ | 2.63 × 10^-10^ | 0.23 | 1.00 | Ambiguous |
| rs2231884 | 11:65656564:C:T | 7 | rs2231884 | 2.26 × 10^-06^ | 3.30 × 10^-06^ | 4.15 × 10^-09^ | 1.10 × 10^-03^ | 0.33 | 0.67 | Ambiguous |
| rs7131432 | 11:121426870:A:T | 8 | rs7131432 | 8.01 × 10^-08^ | 7.90 × 10^-04^ | 1.36 × 10^-09^ | 2.65 × 10^-09^ | 1.00 | 0.99 | Yes |
| rs12900395 | 15:77310345:C:G | 9 | rs12900395 | 4.53 × 10^-05^ | 1.00 × 10^-07^ | 1.06 × 10^-09^ | 1.65 × 10^-08^ | 0.73 | 1.00 | Probable |
| rs150775861 | 16:85757220:A:C | 10 | rs150775861 | 1.16 × 10^-06^ | 6.70 × 10^-04^ | 1.11 × 10^-08^ | 2.36 × 10^-08^ | 1.00 | 1.00 | Yes |
| rs4309 | 17:61559923:C:T | 11 | rs4311 | 3.94 × 10^-07^ | 1.50 × 10^-06^ | 3.38 × 10^-10^ | 1.76 × 10^-05^ | 0.52 | 0.48 | Ambiguous |
| rs4311 | 17:61560763:C:T |  |  | 1.78 × 10^-07^ | 4.50 × 10^-07^ | 3.66 × 10^-11^ | 2.76 × 10^-04^ | 0.32 | 0.68 | Ambiguous |
| rs7221678 | 17:61579612:C:T |  |  | 3.66 × 10^-06^ | 7.20 × 10^-06^ | 1.09 × 10^-08^ | 7.25 × 10^-04^ | 0.35 | 0.65 | Ambiguous |

### Supplementary Table 14: LDtraits novelty check for the four RE2-identified loci with effects in both AD and T2D

rs34016387, rs35366052, rs7131432 and rs150775861

| **Query** | **GWAS Trait** | **PMID** | **RS Number** | **Position (GRCh37)** | **Alleles** | **R2** | **D'** | **Risk Allele** | **Effect Size (95% CI)** | **Beta or OR** | **P value** |
| --- | --- | --- | --- | --- | --- | --- | --- | --- | --- | --- | --- |
| rs35366052 | Total fatty acids levels (UKB data field 23442) | 36764567 | rs35603463 | chr6:32531745 | C=0.644, T=0.356 | 0.86 | 0.97 | 0.43 | 0.03 | 0.024-0.046 | 8.00 × 10^-10^ |
| rs35366052 | Lymphocyte-to-monocyte ratio | 34469753 | rs35464393 | chr6:32530198 | C=0.655, T=0.345 | 0.83 | 0.98 | 0.17 | NA | NA | 2.00 × 10^-10^ |
| rs35366052 | Height | 36224396 | rs35464393 | chr6:32530198 | C=0.655, T=0.345 | 0.83 | 0.98 | 0.16 | 0.04 | 0.037-0.042 | 2.00 × 10^-180^ |
| rs35366052 | TREM2 protein levels | 39789286 | rs28796313 | chr6:32528538 | C=0.637, T=0.363 | 0.83 | 0.94 | 0.68 | 0.06 | 0.046-0.069 | 2.00 × 10^-25^ |
| rs35366052 | Complement factor B levels (CFB.4129.72.1) | 29875488 | rs9256938 | chr6:32547194 | A=0.661, C=0.339 | 0.76 | 0.95 | 0.74 | 0.23 | 0.17-0.29 | 2.00 × 10^-12^ |
| rs35366052 | F-cooking vegetables liking (derived food-liking factor) | 35585065 | rs28819133 | chr6:32527988 | C=0.381, T=0.619 | 0.74 | 0.86 | 0.69 | 0.07 | 0.045-0.092 | 1.00 × 10^-08^ |
| rs35366052 | Total lipids in small HDL (UKB data field 23573) | 36764567 | rs28646006 | chr6:32527793 | C=0.413, T=0.587 | 0.66 | 0.87 | 0.50 | 0.04 | 0.024-0.047 | 4.00 × 10^-10^ |
| rs150775861 | Color vision defects (Tritan) | 37359372 | rs191791258 | chr16:85763664 | C=0.024, G=0.976 | 0.62 | 1.00 | 0.98 | 0.16 | 0.093-0.233 | 7.00 × 10^-06^ |
| rs35366052 | CNTNAP4 protein levels | 39789286 | rs71534539 | chr6:32512754 | A=0.509, G=0.491 | 0.59 | 0.97 | 0.47 | 0.09 | 0.077-0.105 | 2.00 × 10^-42^ |
| rs35366052 | CSDE1 protein levels | 39789286 | rs71534539 | chr6:32512754 | A=0.509, G=0.491 | 0.59 | 0.97 | 0.47 | 0.09 | 0.074-0.101 | 7.00 × 10^-41^ |
| rs35366052 | STAU1 protein levels | 39789286 | rs71534539 | chr6:32512754 | A=0.509, G=0.491 | 0.59 | 0.97 | 0.47 | 0.07 | 0.052-0.08 | 1.00 × 10^-21^ |
| rs35366052 | STOML2 protein levels | 39789286 | rs71534539 | chr6:32512754 | A=0.509, G=0.491 | 0.59 | 0.97 | 0.47 | 0.05 | 0.036-0.064 | 5.00 × 10^-12^ |
| rs35366052 | UGDH protein levels | 39789286 | rs71534539 | chr6:32512754 | A=0.509, G=0.491 | 0.59 | 0.97 | 0.47 | 0.06 | 0.046-0.074 | 2.00 × 10^-17^ |
| rs35366052 | TXNL1 protein levels | 39789286 | rs71534539 | chr6:32512754 | A=0.509, G=0.491 | 0.59 | 0.97 | 0.47 | 0.09 | 0.08-0.107 | 4.00 × 10^-45^ |
| rs35366052 | GABARAPL1 protein levels | 39789286 | rs71534539 | chr6:32512754 | A=0.509, G=0.491 | 0.59 | 0.97 | 0.47 | 0.05 | 0.034-0.062 | 4.00 × 10^-12^ |
| rs35366052 | SPRING1 protein levels | 39789286 | rs71534539 | chr6:32512754 | A=0.509, G=0.491 | 0.59 | 0.97 | 0.47 | 0.07 | 0.056-0.083 | 3.00 × 10^-23^ |
| rs35366052 | LRP2BP protein levels | 39789286 | rs71534539 | chr6:32512754 | A=0.509, G=0.491 | 0.59 | 0.97 | 0.47 | 0.05 | 0.035-0.062 | 5.00 × 10^-12^ |
| rs35366052 | MYL6B protein levels | 39789286 | rs71534539 | chr6:32512754 | A=0.509, G=0.491 | 0.59 | 0.97 | 0.47 | 0.07 | 0.053-0.081 | 5.00 × 10^-24^ |
| rs35366052 | NUBP1 protein levels | 39789286 | rs71534539 | chr6:32512754 | A=0.509, G=0.491 | 0.59 | 0.97 | 0.47 | 0.07 | 0.057-0.085 | 2.00 × 10^-24^ |
| rs35366052 | PAFAH1B3 protein levels | 39789286 | rs71534539 | chr6:32512754 | A=0.509, G=0.491 | 0.59 | 0.97 | 0.47 | 0.11 | 0.099-0.126 | 7.00 × 10^-69^ |
| rs35366052 | MAPRE3 protein levels | 39789286 | rs71534539 | chr6:32512754 | A=0.509, G=0.491 | 0.59 | 0.97 | 0.47 | 0.10 | 0.09-0.117 | 7.00 × 10^-58^ |
| rs35366052 | MORF4L2 protein levels | 39789286 | rs71534539 | chr6:32512754 | A=0.509, G=0.491 | 0.59 | 0.97 | 0.47 | 0.07 | 0.053-0.081 | 4.00 × 10^-21^ |
| rs35366052 | NFATC3 protein levels | 39789286 | rs71534539 | chr6:32512754 | A=0.509, G=0.491 | 0.59 | 0.97 | 0.47 | 0.08 | 0.066-0.093 | 4.00 × 10^-36^ |
| rs35366052 | KIAA2013 protein levels | 39789286 | rs71534539 | chr6:32512754 | A=0.509, G=0.491 | 0.59 | 0.97 | 0.47 | 0.05 | 0.037-0.065 | 3.00 × 10^-13^ |
| rs35366052 | CLEC5A protein levels | 39789286 | rs71536575 | chr6:32531971 | A=0.689, G=0.311 | 0.51 | 0.83 | 0.77 | 0.06 | 0.051-0.078 | 2.00 × 10^-27^ |
| rs35366052 | RBPMS protein levels | 39789286 | rs66931410 | chr6:32518235 | C=0.535, T=0.465 | 0.45 | 0.80 | 0.49 | 0.06 | 0.05-0.076 | 1.00 × 10^-23^ |
| rs35366052 | CD4 on CD4+ T cell | 32929287 | rs28584364 | chr6:32512048 | C=0.476, T=0.524 | 0.41 | 0.78 | 0.57 | 0.36 | 0.26-0.47 | 5.00 × 10^-12^ |
| rs35366052 | CD4 on Central Memory CD4+ T cell | 32929287 | rs28584364 | chr6:32512048 | C=0.476, T=0.524 | 0.41 | 0.78 | 0.57 | 0.33 | 0.23-0.44 | 3.00 × 10^-10^ |
| rs35366052 | CD4 on naive CD4+ T cell | 32929287 | rs28584364 | chr6:32512048 | C=0.476, T=0.524 | 0.41 | 0.78 | 0.57 | 0.32 | 0.21-0.44 | 1.00 × 10^-08^ |
| rs35366052 | CD4 on CD45RA+ CD4+ T cell | 32929287 | rs28584364 | chr6:32512048 | C=0.476, T=0.524 | 0.41 | 0.78 | 0.57 | 0.35 | 0.25-0.46 | 2.00 × 10^-11^ |
| rs35366052 | Physical function (baseline) | 40374629 | rs67961283 | chr6:32512947 | A=0.77, G=0.23 | 0.40 | 0.91 | 0.23 | 0.02 | 0.018-0.023 | 1.00 × 10^-53^ |
| rs35366052 | Height (baseline) | 40374629 | rs67961283 | chr6:32512947 | A=0.77, G=0.23 | 0.40 | 0.91 | 0.23 | 0.03 | 0.026-0.032 | 2.00 × 10^-71^ |
| rs35366052 | TIMD4 protein levels | 39789286 | rs67961283 | chr6:32512947 | A=0.77, G=0.23 | 0.40 | 0.91 | 0.77 | 0.11 | 0.098-0.124 | 5.00 × 10^-77^ |

### Supplementary Table 15: Candidate shared loci for AD and T2D with evidence of distinct causal variants

| **chunk** | **chr** | **st** | **sp** | **Max-abs-Z-ADjan** | **Max-abs-Z-t2dmah** | **logBF4** | **PPA4** |
| --- | --- | --- | --- | --- | --- | --- | --- |
| 80 | 1 | 159913053 | 162346607 | 6.37 | 3.54 | 5.96 | 0.63 |
| 106 | 1 | 206080686 | 208410160 | 8.88 | 5.63 | 32.39 | 1.00 |
| 206 | 2 | 127374341 | 128033327 | 14.01 | 3.36 | 84.35 | 0.71 |
| 269 | 2 | 233550205 | 235150916 | 6.15 | 4.35 | 7.48 | 0.88 |
| 410 | 4 | 10699425 | 12322014 | 6.00 | 4.17 | 5.77 | 0.68 |
| 662 | 6 | 40345115 | 42038449 | 8.26 | 6.06 | 28.15 | 1.00 |
| 666 | 6 | 47311898 | 48391069 | 6.36 | 3.64 | 7.19 | 0.68 |
| 803 | 7 | 98716494 | 100196525 | 7.95 | 4.62 | 21.02 | 0.96 |
| 831 | 7 | 142656310 | 144968221 | 6.65 | 4.19 | 9.00 | 0.77 |
| 868 | 8 | 26682525 | 28162012 | 9.05 | 3.96 | 28.74 | 0.69 |
| 1019 | 10 | 10249396 | 12585768 | 5.72 | 11.84 | 61.27 | 0.59 |
| 1128 | 11 | 58780549 | 62222839 | 8.01 | 4.85 | 21.59 | 0.97 |
| 1141 | 11 | 84381474 | 86618929 | 8.79 | 5.00 | 31.07 | 1.00 |
| 1368 | 14 | 91296860 | 93132080 | 6.42 | 5.78 | 16.01 | 0.99 |
| 1404 | 15 | 58442090 | 59694003 | 6.08 | 4.09 | 7.01 | 0.79 |
| 1487 | 17 | 4696345 | 5741160 | 6.15 | 4.06 | 8.09 | 0.75 |
| 1510 | 17 | 45876022 | 47516523 | 5.64 | 7.97 | 24.03 | 0.63 |
| 1579 | 19 | 610729 | 2098096 | 6.53 | 4.63 | 10.78 | 0.97 |

AD: Alzheimer’s disease, T2D: type 2 diabetes, chr: chromosome, st: start position of the genomic region, sp: end position of the genomic region, chunk: approximately independent genomic region (LD block) defined for the GWAS-PW analysis, max-abs-Z-Adjan: maximum absolute signed Z-score observed among SNPs within the region for the AD GWAS (Jansen et al.), max-abs-Z-t2dmah: maximum absolute signed Z-score observed among SNPs within the region for the T2D GWAS (Mahajan et al.), logBF4: regional log Bayes factor supporting Model 4 (distinct causal variants influencing AD and T2D within the same genomic region) relative to alternative models, PPA4: regional posterior probability that the genomic region follows Model 4. Inference is made at the level of approximately independent genomic regions (LD blocks), and Model 4 indicates evidence for distinct causal variants affecting each trait within the same locus rather than a shared causal variant.

### Supplementary Table 16: Candidate shared loci for AD and T2D with evidence of the same causal variants

| **chunk** | **chr** | **st** | **sp** | **Max-abs-Z-ADjan** | **Max-abs-Z-t2dmah** | **logBF3** | **PPA3** |
| --- | --- | --- | --- | --- | --- | --- | --- |
| 656 | 6 | 31571971 | 32682590 | 6.51 | 9.70 | 52.66 | 0.97 |
| 657 | 6 | 32682664 | 33236268 | 5.09 | 7.76 | 27.91 | 0.99 |
| 1168 | 11 | 121175943 | 122591760 | 6.84 | 4.93 | 14.80 | 0.59 |
| 1452 | 16 | 29036613 | 31382470 | 5.50 | 6.46 | 17.92 | 0.57 |
| 1519 | 17 | 61545589 | 63147477 | 5.22 | 5.47 | 11.76 | 0.83 |
| 1608 | 19 | 44744147 | 46101600 | 52.52 | 8.70 | 1397.24 | 0.98 |
| 1649 | 20 | 54055895 | 56447520 | 6.21 | 3.95 | 8.49 | 0.67 |

AD: Alzheimer’s disease, T2D: type 2 diabetes, chr: chromosome, st: start position of the genomic region, sp: end position of the genomic region, chunk: approximately independent genomic region (LD block) defined for the GWAS-PW analysis, max-abs-Z-Adjan: maximum absolute signed Z-score observed among SNPs within the region for the AD GWAS (Jansen et al.), max-abs-Z-t2dmah: maximum absolute signed Z-score observed among SNPs within the region for the T2D GWAS (Mahajan et al.), logBF3: regional log Bayes factor supporting Model 3 (shared causal variant) relative to alternative models, PPA3: regional posterior probability that the genomic region follows Model 3. Genomic regions listed show evidence consistent with a shared causal configuration influencing both AD and T2D, with inference made at the regional rather than SNP level.

### Supplementary Table 17: Putative causal SNPs for AD and T2D

| **SNP** | **chr** | **pos** | **Z-ADjan** | **Z-t2dmah** | **logBF_3** | **PPA_3** | **chunk** |
| --- | --- | --- | --- | --- | --- | --- | --- |
| rs12140153 | 1 | 62579891 | -0.41 | -5.82 | 10.47 | 0.96 | 38 |
| rs1127215 | 1 | 117532790 | -2.02 | -7.34 | 21.37 | 0.95 | 71 |
| rs340874 | 1 | 214159256 | 1.20 | -10.63 | 49.42 | 0.96 | 109 |
| rs348330 | 1 | 229672955 | 0.18 | -7.61 | 21.38 | 1.00 | 118 |
| rs11680058 | 2 | 16574669 | 0.91 | 5.80 | 11.17 | 0.99 | 142 |
| rs1260326 | 2 | 27730940 | 0.21 | -10.31 | 45.42 | 0.91 | 148 |
| rs11688682 | 2 | 121347612 | 0.61 | -7.73 | 23.55 | 0.99 | 202 |
| rs4663105 | 2 | 127891427 | 14.01 | 0.17 | 90.52 | 1.00 | 206 |
| rs62271373 | 3 | 150066540 | -1.63 | 6.29 | 14.74 | 0.94 | 368 |
| rs362307 | 4 | 3241845 | 2.06 | 6.17 | 14.67 | 0.97 | 402 |
| rs190303429 | 4 | 20673790 | 4.83 | -0.41 | 8.49 | 0.91 | 417 |
| rs145710066 | 5 | 5285361 | 1.57 | 5.00 | 8.53 | 0.94 | 525 |
| rs146886108 | 5 | 14751305 | 0.25 | -8.09 | 26.33 | 0.94 | 531 |
| rs77372998 | 5 | 101250990 | -1.86 | -7.44 | 23.29 | 0.94 | 579 |
| rs116528032 | 6 | 2666633 | 4.59 | 2.22 | 8.65 | 0.94 | 633 |
| rs7756992 | 6 | 20679709 | -0.87 | -20.00 | 192.19 | 0.98 | 645 |
| rs187370608 | 6 | 40942196 | 8.26 | -1.22 | 30.11 | 0.94 | 662 |
| rs11759026 | 6 | 126792095 | 1.99 | -8.93 | 34.39 | 0.99 | 714 |
| rs4279506 | 7 | 23512896 | -1.29 | -5.91 | 10.72 | 0.95 | 763 |
| rs917195 | 7 | 30728452 | 0.37 | -6.62 | 14.70 | 0.97 | 766 |
| rs878521 | 7 | 44255643 | 0.13 | 7.70 | 22.23 | 0.96 | 774 |
| rs149364428 | 8 | 97737741 | 0.81 | 7.06 | 19.35 | 0.91 | 908 |
| rs10974438 | 9 | 4291928 | 0.94 | -7.73 | 22.68 | 0.94 | 939 |
| rs11257655 | 10 | 12307894 | -0.70 | 11.84 | 62.81 | 1.00 | 1019 |
| rs2237895 | 11 | 2857194 | -0.50 | -14.09 | 91.52 | 0.95 | 1096 |
| rs10830963 | 11 | 92708710 | -0.32 | -13.94 | 89.49 | 1.00 | 1146 |
| rs11218343 | 11 | 121435587 | -6.84 | 3.35 | 23.08 | 1.00 | 1168 |
| rs76895963 | 12 | 4384844 | -0.47 | 17.78 | 151.78 | 1.00 | 1182 |
| rs145834816 | 14 | 54385396 | -0.83 | 5.00 | 8.93 | 0.96 | 1345 |
| rs2925979 | 16 | 81534790 | 0.44 | 7.68 | 22.10 | 0.94 | 1473 |
| rs76726049 | 18 | 56189459 | 5.52 | -0.45 | 10.65 | 0.96 | 1560 |
| rs12454712 | 18 | 60845884 | -0.42 | 7.31 | 19.23 | 1.00 | 1564 |
| rs429358 | 19 | 45411941 | 52.46 | 8.70 | 1405.80 | 1.00 | 1608 |
| rs1800961 | 20 | 43042364 | 0.15 | 9.41 | 37.76 | 1.00 | 1644 |
| rs192366755 | 21 | 16413720 | 0.07 | 5.34 | 8.86 | 0.90 | 1656 |

AD: Alzheimer’s disease, T2D: type 2 diabetes, SNP: single-nucleotide polymorphism, chr: chromosome, pos: base-pair position (GRCh37), Z-Adjan: signed Z-score for AD GWAS (Jansen et al.), Z-t2dmah: signed Z-score for T2D GWAS (Mahajan et al.), logBF3: SNP-level log Bayes factor under Model 3 (shared causal variant) conditional on the genomic region, PPA3: SNP-level posterior probability under Model 3 conditional on the region and model, chunk: approximately independent genomic region (LD block) used in the GWAS-PW analysis. SNPs shown represent post hoc selected variants from regions with regional support for Model 3 and are presented as descriptive representatives rather than independent causal assignments.

### Supplementary Table 18: GWS genes shared by AD and T2D in gene-based cross-trait analysis

| **GENE** | **CHR** | **START** | **STOP** | **AD (Jansen et al)** | | **T2D (Mahajan et al)** | | **Stouffer's Method (equal weights)** | | | | **Stouffer's P-value < the individual GWAS p-value** |
| --- | --- | --- | --- | --- | --- | --- | --- | --- | --- | --- | --- | --- |
|  |  |  |  | **Sample size** | **P-value** | **Sample size** | **P-value** | **Z-score AD** | **Z-score T2D** | **Stouffer's Z-score** | **P-value** |  |
| APOC1 | 19 | 45417504 | 45422606 | 264790 | 3.1086 × 10^-15^ | 898130 | 9.6034 × 10^-15^ | -7.80 | -7.66 | -10.93 | 4.21 × 10^-28^ | Yes |
| APOE | 19 | 45409011 | 45412650 | 409171 | 2.5591 × 10^-14^ | 898130 | 1.4296 × 10^-11^ | -7.53 | -6.65 | -10.03 | 5.71 × 10^-24^ | Yes |
| TOMM40 | 19 | 45393826 | 45406946 | 385101 | 7.1998 × 10^-14^ | 898130 | 1.1538 × 10^-10^ | -7.39 | -6.34 | -9.71 | 1.37 × 10^-22^ | Yes |
| ACE | 17 | 61554422 | 61599205 | 376655 | 9.0992 × 10^-07^ | 898130 | 1.4969 × 10^-06^ | -4.77 | -4.67 | -6.68 | 1.21 × 10^-11^ | Yes |
| ACE | 17 | 61562184 | 61599209 | 381061 | 2.1674 × 10^-06^ | 898130 | 1.7542 × 10^-06^ | -4.59 | -4.64 | -6.53 | 3.31 × 10^-11^ | Yes |

*AD: Alzheimer’s disease, GWAS: genome-wide association studies, T2D: type 2 diabetes*

*Stouffer’s Z-scores were calculated using equal weights across studies to avoid dominance of the larger T2D GWAS sample size.*

*Multiple entries for ACE reflect distinct gene definitions or transcript models used in the gene-based association analysis.*

*Genome-wide gene-based significance: P_gene_ < 2.64 × 10⁻⁶ (Bonferroni correction for 18,965 tested genes).*

*Gene-based Z-score signs summarise aggregate evidence and should not be interpreted as definitive per-variant directional concordance; locus-level signed genetic covariance is assessed with LAVA.*

### Supplementary Table 19: T2D genome-wide significant genes showing evidence of association with AD

| **GENE** | **CHR** | **START** | **STOP** | **AD (Jansen et al)** | | **T2D (Mahajan et al)** | | **Stouffer's Method (equal weights)** | | | | **Stouffer's P-value < the individual GWAS p-value** |
| --- | --- | --- | --- | --- | --- | --- | --- | --- | --- | --- | --- | --- |
|  |  |  |  | **Sample size** | **P-value** | **Sample size** | **P-value** | **Z-score AD** | **Z-score T2D** | **Stouffer's Z-score** | **P-value** |  |
| FAF1 | 1 | 50905150 | 51425935 | 396417 | 1.00 × 10^-03^ | 898130 | 3.74 × 10^-10^ | -3.09 | -6.16 | -6.54 | 3.13 × 10^-11^ | Yes |
| CDKN2C | 1 | 51426417 | 51440305 | 399904 | 4.74 × 10^-03^ | 898130 | 7.37 × 10^-09^ | -2.59 | -5.66 | -5.84 | 2.62 × 10^-09^ | Yes |
| RBBP5 | 1 | 205055270 | 205091143 | 413345 | 1.98 × 10^-02^ | 898130 | 1.27 × 10^-06^ | -2.06 | -4.70 | -4.78 | 8.71 × 10^-07^ | Yes |
| RBMS1 | 2 | 161128662 | 161350305 | 408885 | 2.45 × 10^-03^ | 898130 | 2.66 × 10^-10^ | -2.81 | -6.21 | -6.38 | 8.83 × 10^-11^ | Yes |
| RNF123 | 3 | 49726932 | 49758962 | 408596 | 1.06 × 10^-02^ | 898130 | 1.19 × 10^-06^ | -2.30 | -4.72 | -4.96 | 3.44 × 10^-07^ | Yes |
| IP6K1 | 3 | 49761727 | 49823975 | 406105 | 9.64 × 10^-03^ | 898130 | 3.84 × 10^-07^ | -2.34 | -4.94 | -5.15 | 1.30 × 10^-07^ | Yes |
| CDHR4 | 3 | 49828165 | 49837268 | 406665 | 1.66 × 10^-02^ | 898130 | 2.62 × 10^-06^ | -2.13 | -4.55 | -4.73 | 1.14 × 10^-06^ | Yes |
| TRAIP | 3 | 49866034 | 49894007 | 411208 | 6.73 × 10^-03^ | 898130 | 9.93 × 10^-08^ | -2.47 | -5.20 | -5.42 | 2.90 × 10^-08^ | Yes |
| PCGF3 | 4 | 699537 | 764428 | 402965 | 1.43 × 10^-02^ | 898130 | 6.24 × 10^-08^ | -2.19 | -5.29 | -5.29 | 6.24 × 10^-08^ | Yes |
| HCG27 | 6 | 31165537 | 31171745 | 419639 | 5.83 × 10^-04^ | 898130 | 5.82 × 10^-08^ | -3.25 | -5.30 | -6.04 | 7.56 × 10^-10^ | Yes |
| CSNK2B | 6 | 31633013 | 31638120 | 422604 | 1.36 × 10^-02^ | 898130 | 1.53 × 10^-06^ | -2.21 | -4.67 | -4.86 | 5.82 × 10^-07^ | Yes |
| CSNK2B-LY6G5B-1181 | 6 | 31633879 | 31641323 | 410400 | 5.21 × 10^-03^ | 898130 | 5.19 × 10^-09^ | -2.56 | -5.72 | -5.86 | 2.33 × 10^-09^ | Yes |
| LY6G5C | 6 | 31644461 | 31651817 | 402277 | 1.18 × 10^-02^ | 898130 | 1.70 × 10^-06^ | -2.26 | -4.65 | -4.89 | 5.17 × 10^-07^ | Yes |
| MSH5 | 6 | 31707725 | 31732622 | 401965 | 2.00 × 10^-02^ | 898130 | 7.64 × 10^-07^ | -2.05 | -4.81 | -4.85 | 6.10 × 10^-07^ | Yes |
| C6orf10 | 6 | 32256303 | 32339684 | 414158 | 2.24 × 10^-05^ | 898130 | 2.90 × 10^-11^ | -4.08 | -6.55 | -7.52 | 2.82 × 10^-14^ | Yes |
| HLA-DRA | 6 | 32407619 | 32412823 | 414676 | 4.98 × 10^-06^ | 898130 | 1.79 × 10^-12^ | -4.42 | -6.95 | -8.04 | 4.47 × 10^-16^ | Yes |
| HLA-DRB5 | 6 | 32485120 | 32498064 | 147805 | 1.28 × 10^-02^ | 898130 | 1.64 × 10^-06^ | -2.23 | -4.65 | -4.87 | 5.61 × 10^-07^ | Yes |
| HLA-DRB1 | 6 | 32546546 | 32557625 | 249318 | 3.68 × 10^-06^ | 898130 | 5.44 × 10^-15^ | -4.48 | -7.73 | -8.63 | 2.94 × 10^-18^ | Yes |
| HLA-DQA1 | 6 | 32595956 | 32614839 | 275452 | 9.41 × 10^-05^ | 898130 | 8.14 × 10^-10^ | -3.73 | -6.03 | -6.91 | 2.51 × 10^-12^ | Yes |
| HLA-DQB1 | 6 | 32627244 | 32636160 | 203488 | 1.51 × 10^-04^ | 898130 | 1.74 × 10^-13^ | -3.61 | -7.27 | -7.70 | 6.86 × 10^-15^ | Yes |
| RELN | 7 | 103112231 | 103629963 | 405573 | 1.25 × 10^-02^ | 898130 | 1.15 × 10^-06^ | -2.24 | -4.72 | -4.93 | 4.21 × 10^-07^ | Yes |
| PINX1 | 8 | 10622473 | 10697394 | 409441 | 6.71 × 10^-03^ | 898130 | 3.29 × 10^-09^ | -2.47 | -5.80 | -5.85 | 2.45 × 10^-09^ | Yes |
| TP53INP1 | 8 | 95938200 | 95961639 | 407892 | 1.90 × 10^-03^ | 898130 | 2.84 × 10^-10^ | -2.89 | -6.20 | -6.43 | 6.37 × 10^-11^ | Yes |
| PLEKHA1 | 10 | 124134212 | 124191867 | 403133 | 9.19 × 10^-05^ | 898130 | 6.92 × 10^-13^ | -3.74 | -7.09 | -7.66 | 9.66 × 10^-15^ | Yes |
| IGF2 | 11 | 2150342 | 2170833 | 376975 | 2.55 × 10^-03^ | 898130 | 2.25 × 10^-08^ | -2.80 | -5.47 | -5.85 | 2.49 × 10^-09^ | Yes |
| INS-IGF2 | 11 | 2153768 | 2182439 | 375483 | 1.16 × 10^-02^ | 898130 | 6.58 × 10^-07^ | -2.27 | -4.84 | -5.03 | 2.51 × 10^-07^ | Yes |
| ZNF664 | 12 | 124456392 | 124499986 | 391247 | 1.81 × 10^-03^ | 898130 | 7.25 × 10^-07^ | -2.91 | -4.82 | -5.46 | 2.32 × 10^-08^ | Yes |
| PSTPIP1 | 15 | 77285700 | 77329673 | 382177 | 3.62 × 10^-03^ | 898130 | 5.58 × 10^-07^ | -2.69 | -4.87 | -5.34 | 4.58 × 10^-08^ | Yes |
| TMEM219 | 16 | 29952206 | 29984373 | 396486 | 6.47 × 10^-05^ | 898130 | 2.30 × 10^-09^ | -3.83 | -5.86 | -6.85 | 3.68 × 10^-12^ | Yes |
| TAOK2 | 16 | 29984962 | 30003582 | 402616 | 5.07 × 10^-05^ | 898130 | 2.18 × 10^-09^ | -3.89 | -5.87 | -6.90 | 2.62 × 10^-12^ | Yes |
| INO80E | 16 | 30006615 | 30017114 | 410489 | 3.80 × 10^-04^ | 898130 | 7.09 × 10^-08^ | -3.37 | -5.26 | -6.10 | 5.22 × 10^-10^ | Yes |
| DOC2A | 16 | 30016830 | 30034591 | 403079 | 4.98 × 10+ | 898130 | 2.84 × 10^-10^ | -4.42 | -6.20 | -7.51 | 3.01 × 10^-14^ | Yes |
| FAM57B | 16 | 30035748 | 30064299 | 399373 | 1.75 × 10^-05^ | 898130 | 2.23 × 10^-09^ | -4.14 | -5.87 | -7.07 | 7.49 × 10^-13^ | Yes |
| ALDOA | 16 | 30064411 | 30081778 | 380437 | 4.52 × 10^-04^ | 898130 | 2.11 × 10^-08^ | -3.32 | -5.48 | -6.22 | 2.44 × 10^-10^ | Yes |
| PPP4C | 16 | 30087299 | 30096698 | 406595 | 2.87 × 10^-04^ | 898130 | 4.70 × 10^-08^ | -3.44 | -5.34 | -6.21 | 2.66 × 10^-10^ | Yes |
| YPEL3 | 16 | 30103635 | 30108236 | 311498 | 3.67 × 10^-03^ | 898130 | 3.55 × 10^-07^ | -2.68 | -4.96 | -5.40 | 3.30 × 10^-08^ | Yes |
| GDPD3 | 16 | 30116131 | 30125177 | 278400 | 4.51 × 10^-03^ | 898130 | 1.47 × 10^-06^ | -2.61 | -4.67 | -5.15 | 1.29 × 10^-07^ | Yes |
| SREBF1 | 17 | 17713713 | 17740325 | 377136 | 1.21 × 10^-03^ | 898130 | 3.80 × 10^-09^ | -3.03 | -5.78 | -6.23 | 2.34 × 10^-10^ | Yes |
| TOM1L2 | 17 | 17746828 | 17875736 | 388189 | 1.28 × 10^-03^ | 898130 | 1.62 × 10^-09^ | -3.02 | -5.92 | -6.32 | 1.32 × 10^-10^ | Yes |
| GID4 | 17 | 17942606 | 17971718 | 407484 | 4.36 × 10^-04^ | 898130 | 1.77 × 10^-06^ | -3.33 | -4.64 | -5.63 | 8.89 × 10^-09^ | Yes |
| MLX | 17 | 40719086 | 40725257 | 414314 | 6.81 × 10^-03^ | 898130 | 1.45 × 10^-08^ | -2.47 | -5.55 | -5.67 | 7.26 × 10^-09^ | Yes |
| ACE | 17 | 61554422 | 61599205 | 376655 | 9.10 × 10^-07^ | 898130 | 1.50 × 10^-06^ | -4.77 | -4.67 | -6.68 | 1.21 × 10^-11^ | Yes |
| ACE | 17 | 61562184 | 61599209 | 381061 | 2.17 × 10^-06^ | 898130 | 1.75 × 10^-06^ | -4.59 | -4.64 | -6.53 | 3.31 × 10^-11^ | Yes |
| BPTF | 17 | 65821640 | 65980494 | 369763 | 3.36 × 10^-03^ | 898130 | 4.34 × 10^-09^ | -2.71 | -5.75 | -5.99 | 1.08 × 10^-09^ | Yes |
| KPNA2 | 17 | 66031635 | 66042958 | 407495 | 1.23 × 10^-02^ | 898130 | 5.63 × 10^-07^ | -2.25 | -4.87 | -5.03 | 2.43 × 10^-07^ | Yes |
| TOMM40 | 19 | 45393826 | 45406946 | 385101 | 7.20 × 10^-14^ | 898130 | 1.15 × 10^-10^ | -7.39 | -6.34 | -9.71 | 1.37 × 10^-22^ | Yes |
| APOE | 19 | 45409011 | 45412650 | 409171 | 2.56 × 10^-14^ | 898130 | 1.43 × 10^-11^ | -7.53 | -6.65 | -10.03 | 5.71 × 10^-24^ | Yes |
| APOC1 | 19 | 45417504 | 45422606 | 264790 | 3.11 × 10^-15^ | 898130 | 9.60 × 10^-15^ | -7.80 | -7.66 | -10.93 | 4.21 × 10^-28^ | Yes |
| EML2 | 19 | 46110252 | 46148887 | 378961 | 1.57 × 10^-03^ | 898130 | 3.43 × 10^-09^ | -2.95 | -5.79 | -6.19 | 3.10 × 10^-10^ | Yes |
| QPCTL | 19 | 46195741 | 46207247 | 378193 | 1.18 × 10^-03^ | 898130 | 5.53 × 10^-09^ | -3.04 | -5.71 | -6.19 | 3.00 × 10^-10^ | Yes |
| FBXO46 | 19 | 46213887 | 46234162 | 368419 | 2.97 × 10^-04^ | 898130 | 1.22 × 10^-06^ | -3.43 | -4.71 | -5.76 | 4.17 × 10^-09^ | Yes |

*AD: Alzheimer’s disease, GWAS: genome-wide association studies, T2D: type 2 diabetes*

*Stouffer’s Z-scores were calculated using equal weights across studies to avoid dominance of the larger T2D GWAS sample size.*

*Multiple entries for ACE reflect distinct gene definitions or transcript models used in the gene-based association analysis.*

*Genome-wide gene-based significance: P_gene_ < 2.64 × 10⁻⁶ (Bonferroni correction for 18,965 tested genes).*

*Gene-based Z-score signs summarise aggregate evidence and should not be interpreted as definitive per-variant directional concordance; locus-level signed genetic covariance is assessed with LAVA.*

### Supplementary Table 20: Genes reaching genome-wide significance for AD and T2D in the gene-based cross-trait analysis

| **GENE** | **CHR** | **START** | **STOP** | **AD (Jansen et al)** | | **T2D (Mahajan et al)** | | **Stouffer's Method (equal weights)** | | | | **Stouffer's P-value < the individual GWAS p-value** |
| --- | --- | --- | --- | --- | --- | --- | --- | --- | --- | --- | --- | --- |
|  |  |  |  | **Sample size** | **P-value** | **Sample size** | **P-value** | **Z-score AD** | **Z-score T2D** | **Stouffer's Z-score** | **P-value** |  |
| PUM1 | 1 | 31404353 | 31538838 | 393455 | 6.31 × 10^-03^ | 898130 | 4.10 × 10^-06^ | -2.49 | -4.46 | -4.92 | 4.38 × 10^-07^ | Yes |
| UBA7 | 3 | 49842640 | 49851379 | 391709 | 1.05 × 10^-02^ | 898130 | 6.90 × 10^-06^ | -2.31 | -4.35 | -4.71 | 1.26 × 10^-06^ | Yes |
| MFAP3 | 5 | 153418466 | 153600038 | 411742 | 2.89 × 10^-03^ | 898130 | 3.74 × 10^-05^ | -2.76 | -3.96 | -4.75 | 1.01 × 10^-06^ | Yes |
| C6orf47 | 6 | 31626075 | 31628549 | 389070 | 6.28 × 10^-03^ | 898130 | 5.96 × 10^-06^ | -2.50 | -4.38 | -4.86 | 5.83 × 10^-07^ | Yes |
| CLIC1 | 6 | 31698358 | 31707540 | 393533 | 7.00 × 10^-04^ | 898130 | 1.33 × 10^-05^ | -3.19 | -4.20 | -5.23 | 8.48 × 10^-08^ | Yes |
| MSH5-SAPCD1 | 6 | 31707797 | 31732628 | 401580 | 1.92 × 10^-02^ | 898130 | 3.18 × 10^-06^ | -2.07 | -4.51 | -4.66 | 1.62 × 10^-06^ | Yes |
| CYP21A2 | 6 | 32006042 | 32009447 | 255457 | 1.65 × 10^-03^ | 898130 | 1.87 × 10^-05^ | -2.94 | -4.12 | -4.99 | 2.98 × 10^-07^ | Yes |
| TAP2 | 6 | 32781544 | 32806599 | 413179 | 2.01 × 10^-03^ | 898130 | 1.27 × 10^-05^ | -2.88 | -4.21 | -5.01 | 2.71 × 10^-07^ | Yes |
| WIPI2 | 7 | 5229819 | 5273457 | 402938 | 9.18 × 10^-06^ | 898130 | 2.12 × 10^-03^ | -4.28 | -2.86 | -5.05 | 2.19 × 10^-07^ | Yes |
| TMEM106B | 7 | 12250867 | 12282993 | 413040 | 1.16 × 10^-04^ | 898130 | 4.99 × 10^-04^ | -3.68 | -3.29 | -4.93 | 4.10 × 10^-07^ | Yes |
| MAPK8IP1 | 11 | 45907202 | 45928016 | 363263 | 8.57 × 10^-04^ | 898130 | 1.14 × 10^-04^ | -3.14 | -3.69 | -4.82 | 7.04 × 10^-07^ | Yes |
| SPI1 | 11 | 47376411 | 47400127 | 386033 | 1.16 × 10^-03^ | 898130 | 1.71 × 10^-05^ | -3.05 | -4.14 | -5.08 | 1.86 × 10^-07^ | Yes |
| SLC39A13 | 11 | 47428683 | 47438047 | 406250 | 1.13 × 10^-04^ | 898130 | 2.41 × 10^-05^ | -3.69 | -4.06 | -5.48 | 2.11 × 10^-08^ | Yes |
| FIBP | 11 | 65651212 | 65656010 | 410183 | 4.40 × 10^-06^ | 898130 | 5.32 × 10^-06^ | -4.44 | -4.40 | -6.26 | 1.96 × 10^-10^ | Yes |
| FOSL1 | 11 | 65659520 | 65668044 | 386740 | 1.41 × 10^-04^ | 898130 | 1.83 × 10^-04^ | -3.63 | -3.56 | -5.09 | 1.81 × 10^-07^ | Yes |
| METAP2 | 12 | 95867296 | 95909615 | 399933 | 2.41 × 10^-03^ | 898130 | 3.10 × 10^-05^ | -2.82 | -4.01 | -4.83 | 6.99 × 10^-07^ | Yes |
| USP44 | 12 | 95910336 | 95945266 | 401965 | 1.05 × 10^-02^ | 898130 | 2.76 × 10^-06^ | -2.31 | -4.54 | -4.84 | 6.34 × 10^-07^ | Yes |
| CCDC92 | 12 | 124403207 | 124457378 | 399234 | 4.25 × 10^-03^ | 898130 | 3.12 × 10^-06^ | -2.63 | -4.52 | -5.06 | 2.14 × 10^-07^ | Yes |
| DNAH10OS | 12 | 124410971 | 124419531 | 386095 | 1.28 × 10^-02^ | 898130 | 8.20 × 10^-06^ | -2.23 | -4.31 | -4.63 | 1.87 × 10^-06^ | Yes |
| ZNF839 | 14 | 102783714 | 102809044 | 409911 | 3.17 × 10^-03^ | 898130 | 3.79 × 10^-06^ | -2.73 | -4.48 | -5.10 | 1.74 × 10^-07^ | Yes |
| HBQ1 | 16 | 230452 | 231180 | 385590 | 3.00 × 10^-03^ | 898130 | 6.51 × 10^-06^ | -2.75 | -4.36 | -5.03 | 2.50 × 10^-07^ | Yes |
| ASPHD1 | 16 | 29911696 | 29931185 | 399102 | 5.97 × 10^-04^ | 898130 | 6.72 × 10^-05^ | -3.24 | -3.82 | -4.99 | 3.00 × 10^-07^ | Yes |
| KCTD13 | 16 | 29916333 | 29938356 | 403322 | 6.95 × 10^-04^ | 898130 | 1.23 × 10^-05^ | -3.20 | -4.22 | -5.24 | 7.86 × 10^-08^ | Yes |
| C16orf92 | 16 | 30034655 | 30039057 | 406693 | 6.56 × 10^-03^ | 898130 | 3.92 × 10^-06^ | -2.48 | -4.47 | -4.91 | 4.46 × 10^-07^ | Yes |
| STX4 | 16 | 31044210 | 31054296 | 387065 | 6.60 × 10^-06^ | 898130 | 9.48 × 10^-03^ | -4.36 | -2.35 | -4.74 | 1.07 × 10^-06^ | Yes |
| ZNF668 | 16 | 31072164 | 31085641 | 400252 | 7.67 × 10^-06^ | 898130 | 6.00 × 10^-03^ | -4.32 | -2.51 | -4.83 | 6.70 × 10^-07^ | Yes |
| ZNF646 | 16 | 31085743 | 31095517 | 373507 | 5.63 × 10^-06^ | 898130 | 1.55 × 10^-02^ | -4.39 | -2.16 | -4.63 | 1.82 × 10^-06^ | Yes |
| IL34 | 16 | 70613798 | 70694585 | 377550 | 4.35 × 10^-06^ | 898130 | 1.53 × 10^-03^ | -4.45 | -2.96 | -5.24 | 8.05 × 10^-08^ | Yes |
| PLCG2 | 16 | 81772702 | 81991899 | 397764 | 5.15 × 10^-06^ | 898130 | 4.54 × 10^-03^ | -4.41 | -2.61 | -4.96 | 3.45 × 10^-07^ | Yes |
| CHRNE | 17 | 4801069 | 4806369 | 384076 | 4.37 × 10^-04^ | 898130 | 3.24 × 10^-04^ | -3.33 | -3.41 | -4.77 | 9.43 × 10^-07^ | Yes |
| C17orf107 | 17 | 4802713 | 4806227 | 375966 | 8.48 × 10^-05^ | 898130 | 1.21 × 10^-03^ | -3.76 | -3.03 | -4.80 | 7.78 × 10^-07^ | Yes |
| CAMTA2 | 17 | 4871287 | 4890960 | 399963 | 2.39 × 10^-04^ | 898130 | 2.49 × 10^-04^ | -3.49 | -3.48 | -4.93 | 4.08 × 10^-07^ | Yes |
| LRRC48 | 17 | 17876127 | 17920203 | 395918 | 4.94 × 10^-04^ | 898130 | 2.80 × 10^-06^ | -3.29 | -4.54 | -5.54 | 1.51 × 10^-08^ | Yes |
| ATPAF2 | 17 | 17880723 | 17942523 | 395849 | 7.99 × 10^-04^ | 898130 | 3.84 × 10^-06^ | -3.16 | -4.47 | -5.40 | 3.42 × 10^-08^ | Yes |
| DRG2 | 17 | 17991200 | 18011285 | 395511 | 9.85 × 10^-04^ | 898130 | 1.78 × 10^-05^ | -3.09 | -4.13 | -5.11 | 1.59 × 10^-07^ | Yes |
| MYO15A | 17 | 18012020 | 18083116 | 390744 | 4.90 × 10^-03^ | 898130 | 2.60 × 10^-05^ | -2.58 | -4.05 | -4.69 | 1.38 × 10^-06^ | Yes |
| PSMC3IP | 17 | 40724333 | 40729849 | 350957 | 2.29 × 10^-02^ | 898130 | 6.29 × 10^-06^ | -2.00 | -4.37 | -4.50 | 3.38 × 10^-06^ | Yes |
| KCNH6 | 17 | 61600695 | 61626338 | 378796 | 4.13 × 10^-03^ | 898130 | 3.81 × 10^-05^ | -2.64 | -3.96 | -4.66 | 1.55 × 10^-06^ | Yes |
| KDM4B | 19 | 4969125 | 5153606 | 397286 | 9.79 × 10^-05^ | 898130 | 4.86 × 10^-05^ | -3.72 | -3.90 | -5.39 | 3.54 × 10^-08^ | Yes |
| KLC3 | 19 | 45836692 | 45854778 | 400351 | 2.75 × 10^-06^ | 898130 | 5.04 × 10^-03^ | -4.54 | -2.57 | -5.03 | 2.42 × 10^-07^ | Yes |
| AC074212.3 | 19 | 46236509 | 46267792 | 400815 | 7.22 × 10^-04^ | 898130 | 1.47 × 10^-04^ | -3.19 | -3.62 | -4.81 | 7.45 × 10^-07^ | Yes |
| SIX5 | 19 | 46268043 | 46272484 | 402802 | 4.64 × 10^-04^ | 898130 | 3.25 × 10^-04^ | -3.31 | -3.41 | -4.75 | 1.00 × 10^-06^ | Yes |
| DMWD | 19 | 46286205 | 46296060 | 417639 | 3.48 × 10^-04^ | 898130 | 6.33 × 10^-05^ | -3.39 | -3.83 | -5.11 | 1.63 × 10^-07^ | Yes |
| RSPH6A | 19 | 46298968 | 46318577 | 416916 | 1.13 × 10^-04^ | 898130 | 3.26 × 10^-06^ | -3.69 | -4.51 | -5.80 | 3.41 × 10^-09^ | Yes |
| FOXA3 | 19 | 46367247 | 46377055 | 354161 | 4.41 × 10^-04^ | 898130 | 2.59 × 10^-04^ | -3.33 | -3.47 | -4.81 | 7.69 × 10^-07^ | Yes |

*AD: Alzheimer’s disease, GWAS: genome-wide association studies, T2D: type 2 diabetes*

*Stouffer’s Z-scores were calculated using equal weights across studies to avoid dominance of the larger T2D GWAS sample size.*

*Multiple entries for ACE reflect distinct gene definitions or transcript models used in the gene-based association analysis.*

*Genome-wide gene-based significance: P_gene_ < 2.64 × 10⁻⁶ (Bonferroni correction for 18,965 tested genes).*

*Gene-based Z-score signs summarise aggregate evidence and should not be interpreted as definitive per-variant directional concordance; locus-level signed genetic covariance is assessed with LAVA*

### Supplementary Table 21: Expression-driven shared causal genes between Alzheimer’s disease and type 2 diabetes

| **Gene** | **Qtl name** | **Probe ID** | **Probe Chr** | **Probe bp** | **AD** | | | | | | **T2D** | | | | | |
| --- | --- | --- | --- | --- | --- | --- | --- | --- | --- | --- | --- | --- | --- | --- | --- | --- |
|  |  |  |  |  | **Top SNP** | **B SMR** | **Se SMR** | **P SMR** | **P HEIDI** | **FDR p SMR** | **Top SNP** | **B SMR** | **Se SMR** | **P SMR** | **P HEIDI** | **FDR p SMR** |
| FIBP | GTEx (Brain Frontal Cortex BA9) | ENSG00000172500.12 | 11 | 65883921 | rs588114 | -0.043 | 0.011 | 1.75 × 10^-04^ | 0.32 | 4.36 × 10^-02^ | rs588114 | 0.13 | 0.03 | 1.27 × 10^-04^ | 0.01 | 6.42 × 10^-03^ |
| SNX32 | GTEx (Brain Cerebellar Hemisphere) | ENSG00000172803.17 | 11 | 65833641 | rs58568715 | -0.011 | 0.003 | 3.86 × 10^-05^ | 0.83 | 1.53 × 10^-02^ | rs58568715 | 0.03 | 0.01 | 1.26 × 10^-05^ | 0.01 | 1.17 × 10^-03^ |
| FIBP | eQTLGen (Whole Blood) | ENSG00000172500 | 11 | 65886136 | rs1204650 | -0.038 | 0.008 | 5.52 × 10^-06^ | 0.25 | 3.57 × 10^-03^ | rs1204650 | 0.11 | 0.03 | 6.14 × 10^-06^ | 0.01 | 7.15 × 10^-04^ |
| ACE | eQTLGen (Whole Blood) | ENSG00000159640 | 17 | 63487720 | rs4277405 | -0.142 | 0.034 | 2.89 × 10^-05^ | 0.17 | 1.27 × 10^-02^ | rs4277405 | 0.35 | 0.10 | 3.49 × 10^-04^ | 0.03 | 1.28 × 10^-02^ |
| PLEKHA1 | eQTLGen (Whole Blood) | ENSG00000107679 | 10 | 1.22E+08 | rs5013920 | 0.023 | 0.006 | 4.80 × 10^-05^ | 0.21 | 1.71 × 10^-02^ | rs5013920 | 0.12 | 0.02 | 4.89 × 10^-13^ | 0.04 | 9.48 × 10^-10^ |
| ACE | GTEx (Brain Cerebellum) | ENSG00000159640.15 | 17 | 63477061 | rs4308 | -0.027 | 0.007 | 1.35 × 10^-04^ | 0.48 | 3.61 × 10^-02^ | rs4308 | 0.06 | 0.02 | 1.86 × 10^-03^ | 0.06 | 3.75 × 10^-02^ |
| YPEL3 | GTEx (Brain Cerebellar Hemisphere) | ENSG00000090238.11 | 16 | 30092314 | rs11642399 | -0.021 | 0.006 | 1.62 × 10^-04^ | 0.44 | 4.12 × 10^-02^ | rs11642399 | 0.08 | 0.02 | 1.87 × 10^-06^ | 0.07 | 2.80 × 10^-04^ |
| PPP4C | eQTLGen (Whole Blood) | ENSG00000149923 | 16 | 30080677 | rs12596543 | 0.045 | 0.011 | 3.65 × 10^-05^ | 0.11 | 1.47 × 10^-02^ | rs12596543 | -0.17 | 0.03 | 2.21 × 10^-07^ | 0.09 | 5.50 × 10^-05^ |
| KAT8 | GTEx (Brain Cerebellar Hemisphere) | ENSG00000103510.19 | 16 | 31115754 | rs2855475 | -0.026 | 0.006 | 3.94 × 10^-05^ | 0.33 | 1.55 × 10^-02^ | rs2855475 | 0.07 | 0.02 | 3.83 × 10^-04^ | 0.10 | 1.35 × 10^-02^ |
| KAT8 | GTEx (Brain Hypothalamus) | ENSG00000103510.19 | 16 | 31115754 | rs61162043 | -0.027 | 0.007 | 1.98 × 10^-04^ | 0.67 | 4.64 × 10^-02^ | rs61162043 | 0.06 | 0.02 | 1.62 × 10^-03^ | 0.11 | 3.42 × 10^-02^ |
| KAT8 | GTEx (Adrenal Gland) | ENSG00000103510.19 | 16 | 31115754 | rs9936329 | -0.018 | 0.005 | 1.12 × 10^-04^ | 0.11 | 3.14 × 10^-02^ | rs9936329 | 0.05 | 0.01 | 3.29 × 10^-04^ | 0.15 | 1.23 × 10^-02^ |
| KAT8 | eQTL (BrainMeta) | ENSG00000103510.20 | 16 | 31122941 | rs9925964 | -0.016 | 0.004 | 6.80 × 10^-06^ | 0.11 | 4.15 × 10^-03^ | rs9925964 | 0.04 | 0.01 | 3.74 × 10^-04^ | 0.18 | 1.33 × 10^-02^ |
| INO80E | eQTL (BrainMeta) | ENSG00000169592.15 | 16 | 30000754 | rs4787489 | 0.017 | 0.004 | 8.46 × 10^-05^ | 0.39 | 2.65 × 10^-02^ | rs4787489 | -0.07 | 0.01 | 1.77 × 10^-08^ | 0.20 | 7.63 × 10^-06^ |
| BCKDK | eQTLGen (Whole Blood) | ENSG00000103507 | 16 | 31109449 | rs4889619 | 0.040 | 0.009 | 6.73 × 10^-06^ | 0.15 | 4.15 × 10^-03^ | rs4889619 | -0.09 | 0.03 | 3.04 × 10^-04^ | 0.27 | 1.16 × 10^-02^ |
| ZNF668 | eQTLGen (Whole Blood) | ENSG00000167394 | 16 | 31067542 | rs2303222 | 0.029 | 0.006 | 6.76 × 10^-06^ | 0.03 | 4.15 × 10^-03^ | rs2303222 | -0.06 | 0.02 | 1.27 × 10^-03^ | 0.35 | 2.95 × 10^-02^ |
| BCKDK | GTEx (Whole_Blood) | ENSG00000103507.13 | 16 | 31106107 | rs749671 | 0.084 | 0.022 | 1.37 × 10^-04^ | 0.58 | 3.64 × 10^-02^ | rs749671 | -0.18 | 0.06 | 2.62 × 10^-03^ | 0.37 | 4.64 × 10^-02^ |
| HSD3B7 | eQTLGen (Whole Blood) | ENSG00000099377 | 16 | 30987177 | rs4889606 | 0.051 | 0.011 | 7.42 × 10^-06^ | 0.02 | 4.48 × 10^-03^ | rs4889606 | -0.11 | 0.03 | 1.36 × 10^-03^ | 0.46 | 3.07 × 10^-02^ |
| HSD3B7 | GTEx (Whole Blood) | ENSG00000099377.13 | 16 | 30988271 | rs4889606 | 0.055 | 0.014 | 7.68 × 10^-05^ | 0.04 | 2.45 × 10^-02^ | rs4889606 | -0.11 | 0.04 | 2.77 × 10^-03^ | 0.52 | 4.81 × 10^-02^ |
| INO80E | eQTLGen (Whole Blood) | ENSG00000169592 | 16 | 30000754 | rs9925915 | 0.029 | 0.007 | 4.15 × 10^-05^ | 0.10 | 1.59 × 10^-02^ | rs9925915 | -0.12 | 0.02 | 2.51 × 10^-09^ | 0.53 | 1.68 × 10^-06^ |
| INO80E | GTEx (Brain Cerebellum) | ENSG00000169592.14 | 16 | 29995294 | rs11150577 | 0.020 | 0.005 | 2.17 × 10^-04^ | 0.53 | 4.96 × 10^-02^ | rs11150577 | -0.09 | 0.02 | 1.09 × 10^-06^ | 0.54 | 1.91 × 10^-04^ |
| GALNT10 | eQTL (BrainMeta) | ENSG00000164574.16 | 5 | 1.54E+08 | rs9324772 | 0.034 | 0.008 | 2.15 × 10^-05^ | 0.36 | 1.01 × 10^-02^ | rs9324772 | -0.07 | 0.02 | 1.89 × 10^-03^ | 0.65 | 3.79 × 10^-02^ |

### Supplementary Table 22: Methylation-driven shared causal genes between AD and T2D

| **Gene** | **Qtl name** | **Probe ID** | **Probe Chr** | **Probe bp** | **AD** | | | | | **T2D** | | | | |
| --- | --- | --- | --- | --- | --- | --- | --- | --- | --- | --- | --- | --- | --- | --- |
|  |  |  |  |  | **Top SNP** | **B SMR** | **Se SMR** | **P SMR** | **FDR p SMR** | **Top SNP** | **B SMR** | **Se SMR** | **P SMR** | **FDR pSMR** |
| GALNT10 | mQTL_BrainMeta | cg17852245 | 5 | 154308627 | rs6890748 | 0.030 | 0.008 | 1.23 × 10^-04^ | 2.27 × 10^-02^ | rs6890748 | -0.063 | 0.020 | 2.13 × 10^-03^ | 2.36 × 10^-02^ |
| MUS81 | mQTL_BrainMeta | cg04761267 | 11 | 65853673 | rs585557 | 0.008 | 0.002 | 1.25 × 10^-05^ | 4.88 × 10^-03^ | rs585557 | -0.024 | 0.005 | 5.80 × 10^-06^ | 3.37 × 10^-04^ |
| CCDC85B | mQTL_McRae | cg22360649 | 11 | 65890262 | rs2231884 | 0.023 | 0.005 | 6.85 × 10^-06^ | 3.07 × 10^-03^ | rs2231884 | -0.065 | 0.015 | 9.35 × 10^-06^ | 4.91 × 10^-04^ |
| HIRIP3 | mQTL_BrainMeta | cg02492205 | 16 | 30005454 | rs4283241 | -0.007 | 0.002 | 4.57 × 10^-05^ | 1.11 × 10^-02^ | rs4283241 | 0.031 | 0.005 | 2.63 × 10^-09^ | 7.08 × 10^-07^ |
| DOC2A | mQTL_McRae | cg06334689 | 16 | 30007399 | rs12921753 | 0.007 | 0.002 | 1.47 × 10^-05^ | 5.37 × 10^-03^ | rs12921753 | -0.032 | 0.005 | 1.15 × 10^-10^ | 4.09 × 10^-08^ |
| C16orf92 | mQTL_McRae | cg06326092 | 16 | 30023166 | rs4788213 | -0.017 | 0.004 | 3.55 × 10^-05^ | 9.51 × 10^-03^ | rs4788213 | 0.066 | 0.013 | 2.00 × 10^-07^ | 2.56 × 10^-05^ |
| DOC2A | mQTL_BrainMeta | cg06985993 | 16 | 30028734 | rs4788213 | 0.017 | 0.004 | 4.67 × 10^-05^ | 1.12 × 10^-02^ | rs4788213 | -0.063 | 0.012 | 3.87 × 10^-07^ | 4.29 × 10^-05^ |
| HSD3B7 | mQTL_McRae | cg04018474 | 16 | 30996682 | rs9796794 | -0.023 | 0.005 | 2.07 × 10^-05^ | 6.60 × 10^-03^ | rs9796794 | 0.048 | 0.016 | 1.99 × 10^-03^ | 2.24 × 10^-02^ |
| STX1B | mQTL_McRae | cg00249205 | 16 | 31000942 | rs2303222 | -0.028 | 0.007 | 3.20 × 10^-05^ | 9.13 × 10^-03^ | rs2303222 | 0.059 | 0.019 | 2.01 × 10^-03^ | 2.25 × 10^-02^ |
| VKORC1 | mQTL_McRae | cg27583149 | 16 | 31094161 | rs749670 | -0.043 | 0.011 | 1.74 × 10^-04^ | 2.84 × 10^-02^ | rs749670 | 0.091 | 0.031 | 3.92 × 10^-03^ | 3.50 × 10^-02^ |
| BCKDK | mQTL_McRae | cg08374890 | 16 | 31105746 | rs1549293 | 0.040 | 0.010 | 5.63 × 10^-05^ | 1.23 × 10^-02^ | rs1549293 | -0.091 | 0.028 | 9.94 × 10^-04^ | 1.42 × 10^-02^ |
| KAT8 | mQTL_McRae | cg02220965 | 16 | 31116989 | rs1978487 | 0.015 | 0.003 | 6.52 × 10^-06^ | 2.97 × 10^-03^ | rs1978487 | -0.036 | 0.010 | 3.42 × 10^-04^ | 6.88 × 10^-03^ |
| CAMTA2 | mQTL_McRae | cg19427746 | 17 | 4986637 | rs113536807 | 0.010 | 0.003 | 1.07 × 10^-04^ | 2.02 × 10^-02^ | rs113536807 | 0.021 | 0.007 | 3.67 × 10^-03^ | 3.36 × 10^-02^ |
| ACE | mQTL_McRae | cg21657705 | 17 | 63497139 | rs4353 | -0.026 | 0.006 | 1.23 × 10^-05^ | 4.88 × 10^-03^ | rs4353 | 0.078 | 0.018 | 1.22 × 10^-05^ | 5.98 × 10^-04^ |

### Supplementary Table 23: Methylation-driven shared genes (in linkage) between AD and T2D within the APOE region

| **Gene** | **Qtl name** | **Probe ID (CpG sites)** | **Probe Chr** | **Probe bp** | **AD** | | | | | | **T2D** | | | | | |
| --- | --- | --- | --- | --- | --- | --- | --- | --- | --- | --- | --- | --- | --- | --- | --- | --- |
|  |  |  |  |  | **Top SNP** | **B SMR** | **Se SMR** | **P SMR** | **P HEIDI** | **FDR p SMR** | **Top SNP** | **B SMR** | **Se SMR** | **P SMR** | **P HEIDI** | **FDR p SMR** |
| APOE | mQTL_BrainMeta | cg14123992 | 19 | 44904611 | rs7259620 | 0.063 | 0.003 | 1.52 x 10^-129^ | 3.41 x 10^-54^ | 4.22 x 10^-125^ | rs7259620 | -0.019 | 0.006 | 1.29 x 10^-03^ | 8.86 x 10^-11^ | 1.67 x 10^-02^ |
| APOC1 | mQTL_McRae | cg05644480 | 19 | 44914763 | rs59325138 | -0.195 | 0.024 | 6.38 x 10^-16^ | 1.95 x 10^-12^ | 5.90 x 10^-12^ | rs59325138 | 0.069 | 0.024 | 4.30 x 10^-03^ | 2.77 x 10^-04^ | 3.70 x 10^-02^ |
| APOC1 | mQTL_BrainMeta | cg09379229 | 19 | 44914411 | rs5117 | 0.267 | 0.037 | 8.31 x 10^-13^ | 2.19 x 10^-10^ | 2.56 x 10^-09^ | rs5117 | -0.114 | 0.022 | 2.61 x 10^-07^ | 6.29 x 10^-04^ | 3.09 x 10^-05^ |
| NECTIN2 | mQTL_BrainMeta | cg11670000 | 19 | 44849693 | rs1871046 | -0.065 | 0.011 | 5.51 x 10^-09^ | 1.20 x 10^-07^ | 6.37 x 10^-06^ | rs1871046 | 0.082 | 0.023 | 3.05 x 10^-04^ | 3.00 x 10^-01^ | 6.40 x 10^-03^ |
| DMPK | mQTL_BrainMeta | cg10857774 | 19 | 45783425 | rs6509237 | 0.018 | 0.004 | 1.89 x 10^-05^ | 7.10 x 10^-04^ | 6.39 x 10^-03^ | rs8106955 | -0.019 | 0.005 | 2.54 x 10^-04^ | 9.43 x 10^-04^ | 5.68 x 10^-03^ |
| APOE | mQTL_McRae | cg06750524 | 19 | 44906698 | rs769449 | 0.479 | 0.063 | 1.83 x 10^-14^ | 1.22 x 10^-03^ | 8.46 x 10^-11^ | rs769449 | -0.198 | 0.037 | 7.48 x 10^-08^ | 0.02.05 x 10^-02^ | 1.18 x 10^-05^ |
